# Functionally Oriented Genetic Analyses Reveal Potential Transcriptomic and Neurological Mechanisms of Stuttering

**DOI:** 10.64898/2026.08.11.26359888

**Authors:** Alyssa C. Scartozzi, Hannah G. Polikowsky, Ting-Chen Wang, James T. Baker, Heather M. Highland, Lauren E. Petty, Phillip Lin, 23andMe Research Team, Robin M. Jones, Eric R. Gamazon, Chad D. Huff, Nancy J. Cox, Shelly Jo Kraft, Dillon G. Pruett, Jennifer E. Below

**Author notes:** Co-senior authors (Dillon G. Pruett, Jennifer E. Below).

## Abstract

Speech and language are fundamental to the human experience, allowing for the sharing of thoughts and emotions through the coordination of many neurological and linguistic systems. Disruptions in these processes can lead to speech and language disorders, including stuttering, which is characterized by prolongations, blocks, and repetitions of speech sounds. To date, almost 60 genome-wide significant loci have been associated with stuttering. Alas, most of these signals appear in non-coding regions of the genome and thus remain largely uncharacterized. In this study, we probed functionality by leveraging the largest genome-wide association studies (GWAS) of self-reported stuttering in individuals with European genetic ancestry (N case = 78,394, N control = 865,956). We performed transcriptome-wide association studies (TWAS), tested causal effects via Mendelian randomization (MR), and assessed neuroimaging features associated with stuttering. Stuttering was associated with the genetically regulated gene expression (GReX) of 2,875 significant gene-tissue pairs (236 independent signals). Many of these GReX genes were enriched for neurological processes, including synapse organization and nervous system development, and 12 genes were independently supported in a clinically ascertained stuttering cohort. Our MR analysis identified 150 unique causal stuttering genes (53 distinct signals). Additionally, our neuroimaging analysis identified stuttering-associated genetic signals functionally linked with basal ganglia, cerebellum, and superior longitudinal fasciculus neuroimaging features. Together, these findings characterize transcriptomic signatures of stuttering and illuminate the neurological mechanisms driving this complex trait.

## Introduction

Developmental stuttering is a common speech condition characterized by sound prolongations, syllable and word repetitions, and blocks that disrupt the forward progression of speech. Onset typically occurs between 2-5 years of age^1^, with 1% of the population continuing to stutter into adulthood with or without the aid of speech therapy^2^. Stuttering has a strong genetic influence, with twin-based heritability estimates ranging from 0.42 to 0.84^3–9^. To date, the majority of genetic studies on stuttering have focused on families or population isolates with high rates of developmental stuttering, identifying seven candidate causal stuttering genes with large effect estimates that are not generalizable to the general population^10–15^. Recent advancements in phenotyping tools and large-scale biobanks have emerged to explore the genetic influence of stuttering at the population level. Thus far, three genome-wide association studies (GWAS) have uncovered nearly 60 unique loci contributing to stuttering risk^16–18^. Together, these studies suggest that stuttering risk is influenced by both common and rare genetic variation^10–18^. However, the biological interpretation of the identified loci has been difficult, especially since most of the GWAS signals are in non-coding regions of the genome. Most notably, one of these studies includes over 1 million self-reported stuttering cases and controls^16^, creating opportunities for post-GWAS discovery. Functionally characterizing and prioritizing candidate genes that impact stuttering risk is a crucial next step toward elucidating the molecular etiology of stuttering, which currently remains understudied.

Since the inception of GWAS, various genomic tools have been developed to identify the biological mechanisms underlying GWAS signals, often with a focus on gene expression. For example, GWAS-identified SNPs are more likely to be expression quantitative trait loci (eQTLs) than minor allele frequency-matched SNPs, highlighting the importance of gene expression in trait variation^19^. However, measured gene expression is frequently confounded by environmental, temporal, and batch effects. Genetically regulated gene expression (GReX) may overcome these limitations by capturing the portion of gene expression variation explained by inborn regulatory genetic factors^20^. Furthermore, GReX methods allow for the imputation of gene expression in tissues critical to stuttering such as the brain, which cannot easily be directly sampled. Transcriptome-wide association studies (TWAS)^21^ leverage the GReX associated with a trait of interest to identify regulatory mechanisms while reducing the multiple-testing burden compared to GWAS. Joint-Tissue Imputation (JTI)^22^, a recently developed TWAS method, improves existing GReX imputation models by leveraging cross-tissue similarity in eQTL effects. TWAS, and specifically, JTI and its causal inference extension MR-JTI, have successfully been used to elucidate disease relevant biology for other complex traits, such as Parkinson’s Disease^23^, endometrial cancer^24^, and cardiometabolic traits^25^. In this study, we leveraged the largest GWAS of self-reported stuttering and performed TWAS utilizing JTI trained eQTL models and S-PrediXcan^22^ to identify genes for which GReX is associated with stuttering, followed by post hoc Mendelian randomization (MR)^26^ analysis to identify putatively causal stuttering-associated genes. Together, this integrative approach leverages gene expression to map genetic risk to the underlying biological mechanisms of stuttering.

Although there is no definitive consensus on the neurological mechanisms contributing to stuttering, there are numerous neural circuits associated with speech production that have been proposed as well as broader neurological differences related to cognitive, language, and emotional processes that have been identified, all of which are not necessarily mutually exclusive (for summary, see ^27,28^). Imaging studies have shown that individuals who stutter may exhibit: (i) decreased white matter integrity along parts of the left arcuate/superior longitudinal fasciculus (dorsal auditory tract)^29–31^, (ii) hemispheric asymmetry during speech production and language processing^32–35^, (iii) disruptions in the connections between the cerebral cortex, basal ganglia and thalamus (cortico-basal ganglia-thalamocortical loop)^36–38^, (iv) structural, function, or connectivity differences in the cerebellum^39–43^, (v) altered morphology and/or activation of areas in the limbic region such as larger nucleus accumbens^44^ and increased amygdala activation during speech tasks^45,46^. Resources that link GWAS with neuroimaging-derived phenotypes (NIDPs), including white matter tracts and structural features, provide an opportunity to map how genetic variants associated with stuttering might influence the neurological mechanisms of stuttering. These NIDPs allow for characterization of the molecular and genetic neurological pathways underlying trait and disease mechanisms^47,48^. BrainXcan^49^, which integrates GWAS summary statistics with neuroimaging data from the UK Biobank, including structural, diffusion, and functional MRI is an analysis approach built to probe the genetic underpinnings of NIDPs.

Our current study investigates the genetic etiology of stuttering risk using post-GWAS functional analyses. First, we performed TWAS^22^ using summary statistics from a large GWAS of self-reported stuttering^16^ across 49 different GTEx tissues^50^. Then, we performed gene-set enrichment analyses using the resulting statistically significant stuttering genes. We subsequently used a clinically ascertained stuttering cohort, the International Stuttering Project, to validate self-reported stuttering GReX results^17,51^. Next, we used BrainXcan^49^ to explore neuroimaging features associated with stuttering. Finally, to identify potential causal genes from our GReX findings, we performed debiased-inverse variance weighted Mendelian randomization (MR)^26^. Together, these efforts establish a functional genetic framework linking genetic risks to the regulatory and neurological etiology of stuttering.

## Results

### Study Overview

We performed transcriptome-wide association analyses using meta-analyzed GWAS results from Polikowsky, Scartozzi & Shaw *et al.*, focusing on the predominantly European genetic ancestry self-reported stuttering analyses due to the disproportionate size of this sample and similar genetic ancestry to GTEx^50^ and UK Biobank participants^49^. In this analysis, participants who reported ‘yes’ to the question ‘Have you ever had a stammer or stutter?’ were classified as cases, while those who reported ‘no’ were classified as controls. The meta-analyzed summary-level results used for the present analyses are based on male and female stratified GWAS consisting of a total of 78,394 cases and 865,956 controls^16^.

### TWAS Results for Self-Reported Stuttering

We performed TWAS analyses using S-PrediXcan with Joint-Tissue Imputation (JTI)-trained eQTL prediction models^22^ to investigate the association of genetically regulated gene expression (GReX) with self-reported stuttering across the 49 GTEx tissues^50^. We found GReX of 620 unique genes across all tissues, or 2,875 significant gene-tissue pairs, was associated with self-reported stuttering (*p*-value < 0.05, after FDR correction, Table 1 and Figure 1), resulting in 236 independent signals (all pairwise r^2^ < 0.3; see Methods, Supplementary Figures 6-27). The number of significant GReX genes per tissue is given in Supplementary Figure 1. Like many neurodevelopmental disorders, stuttering is a male-skewed condition, especially in adulthood^1,52^. The divergence in stuttering prevalence in males and females between early childhood and adulthood has historically fueled the hypothesis that sex-specific or sex-influenced genetic factors modulate risk^53^. Therefore, in addition to the primary analysis in men and women, we present sex-stratified analyses within the Supplementary Information (Supplementary Figures 1-4 and 29-52, Supplementary Tables 1-2).

**Figure 1.**
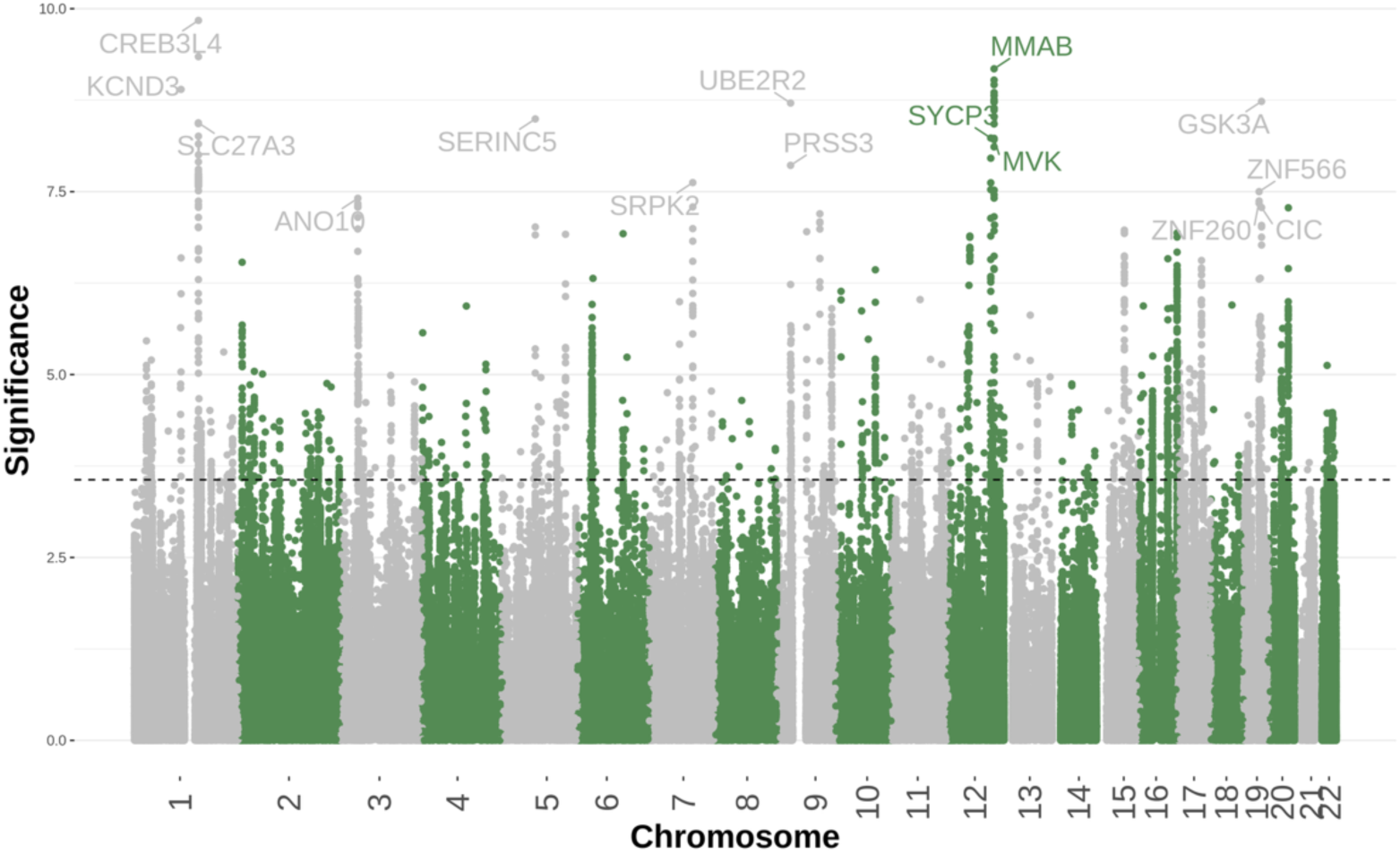
TWAS Results for Self-reported Stuttering Across 49 GTEx Tissues. Chromosome is displayed on the x-axis and −log(*p*-value) is seen on the y-axis. The dotted line represents the FDR significance threshold (FDR *p*-value < 0.05). The first instance of the top 15 unique genes are labeled.

**Table 1.** FDR Significant GReX Results for Stuttering. In excel sheet.

### Gene-set Enrichment Analysis

Using the 620 unique stuttering-associated genes, we conducted gene-set enrichment analyses using g:Profiler’s tool, g:GOSt^54^, prioritizing gene sets that were identified as driver terms (see Methods). We found that stuttering-associated genes were enriched for many neurological processes such as nervous system development, synapse organization, and axon initial segment (FDR adjusted *p*-value < 0.05). Driver term results can be found in Table 2. Full gene-set enrichment results can be found in Supplementary Table 3.

**Table 2.** Gene-set Enrichment Analysis Driver Terms for our FDR Significant GReX results. In excel sheet.

### Replication of Identified Genes

To address the limitation of self-reported phenotyping in our discovery analyses, we sought replication of the identified GReX genes within the International Stuttering Project, or the ISP, which is a clinically ascertained stuttering study^17^. To assess replication, we examined the 620 identified genes by querying ISP GReX association results presented in Pruett et al. (N case = 1,345, and N control = 6,759)^51^ and determined whether gene-tissue pairs surpassed a replicative significance threshold (*p*-value < 4.59 x 10^−4^, based on Bonferroni correction for the 620 tested genes). Twelve genes identified in our analysis replicated in the ISP study: *RABEP1*, *NLRP1*, *NUP88*, *C1QBP*, *EPHA4*, *RPAIN*, *PLB1*, *TMEM42*, *AC004148.2*, *AC079834.2*, *CTF1*, and *AL358472.2*. Across both studies, the GReX of *RABEP1* in brain caudate basal ganglia, brain nucleus accumbens basal ganglia, and brain cortex; GReX of *NUP88* in testis; GReX of *C1QBP* in the nerve tibial, brain cerebellum, whole blood, brain hypothalamus, heart left ventricle, esophagus gastroesophageal junction, brain cerebellar hemisphere, liver, testis, prostate, heart atrial appendage, artery tibial, esophagus muscularis, thyroid, brain cortex, brain cortex, lung, and adipose subcutaneous; GReX of *EPHA4* in the esophagus gastroesophageal junction, GReX of *RPAIN4* in the testis and cells cultured fibroblasts; GReX of *AC004148.2* in the cells cultured fibroblasts; GReX of *AC079834.2* in the esophagus gastroesophageal junction, esophagus muscularis, and colon transverse surpassed FDR significance in the self-reported stuttering TWAS (FDR *p*-value < 0.05) and replicative significance in the ISP TWAS (*p*-value < 4.59 x 10^−4^). Full replication results for the ISP can be found in Supplementary Table 4.

### Neuroimaging Features Associated with Transcriptomic Signatures of Self-Reported Stuttering

We used BrainXcan to identify brain imaging features associated with self-reported stuttering status. We found 43 features associated with stuttering (38 diffusion MRI [dMRI] and five structural MRI [T1]; FDR *p*-value < 0.05; Figure 2, Table 3). The five T1 structural imaging features associated with the trait were increased gray-cortical matter volume in the left occipital fusiform gyrus, increased gray-cerebellum volume in the vi cerebellum, increased gray-cortical volume in the left intracalcarine cortex, decreased gray-cortical volume in the paracingulate gyrus, and increased subcortical volume in the putamen (Figure 2a and Table 3). The dMRI microstructural metrics consisted of mean intra-cellular volume fraction (ICVF), weighted-mean ICVF (w-ICVF), orientation dispersion index (OD), weighted-mean orientation dispersion index (w-OD), fractional anisotropy (FA), and w-ISOVF (weighted mean isotropic or free water volume fraction) measures in the following white matter tracts: right anterior corona radiata (OD), left (ICVF) and right (ICVF and FA) cingulum hippocampus, right external capsule (FA), forceps major (w-ISOVF), fornix (FA), left and right crus of the fornix stria terminalis (ICVF), left (ICVF) and right (ICVF) inferior cerebellar peduncle, left inferior longitudinal fasciculus (w-ICVF), left (w-ICVF) and right (ICVF) medial lemniscus, middle cerebellar peduncle (ICVF and w-ICVF), left (w-ICVF) and right (w-ICVF) parahippocampal part of the cingulum, PC-OD-Tract-Based Spatial Statistics(‘TBSS’)-1 (OD), left (ICVF and OD) and right (ICVF and OD) posterior corona radiata, left posterior thalamic radiation (ICVF), splenium of the corpus callosum (ICVF), left (ICVF) and right (ICVF) superior corona radiata, left superior fronto-occipital fasciculus (ICVF), left (ICVF and w-ICVF) and right (ICVF, w-ICVF and w-ISOVF) superior longitudinal fasciculus, left superior thalamic radiation (w-OD), left (ICVF) and right (ICVF and OD) tapetum, and left uncinate fasciculus (ICVF) (Figure 2b and Table 3). Many of these brain regions and white matter tracts relate to facilitating sensory and motor information integration (i.e., posterior and superior corona radiata^55^ and superior thalamic radiation^56^), processing visual information (i.e., inferior longitudinal fasciculus^57^), bilateral connectivity (i.e., tapetum and splenium of the corpus callosum^58^), and speech motor learning (i.e., cerebellum^59^ and putamen^60^).

**Figure 2.**
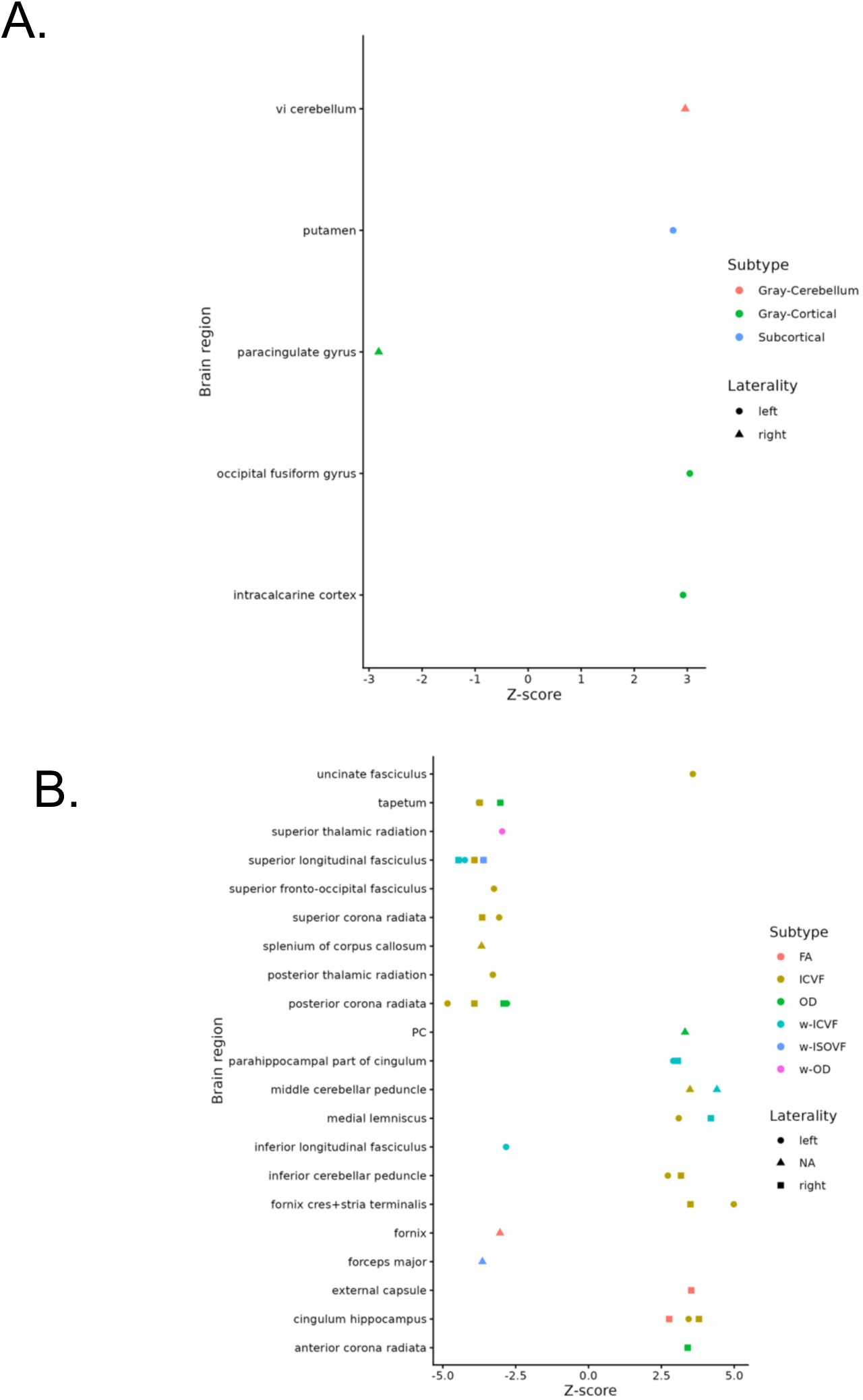
FDR significant T1 structural imaging (A) and diffusion MRI (B) associations with stuttering via BrainXcan. The y-axis represents the T1 structural brain region or the dMRI functional white matter tract. The x-axis represents the z-scores of region-specific feature associations with stuttering risk in using GWAS results. ICVF: intra-cellular volume fraction, w-ICVF: weighted-mean ICVF, OD: orientation dispersion index, w-OD weighted-mean OD, FA: fractional anisotropy, w-ISOVF: weighted-mean isotropic or free water volume fraction. FDR *p*-value < 0.05.

**Table 3.** FDR significant neuroimaging features (T1 and dMRI) associated with stuttering risk via BrainXcan. In excel sheet.

### Mendelian Randomization

To identify genes with putative causal effects on regulation, we performed debiased-inverse variance weighted Mendelian randomization^26^ on our TWAS results. One-hundred and fifty unique genes, comprising 365 gene-tissue pairs, from our analysis showed significant causal effects after filtering criteria based on having more than five eQTLs used, an FSTAT > 10, and surpassing FDR correction across the 49 GTEx tissues^50^, resulting in 53 independent signals (see Methods, Table 4). Additional Mendelian randomization results using MR-Egger and Weighted Median^61^ can be found within Supplementary Table 5.

**Table 4.** Debiased-Inverse Variance Weighted MR Significant Results for Stuttering. In excel sheet.

## Discussion

The current study expands exploration of the genetic etiology of stuttering by adopting the TWAS approach alongside causal inference, identifying novel genes that have not been previously reported for stuttering. Overall, our analyses revealed significant associations of GReX of 2,875 gene-tissue pairs (620 unique genes) with stuttering, corresponding to 236 independent signals (all pairwise r^2^ < 0.3). Many of these genes have been implicated in neuronal signaling^62–64^, and have been previously associated with traits comorbid with stuttering, including sleep- and sleep-related disorders^65,66^, depression^67–71^, and weight^65^. Discussed in greater detail below, the stuttering-associated genes we identified are enriched in gene-sets spanning many neurological and developmental processes, including terms for synapse and nervous system development. We further show independent support for twelve GReX genes using a clinically ascertained stuttering dataset. Next, by examining NIDPs influenced by stuttering by leveraging BrainXcan^49^, our results are convergent with prior neuroimaging studies on stuttering, specifically showing involvement of the basal ganglia^44,72–74^, cerebellum^39–43^, and superior longitudinal fasciculus^29–31^ Finally, Mendelian randomization analyses identified 150 unique putative causal stuttering genes, resulting in 53 independent signals (all pairwise r^2^ < 0.3). In sum, these efforts begin to elucidate and characterize how gene expression differences impact stuttering and highlight potential connections between these transcriptomic signatures and neuroimaging brain features.

Our top TWAS genes have been implicated in many biological processes, including neuronal signaling and behaviors (*KCND3*^62^, *CIC*^63^, and *GSK3*A^64^), ion transport and signaling (*ANO10*^75^ and *PRSS3*^76^); metabolic and lipid pathways (*MMAB*^77^, *MVK*^78^, *SLC27A3*^79^ and *SERINC5*^80^); DNA binding, transcriptional regulation and signaling (*CREB3L4*^81^, *CIC*^63^, *UBE2R2*^82^*, ZNF566*^83^, *ZNF260*^84^, and *SYCP3*^85^). Moreover, many of these genes have been previously associated with traits comorbid with stuttering, including sleep-related traits and disorders (i.e., insomnia (*ANO10*^86^, *MVK*^86^, *MMAB*^86,87^, and *ZNF566*^86^, short sleep duration (*MVK*^88^ and *MMAB*^88^) chronotype (*SLC27A3*^89^), Indeed, an Electronic Health Record (EHR) review^65^ found broadly defined sleep disorders associated with stuttering, with other additional studies showing an association between stuttering and insomnia in children^66^. Furthermore, many genes were implicated in other comorbid traits spanning obesity/weight^65^ and depression^67–71^, such as BMI (*MMAB*^90^, *KCND3*^91–95^, *UBE2R2*^90,91,96^, *MVK*^91–93,96,97^), and *ANO10*^93,97^), depression (*ANO10*^98^), depressive symptoms (*MMAB*^99^), and major depressive disorder (*ANO10*^100^). More broadly, many neurological and developmental processes were significant in gene-set enrichment analyses for our significant TWAS results, including the driver terms of synapse and nervous system development. Other significant gene-set enrichments included terms spanning synaptic signaling and transmission, synaptic vesicle functions, neurogenesis, developmental processes, neurotransmitter secretion, and more (Table 2 and Supplementary Table 3). Furthermore, many significant TWAS results were found in stuttering-related and adjacent tissues (i.e., esophagus muscularis, lung, brain cerebellum). Together, these results point to tissue biology relevant to fluent speech production, which requires somatosensory coordination between the respiratory system, relevant anatomical components (*i.e.*, tongue, lips), as well as brain regions involved in auditory processing and motor planning.^101^ The cerebellum has been implicated in stuttering^39–43^ and supports orofacial movement and speech motor control^59^. Thus, these functional studies of genetic risk factors for stuttering provide convergent evidence for key neurological systems relevant for stuttering.

We further found that twelve of our stuttering-associated genes independently replicated in the International Stuttering Project, a clinically ascertained stuttering cohort. Many of these genes have been found to be involved in intracellular membrane trafficking (*RABEP1*^102^), immune responses (*NLRP1*^103^, *C1QBP*^104^, and *CTF1*^105^), neural signaling (*EPHA4*^106^), lipid metabolism (*PLB1*^107^) and nuclear transport (*RPAIN*^108^ and *NUP88*^109^). Further, these genes have been previously found to be linked to traits associated with stuttering comorbidities^16,65,67–71,110,111^. Specifically, *RABEP1*^90,91^, *NLRP1*^91^, and *NUP88*^90,91^ have previously been associated with BMI, and *RABEP1*^112^ and *NUP88*^112^ have been associated with obesity. An EHR review^65^ found both overweight/obesity and symptoms concerning nutrition were associated with stuttering, suggesting differences in weight regulation. Additionally, *TMEM42* was previously found to be associated with major depressive disorder^100^, aligning with studies that have shown that individuals who stutter are more likely to experience depression^67–71^. *PBL1* was found to be previously associated with age of onset of allergic disease (asthma, hay fever and/or eczema)^113^, where the prevalence of allergies and asthma have been identified in children^110^ and adults who stutter^111^. *EPHA4* was previously found to be associated with cortical surface area^114,115^ and thickness^114^, as well as alcohol consumption (i.e., drinks per week)^116^. Previous neuroimaging studies have shown decreased cortical thickness in ventral motor and premotor areas of the left hemisphere in children who stutter^117^. Furthermore, the GWAS study underlying the TWAS presented here also showed genetic correlations of BMI, depression and alcohol consumption with stuttering^16^. The biological mechanisms underlying these traits and stuttering remain elusive, highlighting potentially notable comorbidities worthy of functional follow-up.

More broadly, our BrainXcan findings implicated brain pathways involved in speech production and highlighted their genetic contributions to stuttering. Specifically, we found neuroimaging features of the superior longitudinal fasciculus associated with stuttering. We found decreased ICVF and w-ICVF in the left and right superior longitudinal fasciculus (SLF), suggesting lower neurite or axonal density. We also observed decreased w-ISOVF in the right SLF, indicating reduced extracellular free water. The SLF is a major white matter tract that connects the perisylvian areas in the ipsilateral hemisphere (i.e., the frontal, temporal, and parietal lobes)^118^. As part of the dorsal auditory tract^119^, the SLF connects speech motor and auditory regions, integrating auditory processing and speech motor networks ^120^. Consistent with previous studies showing decreased white matter integrity in the SLF in both children and adults who stutter^29–31,121^, our results extend these neuroimaging findings by showing that SLF microstructural differences are associated with genetic factors associated with stuttering.

Furthermore, our BrainXcan results revealed increased volume of the left putamen associated with stuttering. The putamen is a subcortical structure within the dorsal striatum of the basal ganglia^122^ and plays key roles in learning, motor control^60^ and language processing^123,124^. Previous literature has reported reduced putamen volume in children with persistent stuttering^12^5. By contrast, adults with persistent stuttering exhibited increased neural activation in the putamen^126^ and caudate nucleus^127^ (two components of the basal ganglia). More broadly, the basal ganglia is involved in motor relay^128^, which is important for the voluntary speech motor movements required for communication. Specifically, the basal ganglia receive motor, sensory, and cognitive input from multiple regions of the cerebral cortex and projects to the thalamus, which facilitates cortical activation needed for speech movements^128^. Notably, the basal ganglia is part of the cortico-basal ganglia-thalamocortical loop, which has been implicated in stuttering^36,37^. Indeed, neuroimaging studies have shown differences in the basal ganglia of individuals who stutter^44,72–74^. Together, these results highlight the need for further investigations into the connection between the basal ganglia and stuttering.

Lastly, we found increased gray-cerebellum volume of the right vi cerebellum associated with stuttering. The right cerebellar hemisphere primarily connects to the left cerebral hemisphere, supporting functions such as orofacial movement and speech motor control^59^. Further evidence also suggests that the cerebellum is involved in higher-order cognitive processes, including working memory, semantic judgement, spatial processing, procedural learning, and decision making^129^. Many studies have found structural, functional, or connectivity differences involving the cerebellum of people who stutter^39–43^. Functionally, the cerebellar-cortical loops seem to support self-initiated movements and adjustment of those movements in response to perceived error, suggesting that this function may be affected in people who stutter^59^. Overall, the cerebellar neuroimaging features identified in this study further corroborate and extend previous neuroimaging findings in stuttering.

Our study has several limitations. Specifically, as the phenotype in our discovery analyses is self-reported stuttering it may represent stuttering more broadly, including other non-developmental forms of stuttering and individuals who may have recovered from stuttering. However, this study provides the foundation to perform follow-up studies with deeper phenotyping, including better characterization of recovery status and severity measures. Given that developmental stuttering is most common in young children, we note that the JTI^22^ and BrainXcan^49^ models were trained using data from adults. Ideally, GReX models and related resources would be derived from children; however, no such models or resources are currently available. In the future, the development of newer resources, such as datasets with children or adolescents (i.e., developmental GTEx^130^) which reflect the developmental period closer to stuttering onset may better inform these analysis. Although stuttering is a highly sexually dimorphic trait ^1,52^, we utilized sex-combined JTI tissues models for our TWAS analyses^22,50^. The creation of sex-stratified GReX resources may allow for further explorations into sex-specific biological mechanisms. Due to this limitation, we cannot thoroughly investigate sex-specific mechanisms contributing to stuttering in both males and females. Research into sex-specific mechanisms represents an important next step for future research.

In sum, this study advances our understanding of the functional genetic etiology of stuttering by integrating transcriptome-wide and Mendelian randomization approaches, enhancing the discovery of stuttering-associated genes. Our findings reveal that the GreX of stuttering-associated genes are enriched for neurological processes, with 12 of these genes replicated in an independent clinically ascertained stuttering cohort, and also offer convergent support for neuroimaging circuits and brain regions previously implicated in stuttering. Together, these findings not only deepen our mechanistic insight into the genetic basis of stuttering but also establish a foundation for future investigation into specific neurobiological pathways driven by genetic factors influencing this complex trait.

## Methods

### Description of Datasets

23andMe Research Institute: We utilized genome-wide association studies of European ancestry male and female research participants of 23andMe Research Institute. Case status was defined by participants’ answer to the question “Have you ever had a stutter or stammer?” Participants who answered “yes” were classified as cases and participants who answered “no” were classified as controls. See Polikowksy, Scartozzi, and Shaw *et al.* for more information on this study and genotyping information^16^.

International Stuttering Project (ISP): To validate GReX findings from our self-reported stuttering phenotype, we used GReX association results from the International Stuttering Project, an internal stuttering cohort. See Polikowsky *et al.* for a detailed description of this study and genotyping information. Genome-wide GReX findings for this study have been reported in a previous manuscript (see ^51^).

### Joint Tissue Imputation

To investigate the molecular etiology associated with stuttering, we used an extension of S-PrediXcan, Joint-Tissue Imputation (JTI)^22^, which leverages cross-tissue similarity to predict GReX association with self-reported stuttering (N case = 78,394, N control = 865,956). JTI tissue and covariance models were generated using transcriptome data from the Genotype-Tissue Expression study (GTEx) v8^131^. As recommended by Zhou et al.^22^, we utilized models for all 49 available tissues, agnostic to potential biological relevance. To determine significance, we applied a within-study FDR correction across all genes and tissues for each sex-stratified analysis; any adjusted *p*-value < 0.05 was considered significant.

To distinguish independent signals in our JTI results, we computed the Pearson’s correlation coefficient for each pair of individual-level gene-tissue GReX values, to evaluate co-expressed and independent signals based on clustering. Clustering investigations were performed in a chromosome-specific manner. Since we did not have access to the 23andMe Research Institute’s individual-level data, we predicted the individual-level GReX for all identified genes using the 1000 Genomes Project EUR superpopulation genetic data^132^. Then, we created a correlation matrix for each gene-tissue pair per chromosome. Next, we leveraged IMUS^133^ to determine the maximum related set of gene-tissue pairs using an r^2^ of 0.3. Gene-tissue results not found within the maximum related sets were determined to be independent. Heatmaps showing GReX correlation per chromosome for our analyses can be found in Supplementary Figures 6-27. Due to the sexual dimorphic nature of stuttering^1,52^, we present sex-stratified TWAS results within the Supplementary Information file.

### Gene-set Enrichment Analysis via g:GOSt

To better understand the types of biological processes enriched in our GReX genes, we performed gene-set enrichment analyses using g:Profiler’s tool, g:GOSt^54^. This online tool performs statistical enrichment analysis on user-defined gene lists using multiple sources of functional evidence (i.e., Gene Ontology terms, biological pathways, regulatory motifs, human disease annotation, and protein-protein interactions). To run this analysis, we provided our input gene list of the 620 unique stuttering GReX genes and selected humans as our organism. All gene-sets were determined to be significant if they surpassed an FDR-adjusted *p*-value < 0.05. Gene-sets were prioritized as driver terms through g:GOSt’s two-stage filtering algorithm that first groups significant GO terms into related sub-ontologies and then applies a simple greedy search to identify leading gene-sets, adjusting *p*-values accordingly.

### GReX Replication in the ISP

To validate our findings beyond a self-reported stuttering discovery phenotype, we sought to replicate our significant GReX genes using TWAS results generated from the International Stuttering Project, a clinically ascertained stuttering study.^51^. Replication analyses were performed by querying the 620 unique GReX genes within the ISP set (replication-based *p*-value of 8.065 x 10^−5^ [0.05/620 unique genes]) agnostic of tissue matching.

### Probing Neuroimaging Features

To examine neuroimaging features associated with stuttering, we performed BrainXcan using our self-reported stuttering results. BrainXcan was run using the following default parameters: ridge NIDP prediction model type, residual set of NIDPs, and a CV spearman cutoff for models of 0.1. BrainXcan is an extension of S-PrediXcan, which infers genetically predicted structural brain features from trained NIDP models^49^. Results were considered significant if they had an FDR-adjusted *p*-value < 0.05.

### Mendelian Randomization

To assess the causality of the gene-tissue results identified by TWAS, we performed debiased-inverse variance weighted Mendelian randomization (MR)^26^ on our significant TWAS results (2,875 gene-tissue pairs, Figure 1 and Table 1). All instrumental variables were obtained using the GTEx v8 tissue-specific eQTL target for the gene of interest^131^. We performed LD clumping using the 1000 Genomes Project EUR superpopulation reference panel^132^ and bigsnpr R package^134^, since we do not have individual-level data available for our discovery analyses. Next, an FDR correction was applied to our MR results. MR gene-tissue pairs were considered significant if: (i) there were more than five eQTLs used, (ii) the FSTAT was > 10, and (iii) the FDR adjusted *p*-value was < 0.05, resulting in the significance of 365 gene-tissue pairs. To better understand the impact of method assumptions on our MR results, we also performed Mendelian randomization using MR-Egger and Weighted Median methods, which can be found in Supplementary Table 7, respectively.

Similar to the clustering used in our GReX results, we identified independent signals by computing Pearson’s correlation coefficient for each MR-significant gene-tissue pair, leveraging individual-level data from the 1000 Genomes Project EUR superpopulation genetic data^132^. Next, we created a correlation matrix of GReX for each gene-tissue pair within a given chromosome. To determine the maximum related set of gene-tissue pairs, we used IMUS^133^ and an r^2^ of 0.3. Gene-tissue results not found within the IMUS maximum related sets were counted as independent. Heatmaps showing GReX correlation per chromosome for the MR analyses can be found within Supplementary Figures 53-69.

## Ethics

We have complied with all ethical guidelines. All participants provided informed consent to participate in the research. This study has been approved by Vanderbilt IRB (181575 and 180583).

## Acknowledgements

We would like to thank the research participants and employees of 23andMe Research Institute for making this work possible as well as participants in the International Stuttering Project. This research was supported by NIH grants from the National Institutive on Deafness and Other Communication Disorders (NIDCD) to Vanderbilt University Medical Center and Wanye State University (1R03DC015329 and R01DC017175) and to Vanderbilt University Medical Center (5R21DC016723 and R01DC020311) supporting H.G.P., D.M.S., J.E.B., E.J.L., S.K., D.G.P., and R.M.J. D.G.P. received funding from the National Center for Advancing Translational Sciences of the NIH under Award Number TL1TR002244. A.C.S. received funding from the NIDCD under the Award Number F31DC022482. The content is solely the responsibility of the authors and does not necessarily represent the official views of any funding agencies.

## Author Contributions

J.E.B. and D.G.P. oversaw the entire study. J.E.B. and S.J.K. conceived the study. A.C.S. and D.G.P. drafted the manuscript, with contributions from H.G.P., L.E.P., T.C.W., H.M.H., and J.T.B. R.M.J., C.D.H., E.R.G., N.J.C., L.E.P., and H.G.P. provided insights on data analysis and interpretation of results. A.C.S. primarily performed all analyses, with assistance from T.C.W., J.T.B., L.E.P., and P.L. All authors critically reviewed the manuscript.

## Data and Code Availability

GWAS summary statistics for stuttering will be made available through the 23andMe website to qualified researchers under agreement with 23andMe that protects the privacy of the 23andMe participants. Interested investigators should visit the 23andMe Publication Dataset Access Program at https://research.23andme.com/dataset-access.

