## Supplementary Information for "Functionally Oriented Genetic Analyses Reveal Potential Transcriptomic and Neurological Mechanisms of Stuttering"

**Results**

***TWAS Results for Self-Reported Stuttering***

Using our sex-stratified self-reported stuttering results, we performed S-PrediXcan using JTI[1] eQTL tissue models to investigate genetically regulated gene expression (GReX) of female and male self-reported stuttering across the 49 available human tissues. In the female analysis, we found the GReX of 109 unique genes across all tissues, or 400 significant gene-tissue pairs, was associated with female self-reported stuttering ($p$-value < 0.05, after FDR correction, Supplementary Table 1 and Supplementary Figure 2), resulting in 36 independent signals (see Supplementary Methods). In the male analysis, we found the GReX of 33 unique genes across all tissues, or 75 gene-tissue pairs, was associated with male self-reported stuttering ($p$-value < 0.05, after FDR correction, Supplementary Table 2 and Supplementary Figure 3), resulting in 23 independent signals (see Supplementary Methods). The number of significant GReX genes per tissue for female and male GReX results can be found within Supplementary Figures 4 and 5, respectively.

**Methods**

***Sex-Stratified Self-reported Stuttering Summary Statistics***

Since the phenotype of stuttering is sexually dimorphic, where females are more likely to recover than males[2,3], we performed transcriptome-wide association analyses using the predominantly European genetic ancestry male and female self-reported stuttering GWAS results from [4]. Cases were individuals who reported 'yes' to the question 'Have you ever had a stammer or stutter?', while controls were those who reported 'no.' The female summary-level results consisted of 40,137 cases and 529,934 controls. The male summary-level results consisted of 38,257 cases and 336,022 controls.

***Joint Tissue Imputation***

Due to the sexually dimorphic nature of stuttering as a phenotype[2,3] as well as differences in top GWAS signals between males and females[4], we report sex-stratified TWAS results. Similar to our sex-combined results, we ran S-PrediXcan using JTI[1] to impute the genetically regulated gene expression (GReX) associated with male and female self-reported stuttering across the 49 GTEx tissues[5]. We applied a within-study FDR correction across all genes and tissues for each sex-stratified analysis. An FDR adjusted $p$-value < 0.05 was considered significant.

Using the same pipeline to identify independent signals in our sex-combined TWAS analyses, we calculated the Pearson's correlation coefficient between all individual-level gene-tissue pairs, allowing us to separate co-expressed and independent signals using a clustering approach for each chromosome. Since we did not have access to individual-level data from 23andMe Research Institute, we leveraged the 1000 Genomes Project EUR superpopulation genetic data[6] to impute GReX. After, correlation matrices were generated for each gene-tissue pair per chromosome. Next, to determine the maximum related set of gene-tissue pairs, we used IMUS[7] and an $r^2$ of 0.3. Gene-tissue pairs not identified within the maximum related set were deemed independent. Heatmaps showing chromosome-specific GReX correlations can be found in Supplementary Figures 29-46 for our female analyses and Supplementary Figures 47-52 for our male analysis.

Since we performed our analyses using sex-stratified summary statistics, but used sex-combined transcriptome JTI weights[1], we further probed sex-specificity of our results. Here, we leveraged previous investigations looking at sex-specific gene regulation and expression in the GTEx v8 dataset across 44 tissues[8]. Within Olivia *et al.*, authors discovered a total of 13,294 sex-biased differentially expressed genes across tissues. Using results available in their Supplementary Tables, we were able to see if any of the top 500 sex-biased gene-expressed genes across the 44 tissues were identified within our tissue-specific GReX results (Supplementary Tables 1 and 2).


### Supplementary Tables

In excel sheet.
**Supplementary Table 1. FDR Significant GReX Results for Stuttering in Females.**
*Indicate top 500 tissue-specific sex-biased expressed genes reported in Olivia *et al.*

In excel sheet.
**Supplementary Table 2. FDR Significant GReX Results for Stuttering in Males.**
*Indicate top 500 tissue-specific sex-biased expressed genes reported in Olivia *et al.*

In excel sheet.

**Supplementary Table 3. Full Gene-set Enrichment Results using the 620 unique GReX genes.**

In excel sheet.

**Supplementary Table 4. Replication Results from the ISP GReX Analysis.**
Results surpassed a replication-based *p*-value < $4.59 \times 10^{-4}$, based on Bonferroni correction for the 620 tested genes.

In excel sheet.
**Supplementary Table 5. Other Types of Mendelian randomization Methods on the GReX results.**

### Supplementary Figures

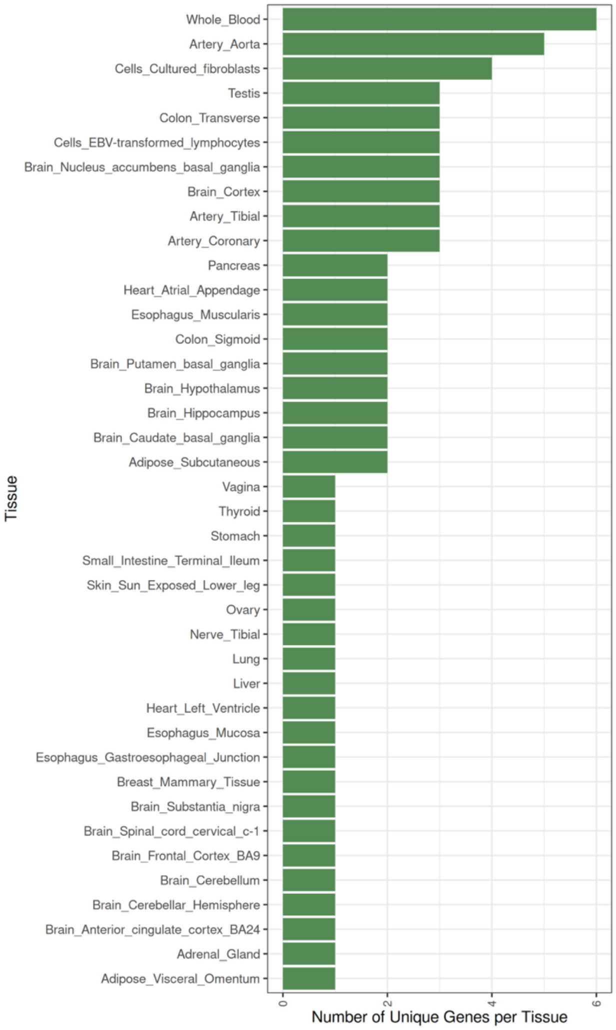


**Supplementary Figure 1. Histogram of Number of Unique GReX Genes per Tissue in Sex-combined Stuttering.** The number of unique GReX genes is depicted on the x-axis and tissue type is found on the y-axis. Note: the number of results that map to specific tissue will be influenced by the sample size of the eQTL discovery in GTEx[5].

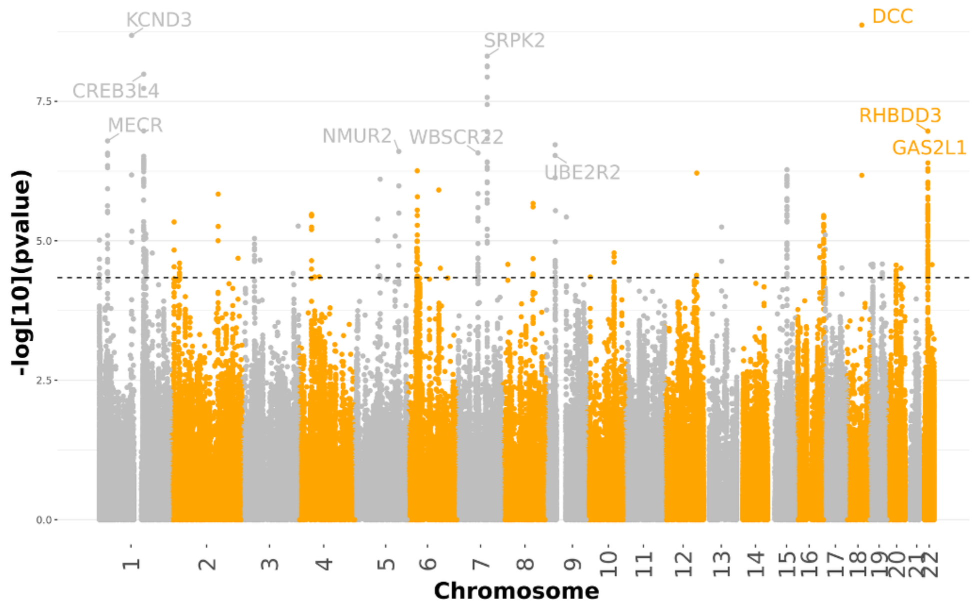


**Supplementary Figure 2. TWAS Results for Stuttering in Females Across 49 Tissues.** Chromosome is displayed on the x-axis and -log(p-value) is seen on the y-axis. The dotted line represents the FDR significance threshold (FDR p-value < 0.05). The top 10 GReX genes were labeled.

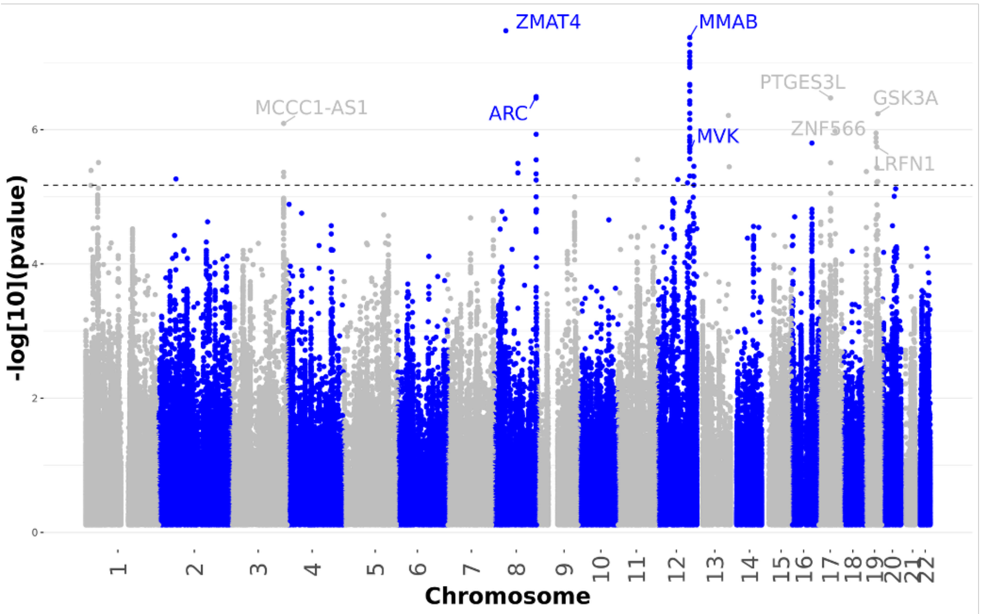


**Supplementary Figure 3. TWAS Results for Stuttering in Males Across 49 Tissues.** Chromosome is displayed on the x-axis and -log(*p*-value) is seen on the y-axis. The dotted line represents the FDR significance threshold (FDR *p*-value < 0.05). The top 10 GReX genes were labeled.

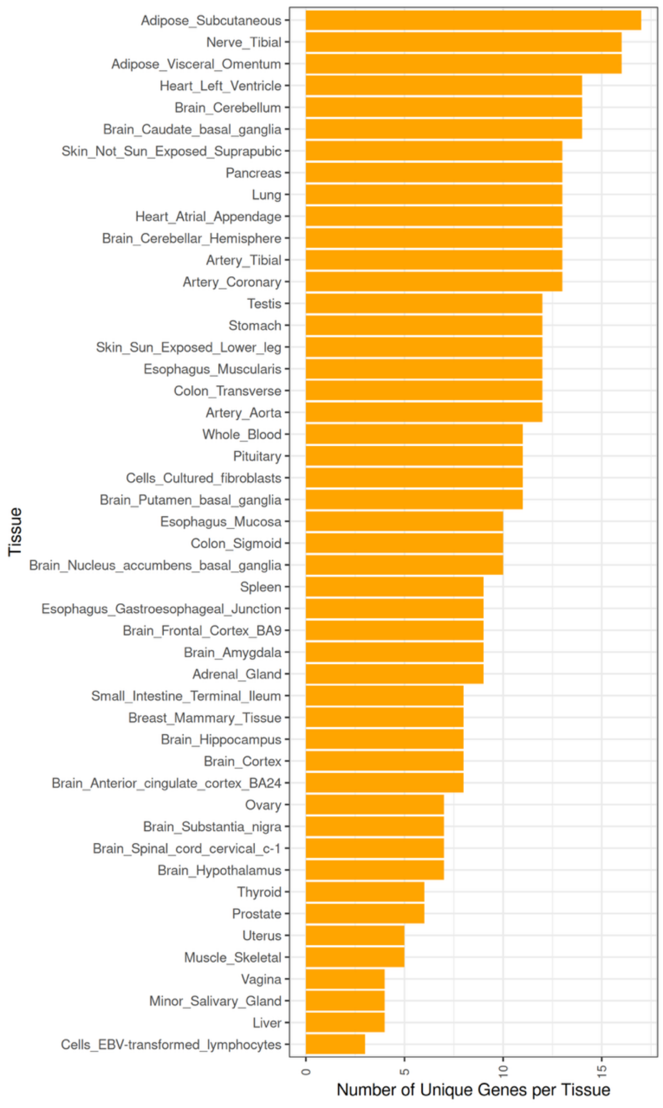


**Supplementary Figure 4. Histogram of Number of Unique GReX Genes per Tissue Female.** The number of unique GReX genes is depicted on the x-axis and tissue type is found on the y-axis. Note: the number of results that map to specific tissue will be influenced by the sample size of the eQTL discovery in GTEx[5].

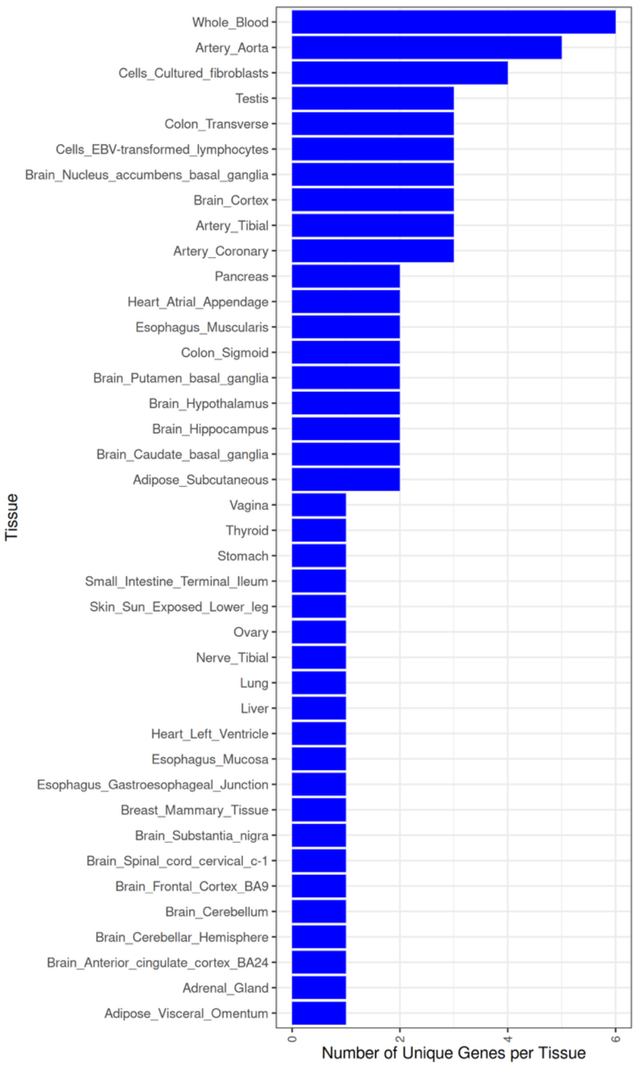


**Supplementary Figure 5. Histogram of Number of Unique GReX Genes per Tissue Male.** The number of unique GReX genes is depicted on the x-axis and tissue type is found on the y-axis. Note: the number of results that map to specific tissue will be influenced by the sample size of the eQTL discovery in GTEx[5].

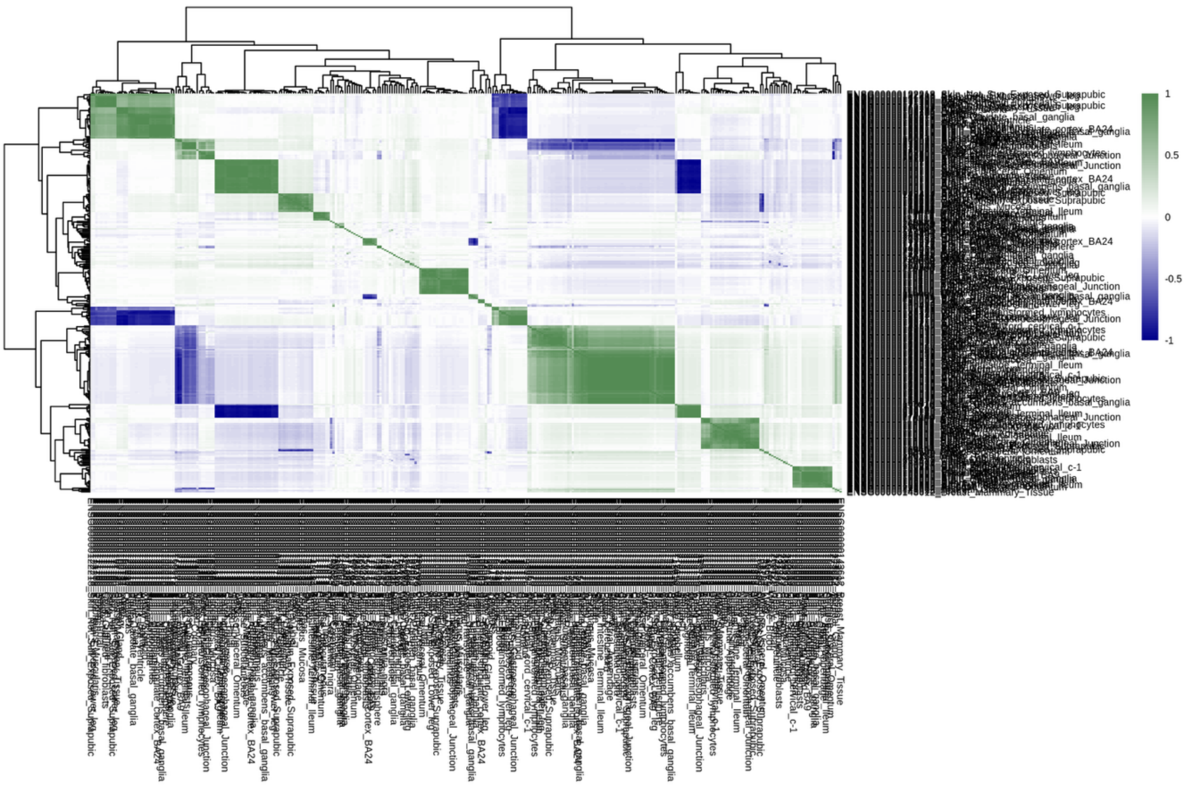


**Supplementary Figure 6. Correlation of GReX on Chromosome 1 for Sex-Combined Stuttering Gene-Tissue Pairs.**

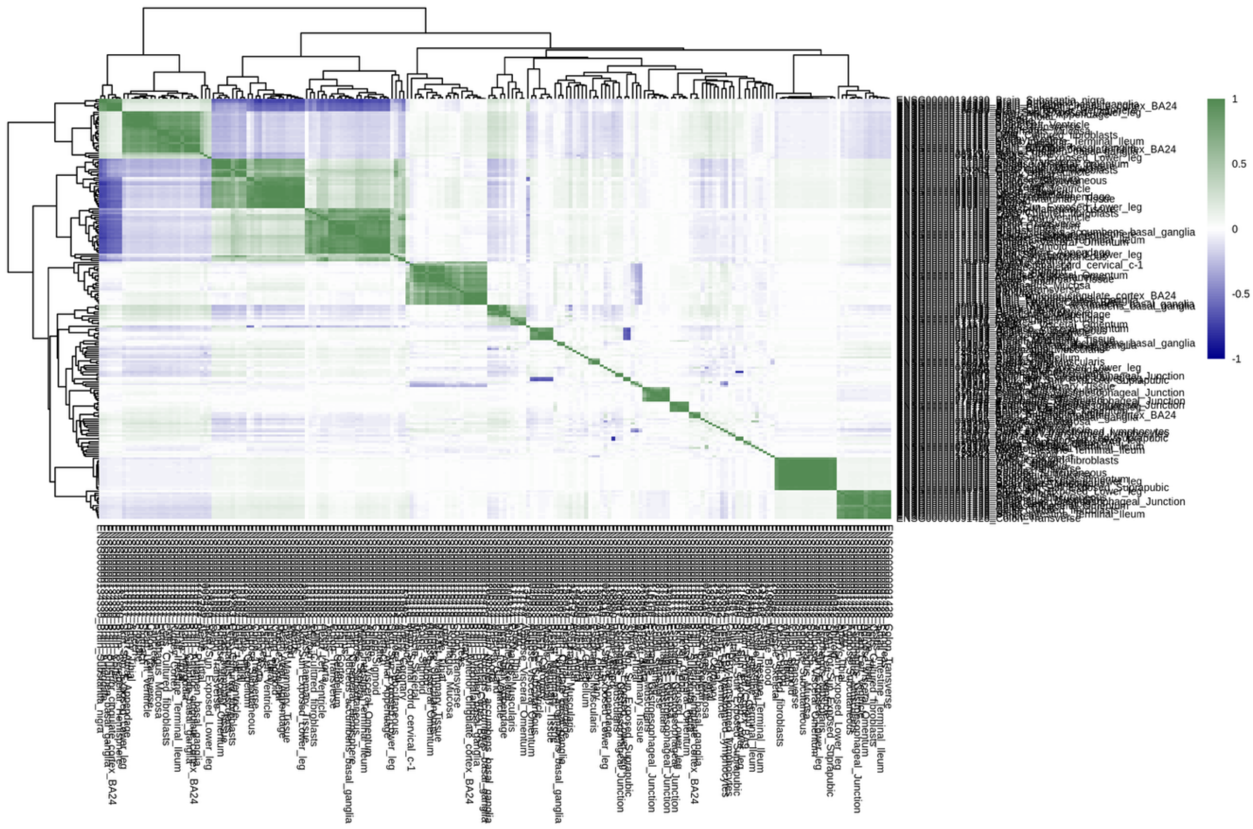

**Supplementary Figure 7. Correlation of GReX on Chromosome 2 for Sex-Combined Stuttering Gene-Tissue Pairs.**

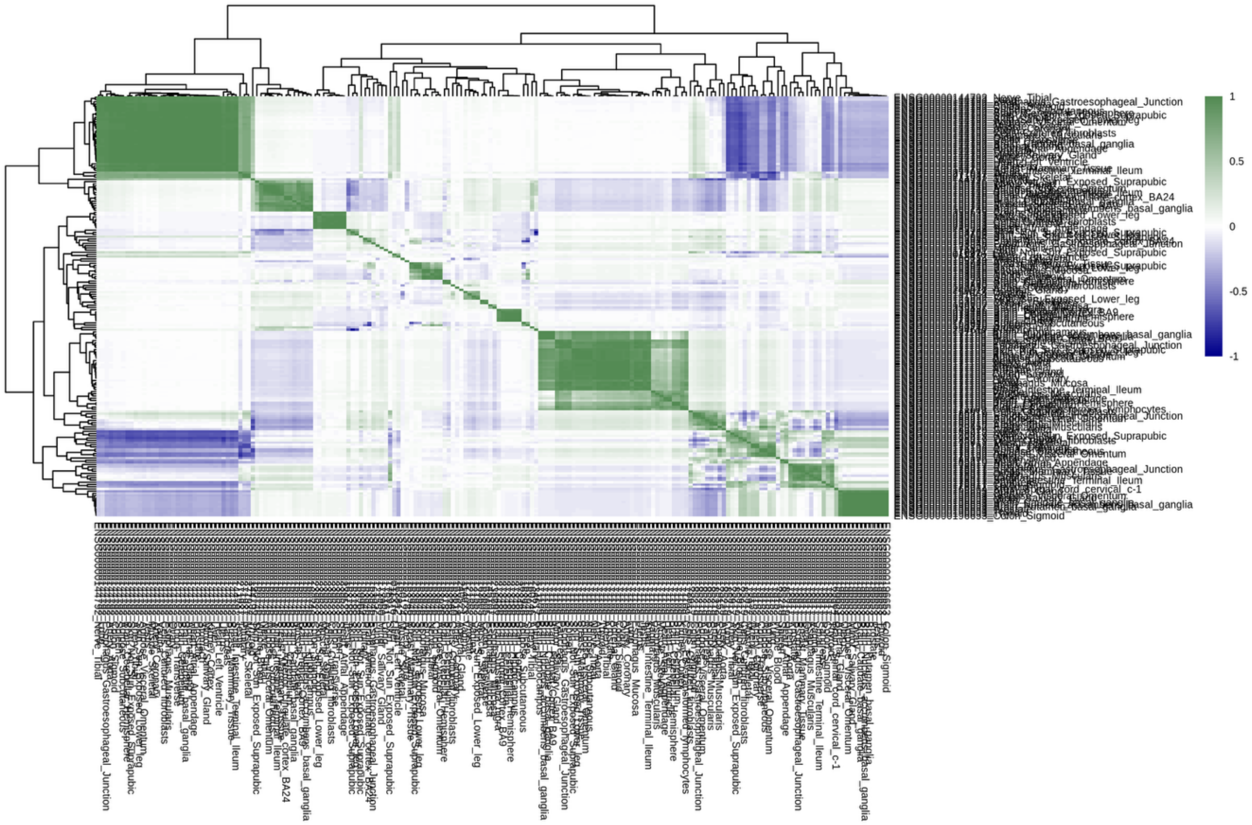


**Supplementary Figure 8. Correlation of GReX on Chromosome 3 for Sex-Combined Stuttering Gene-Tissue Pairs.**

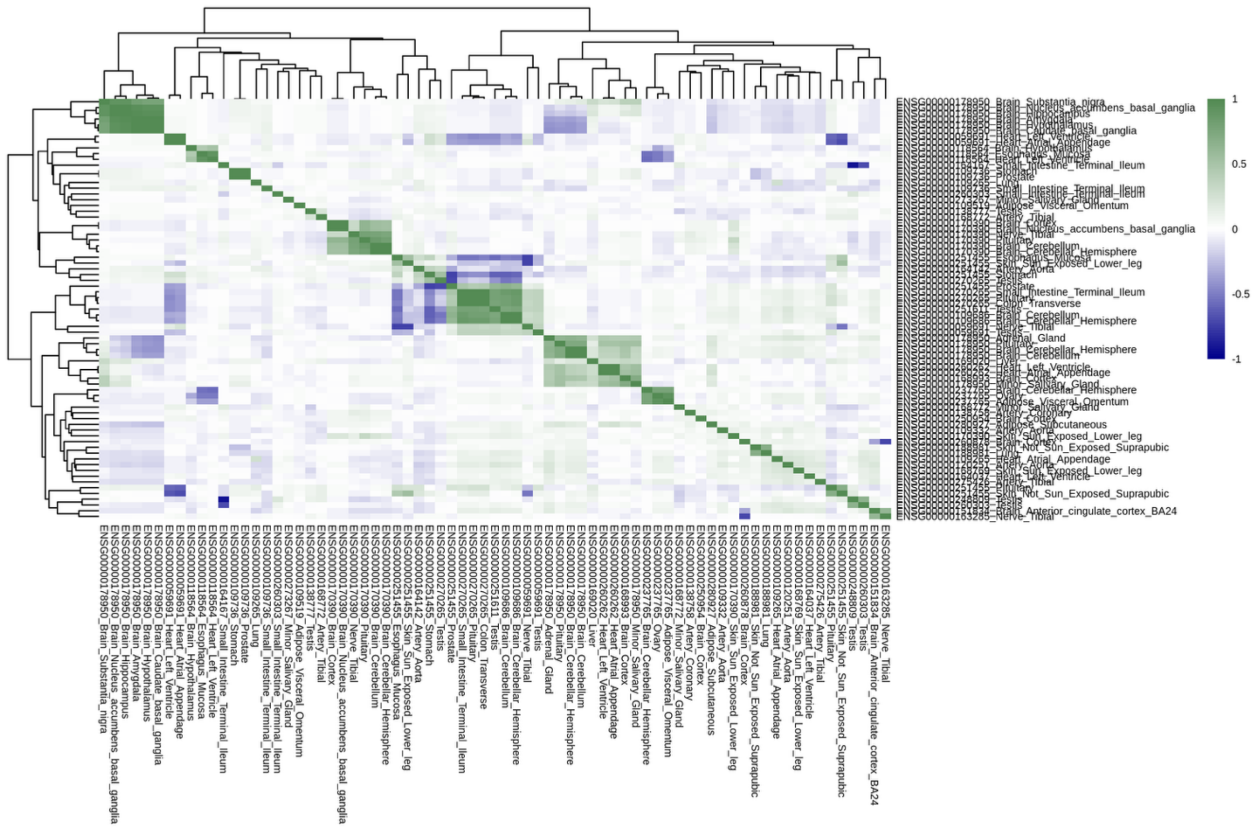

**Supplementary Figure 9. Correlation of GReX on Chromosome 4 for Sex-Combined Stuttering Gene-Tissue Pairs.**

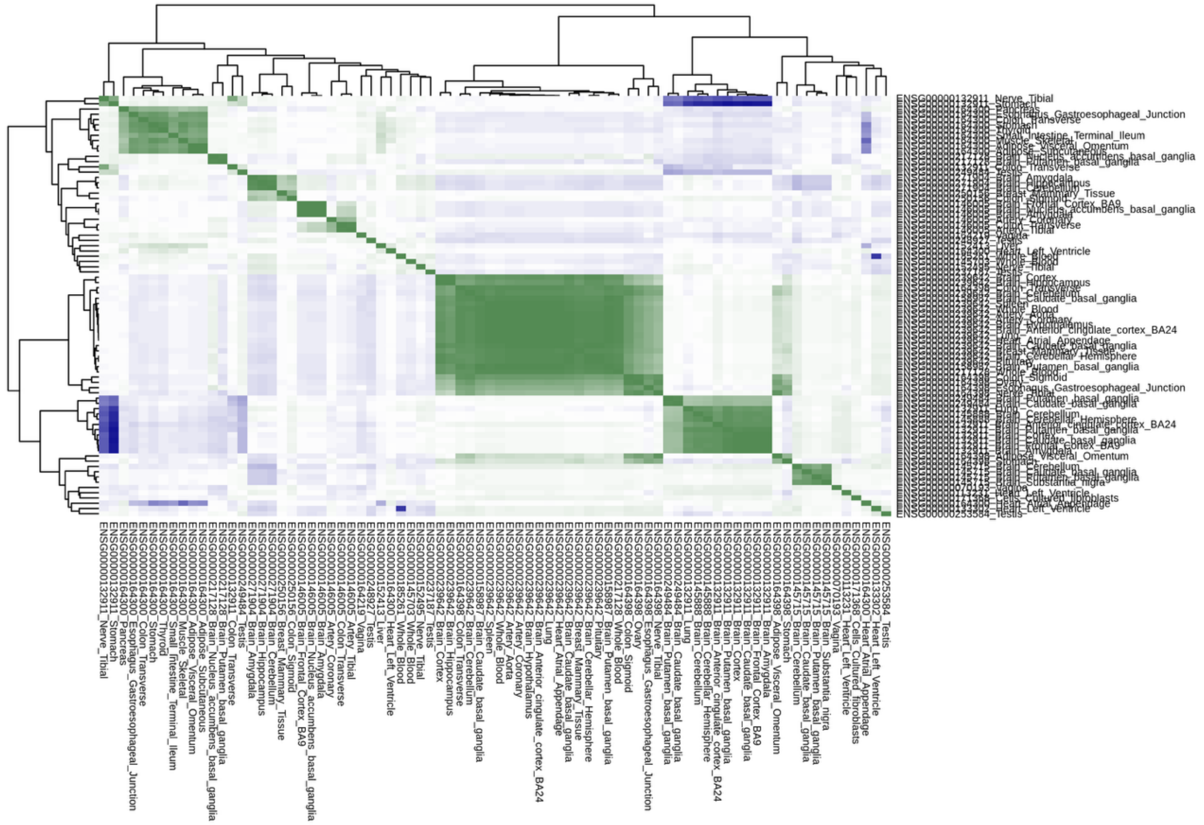


**Supplementary Figure 10. Correlation of GReX on Chromosome 5 for Sex-Combined Stuttering Gene-Tissue Pairs.**

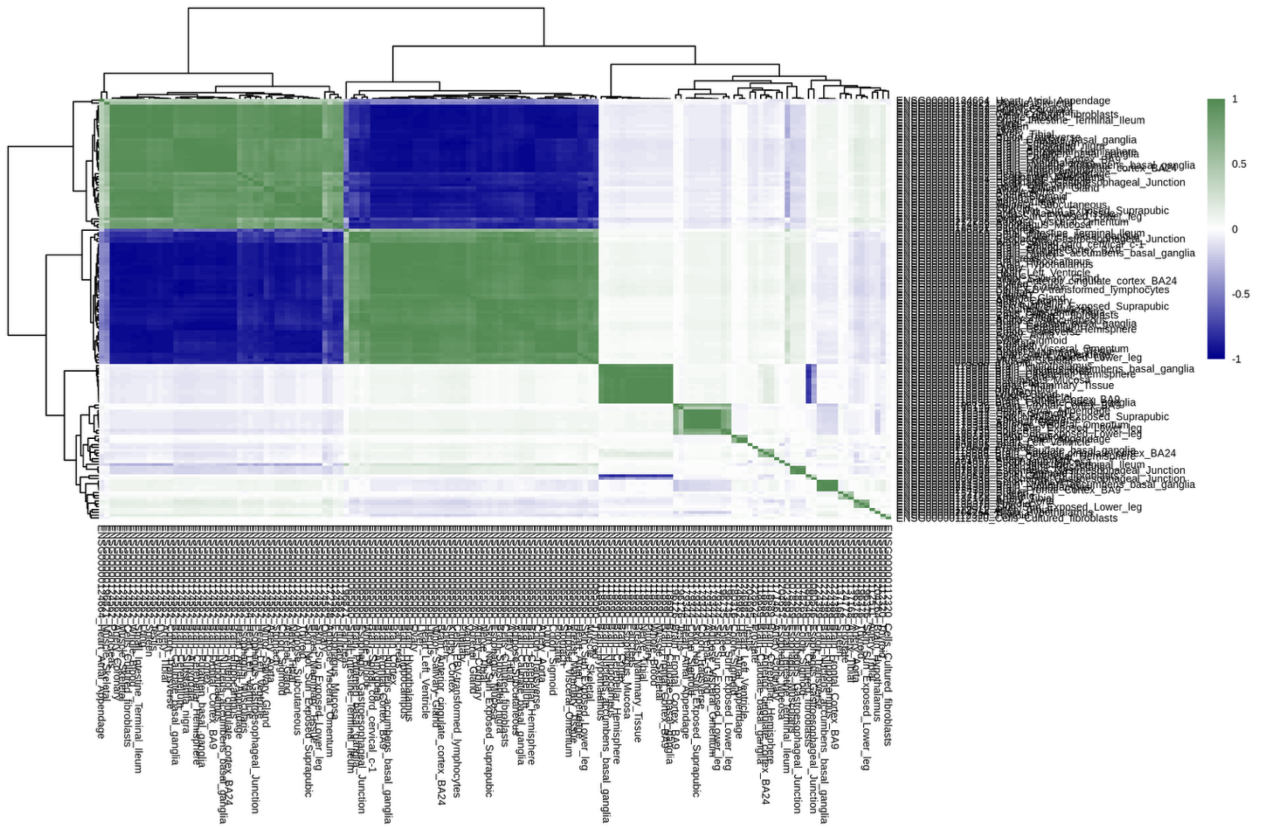


**Supplementary Figure 11. Correlation of GReX on Chromosome 6 for Sex-Combined Stuttering Gene-Tissue Pairs.**

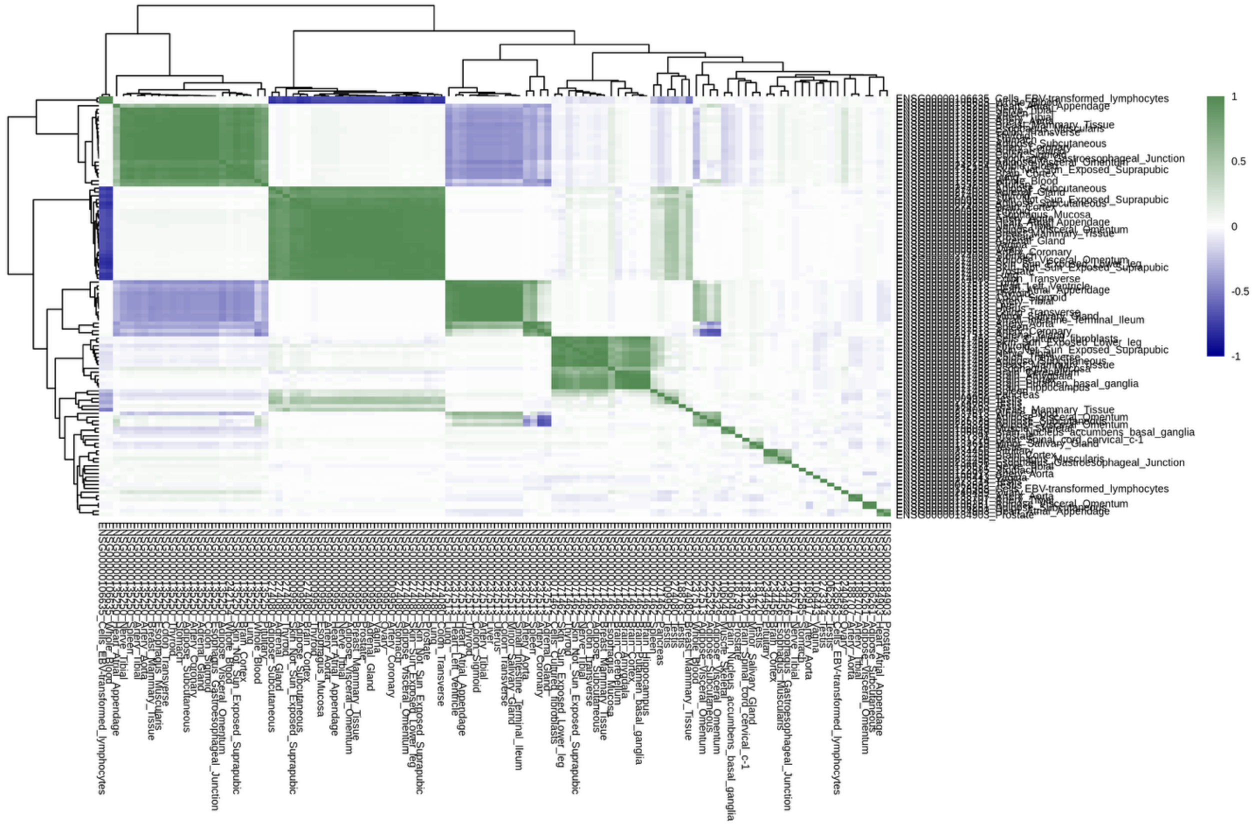


**Supplementary Figure 12. Correlation of GReX on Chromosome 7 for Sex-Combined Stuttering Gene-Tissue Pairs.**

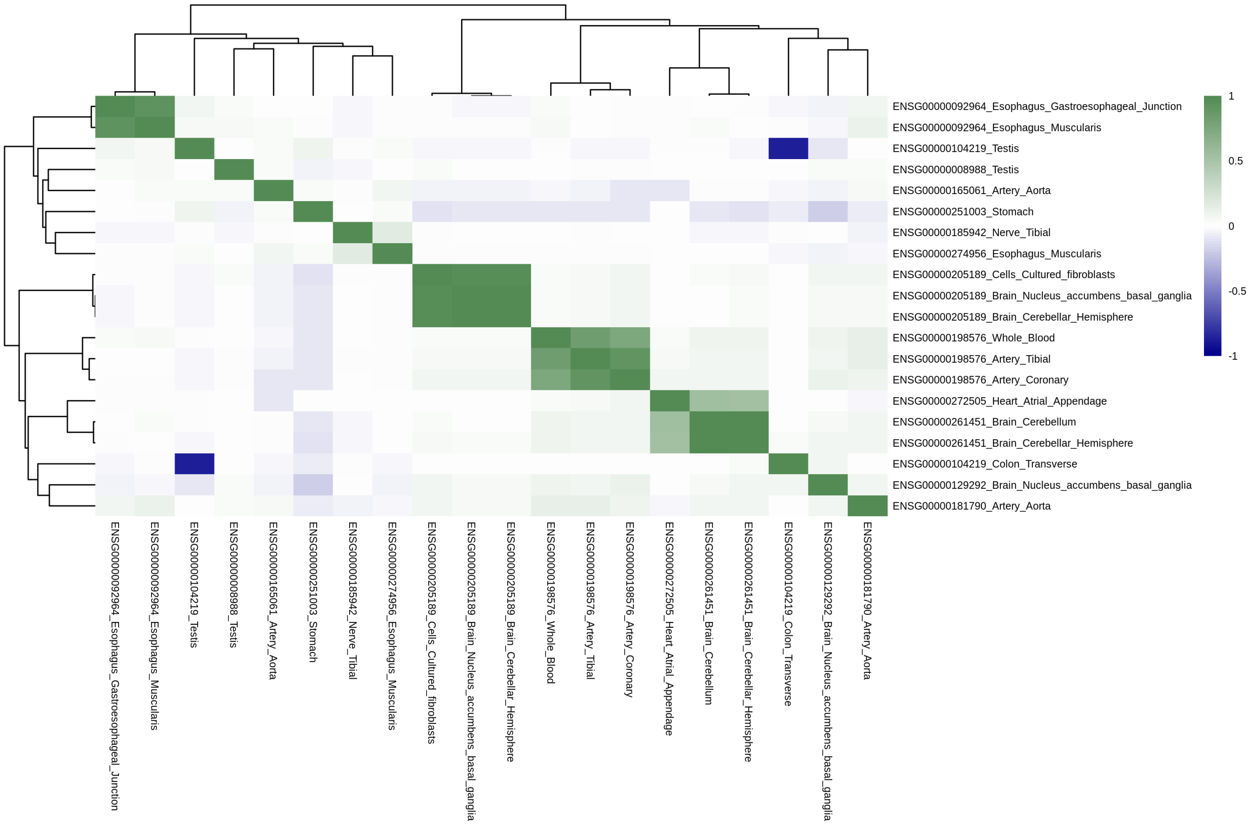


**Supplementary Figure 13. Correlation of GReX on Chromosome 8 for Sex-Combined Stuttering Gene-Tissue Pairs.**

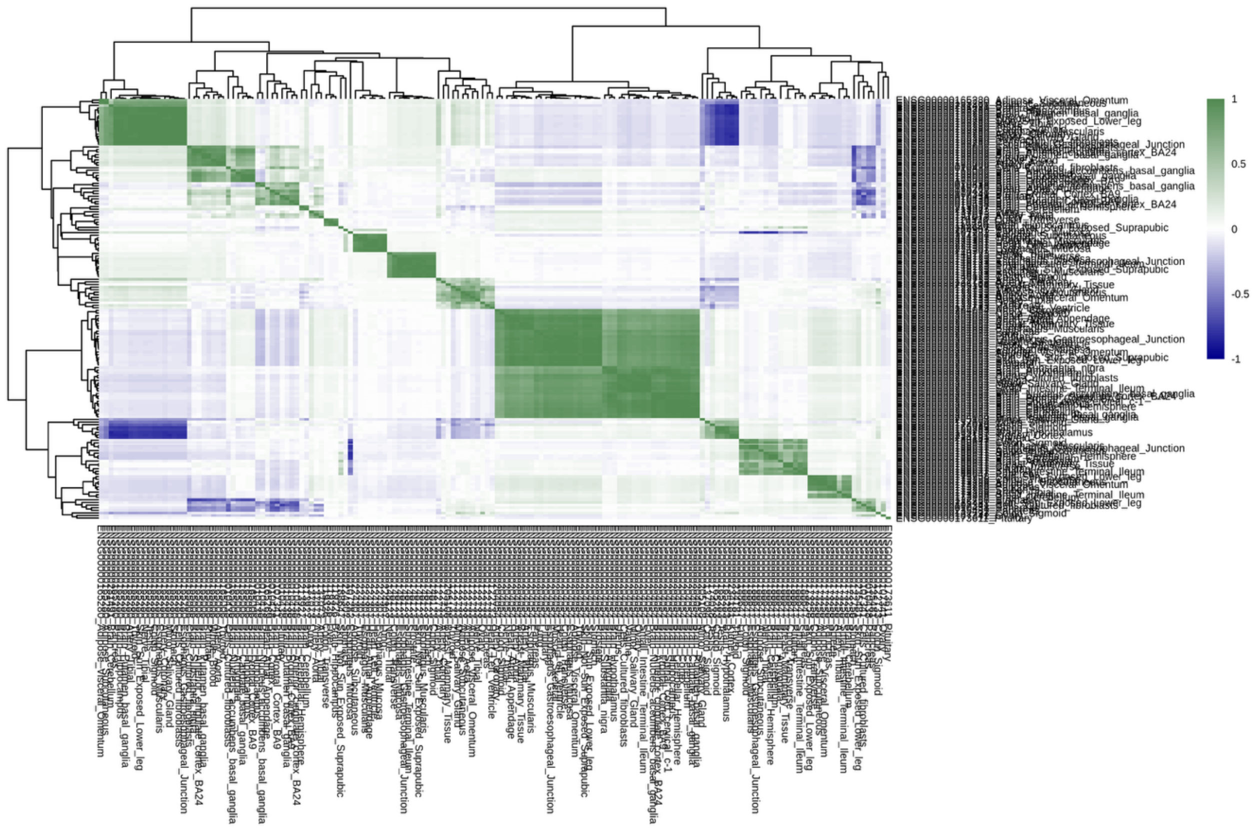


**Supplementary Figure 14. Correlation of GReX on Chromosome 9 for Sex-Combined Stuttering Gene-Tissue Pairs.**

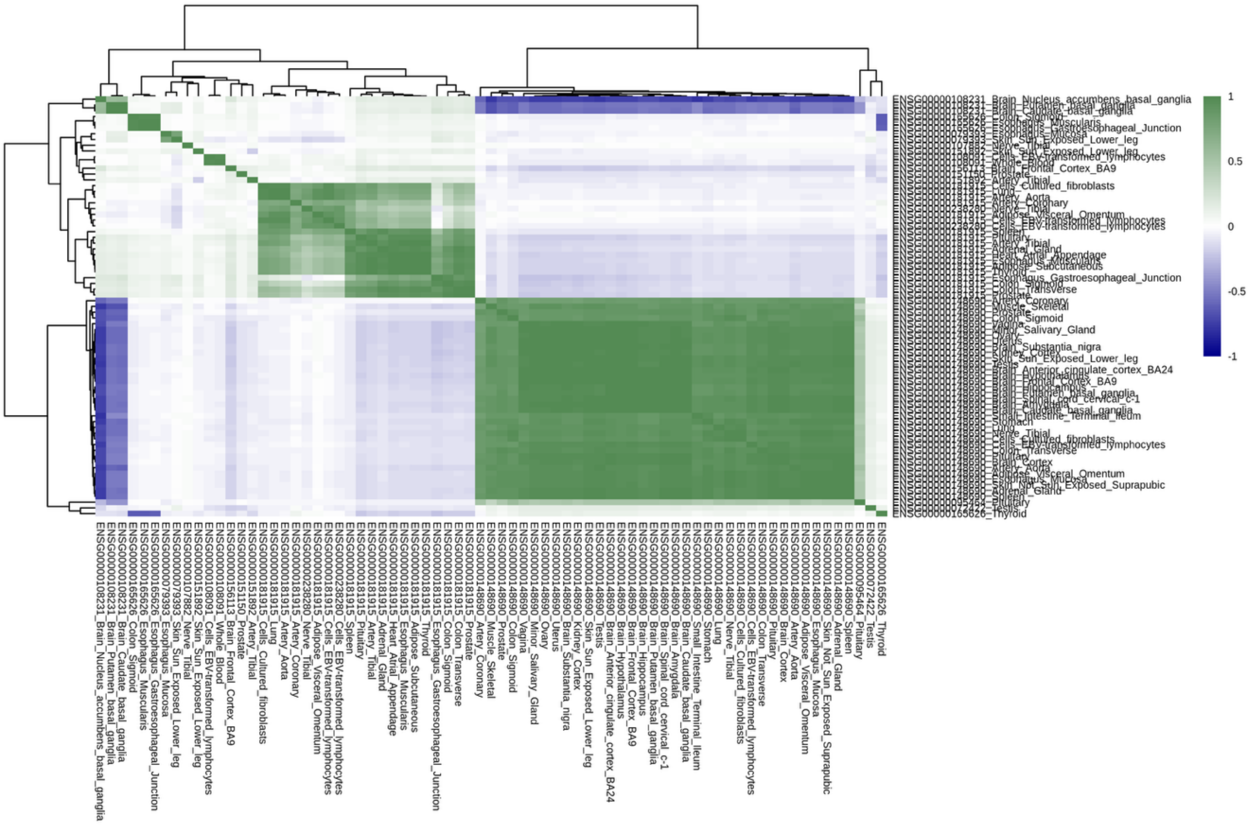


**Supplementary Figure 15. Correlation of GReX on Chromosome 10 for Sex-Combined Stuttering Gene-Tissue Pairs.**

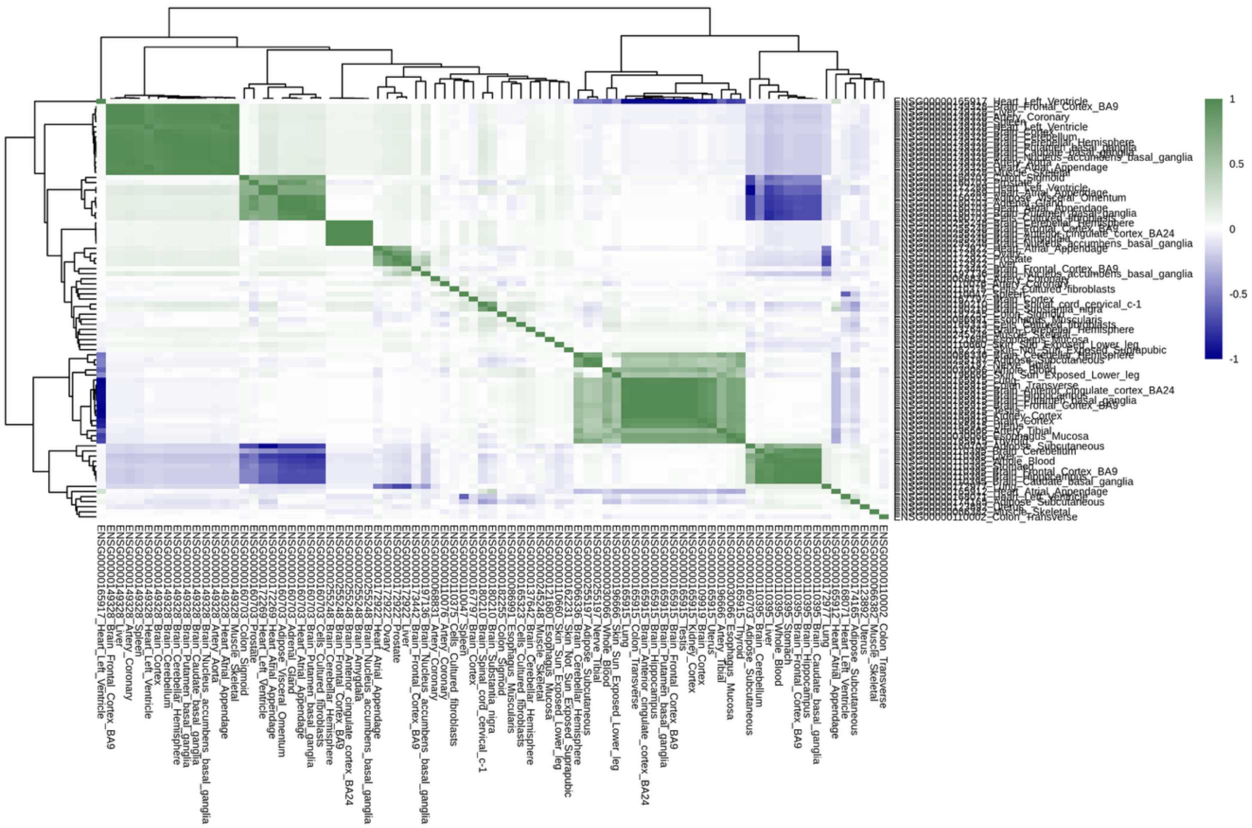


**Supplementary Figure 16. Correlation of GReX on Chromosome 11 for Sex-Combined Stuttering Gene-Tissue Pairs.**

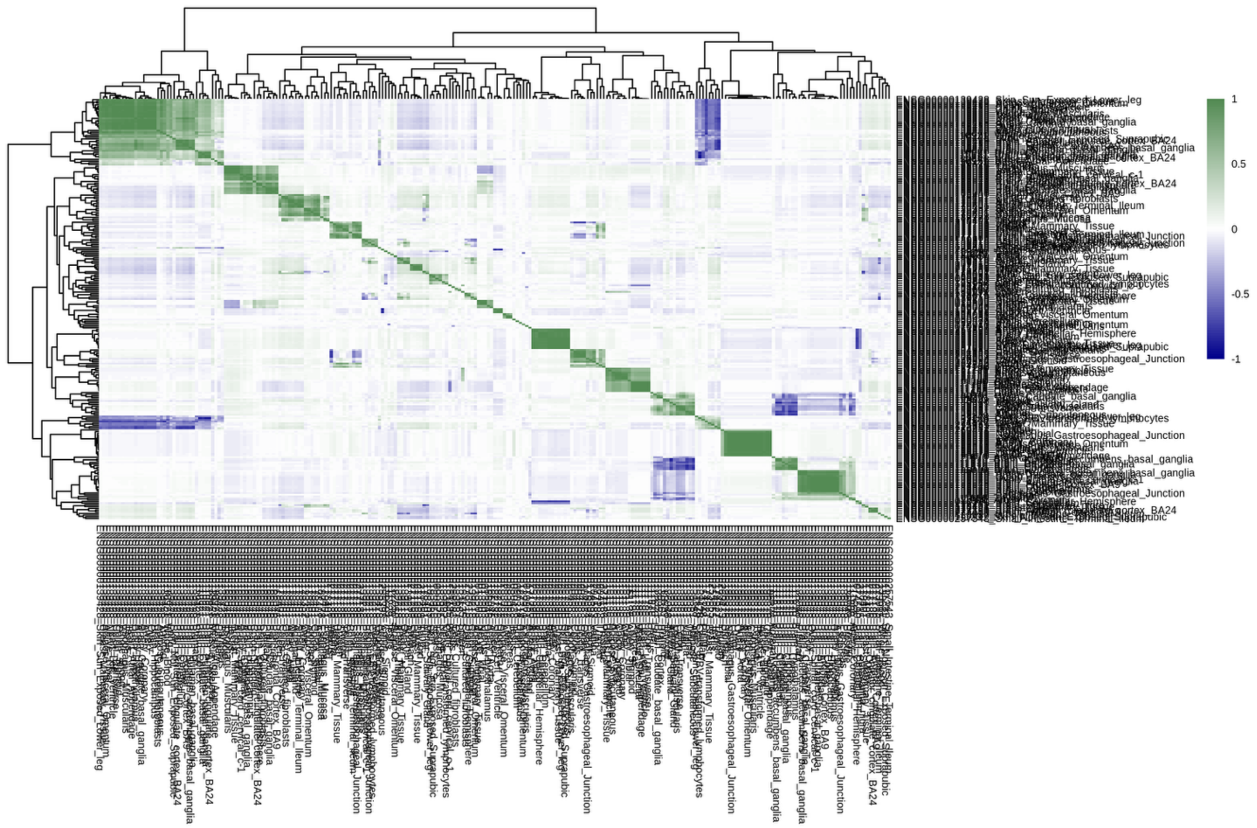

**Supplementary Figure 17. Correlation of GReX on Chromosome 12 for Sex-Combined Stuttering Gene-Tissue Pairs.**

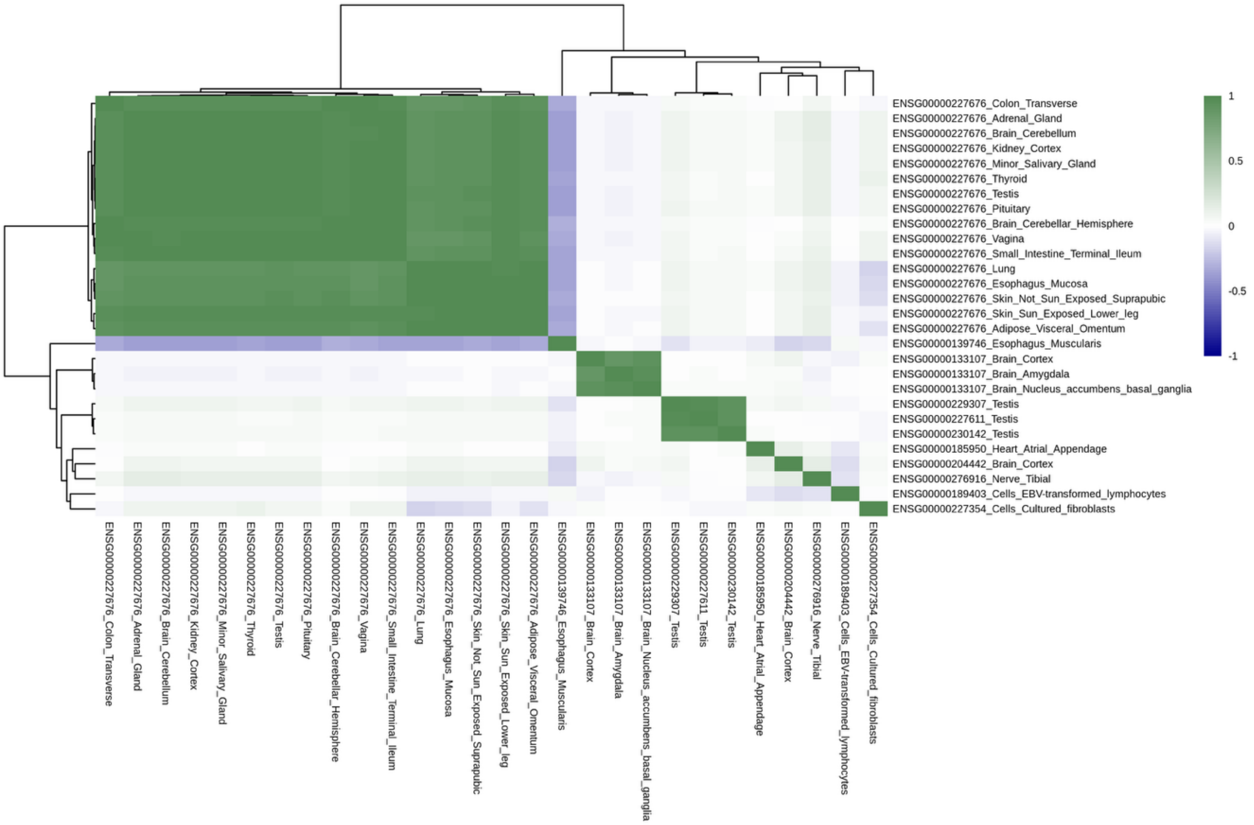


**Supplementary Figure 18. Correlation of GReX on Chromosome 13 for Sex-Combined Stuttering Gene-Tissue Pairs.**

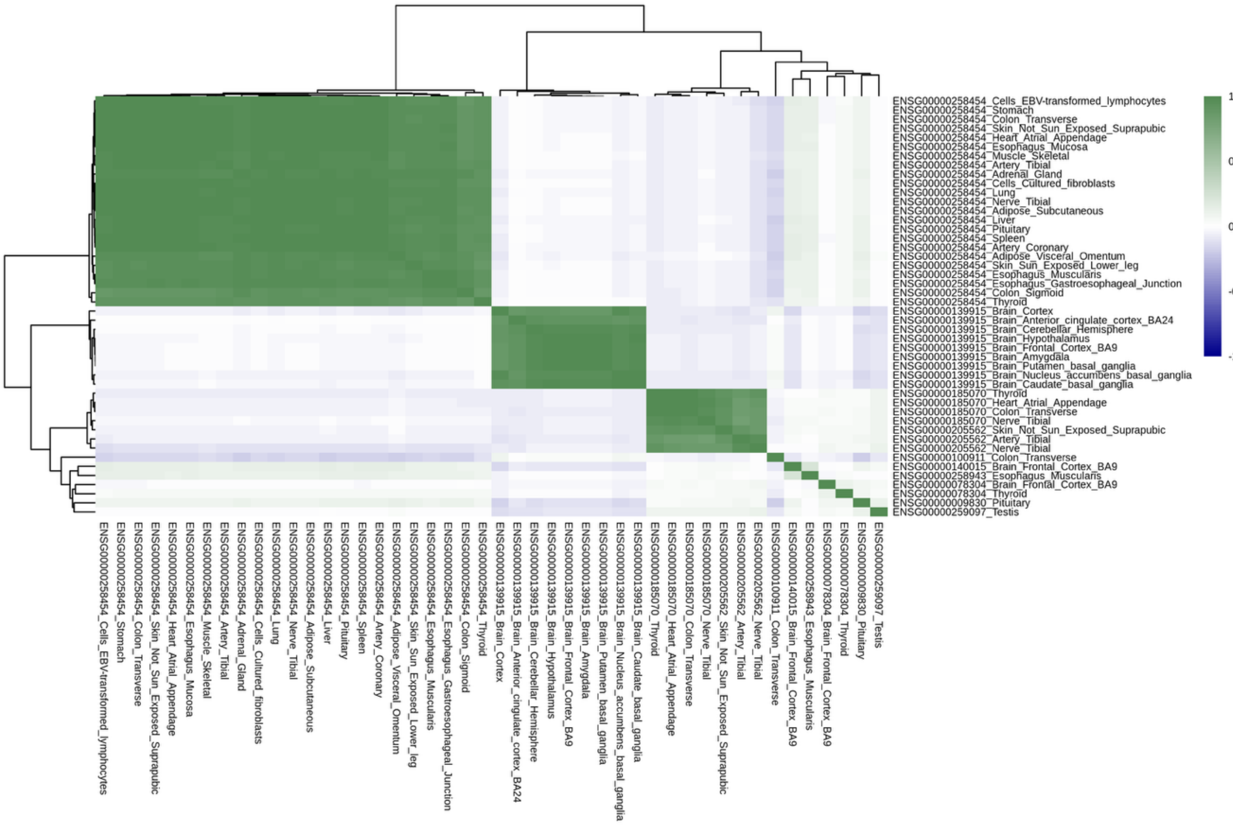


**Supplementary Figure 19. Correlation of GReX on Chromosome 14 for Sex-Combined Stuttering Gene-Tissue Pairs.**

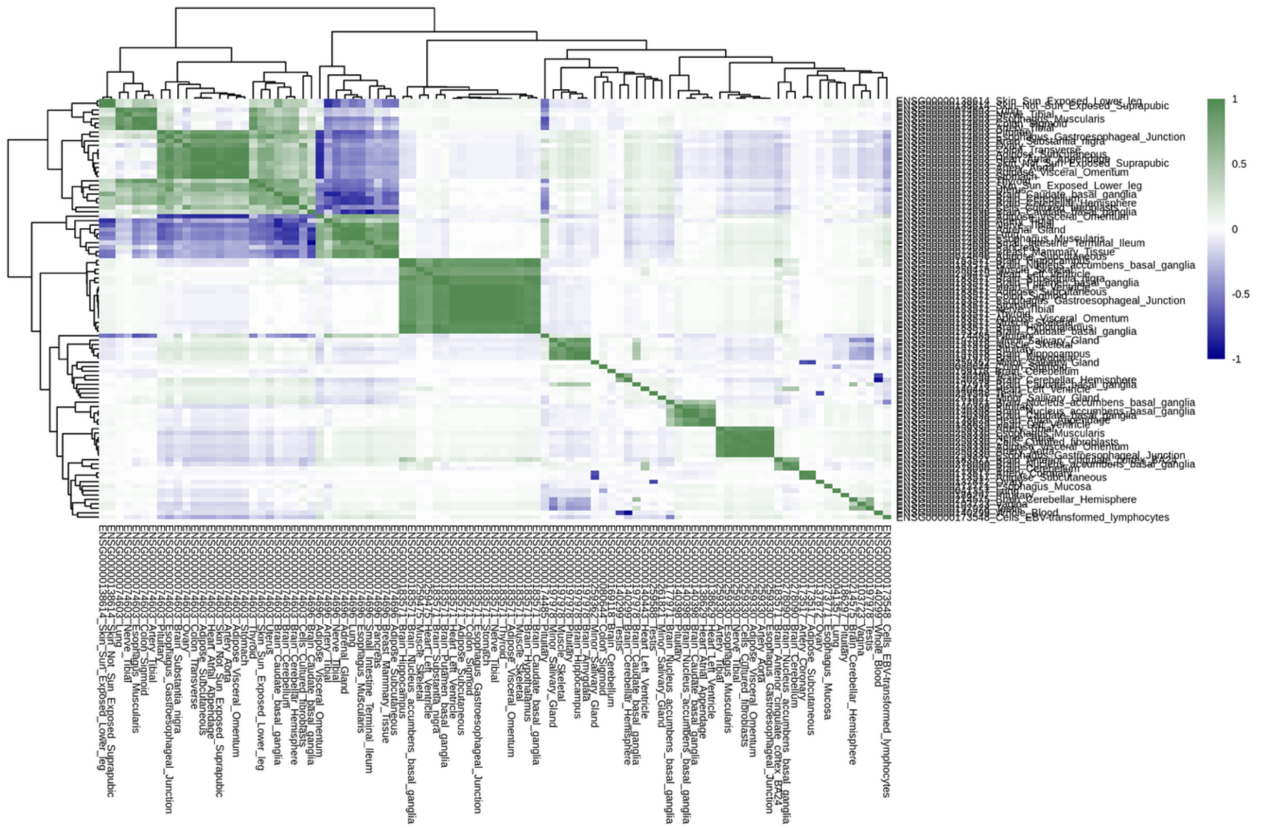


**Supplementary Figure 20. Correlation of GReX on Chromosome 15 for Sex-Combined Stuttering Gene-Tissue Pairs.**

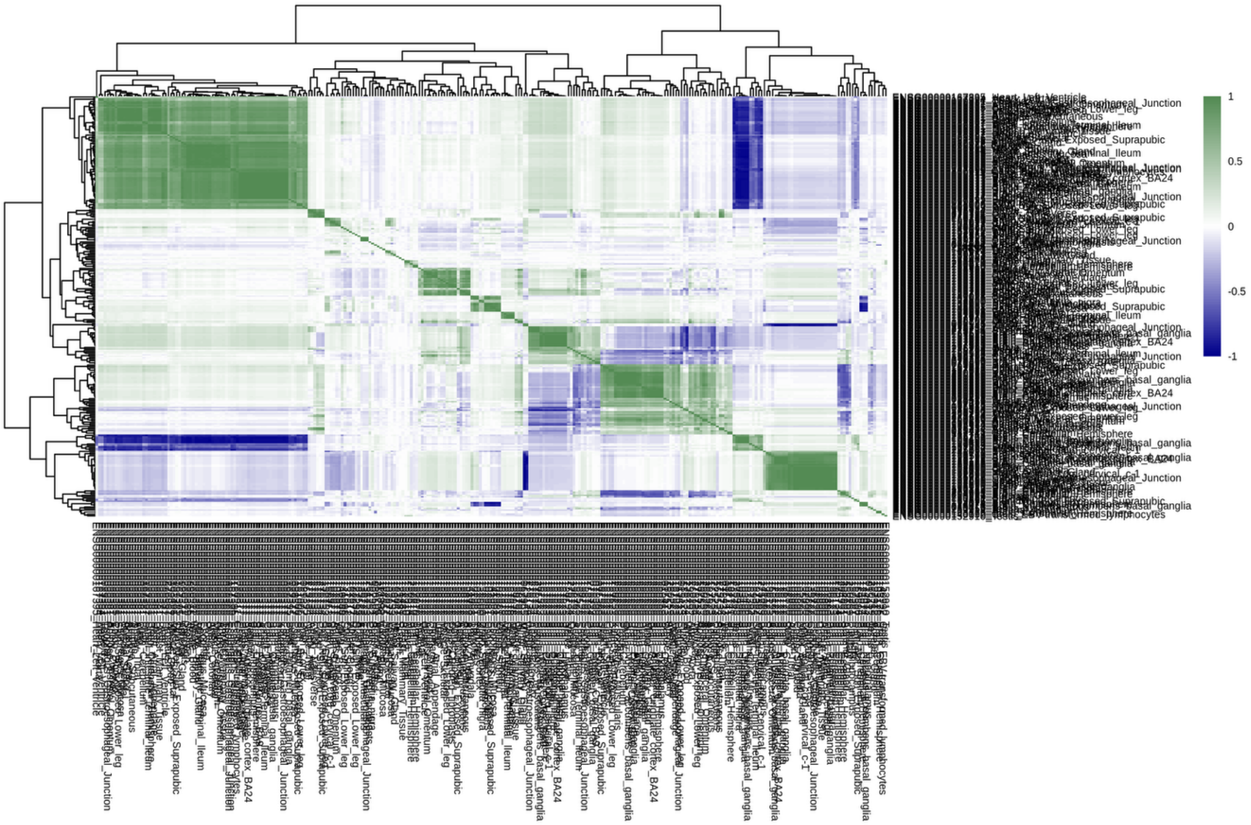

**Supplementary Figure 21. Correlation of GReX on Chromosome 16 for Sex-Combined Stuttering Gene-Tissue Pairs.**

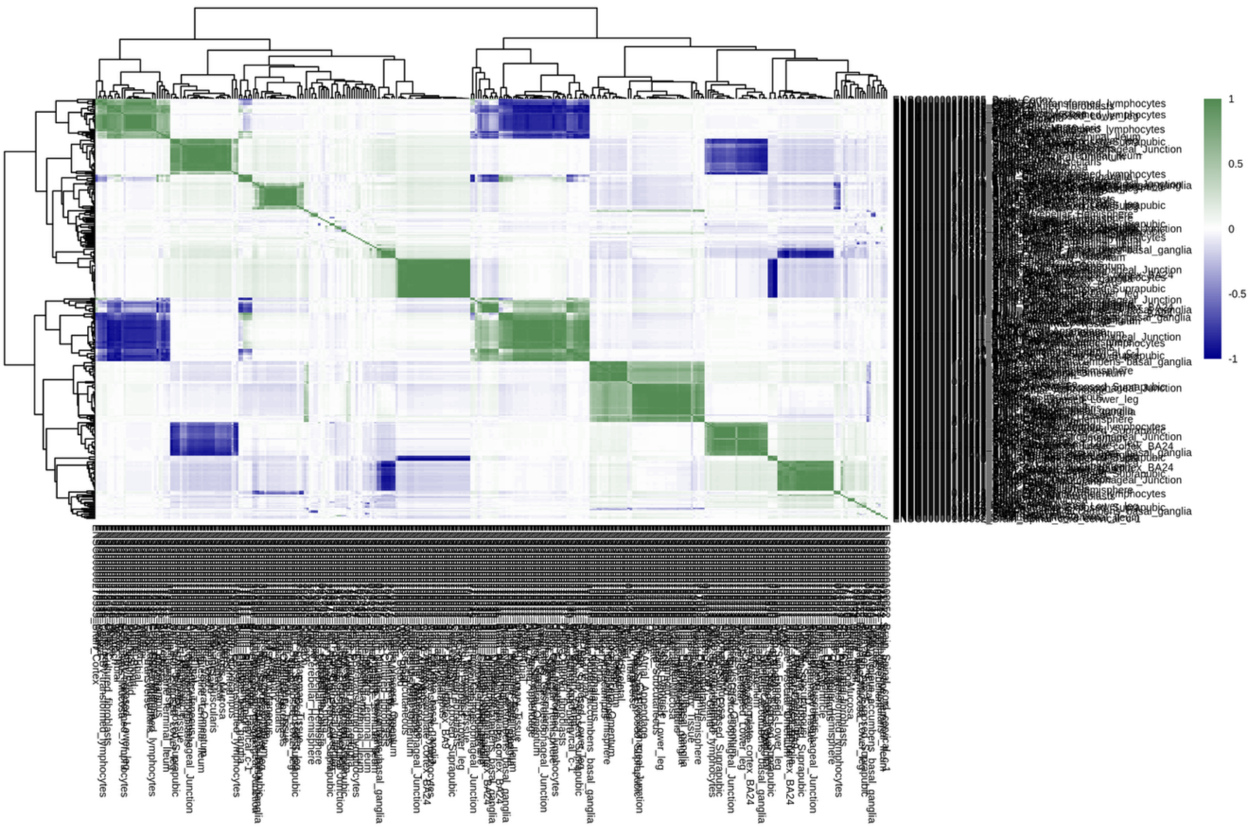


**Supplementary Figure 22. Correlation of GReX on Chromosome 17 for Sex-Combined Stuttering Gene-Tissue Pairs.**

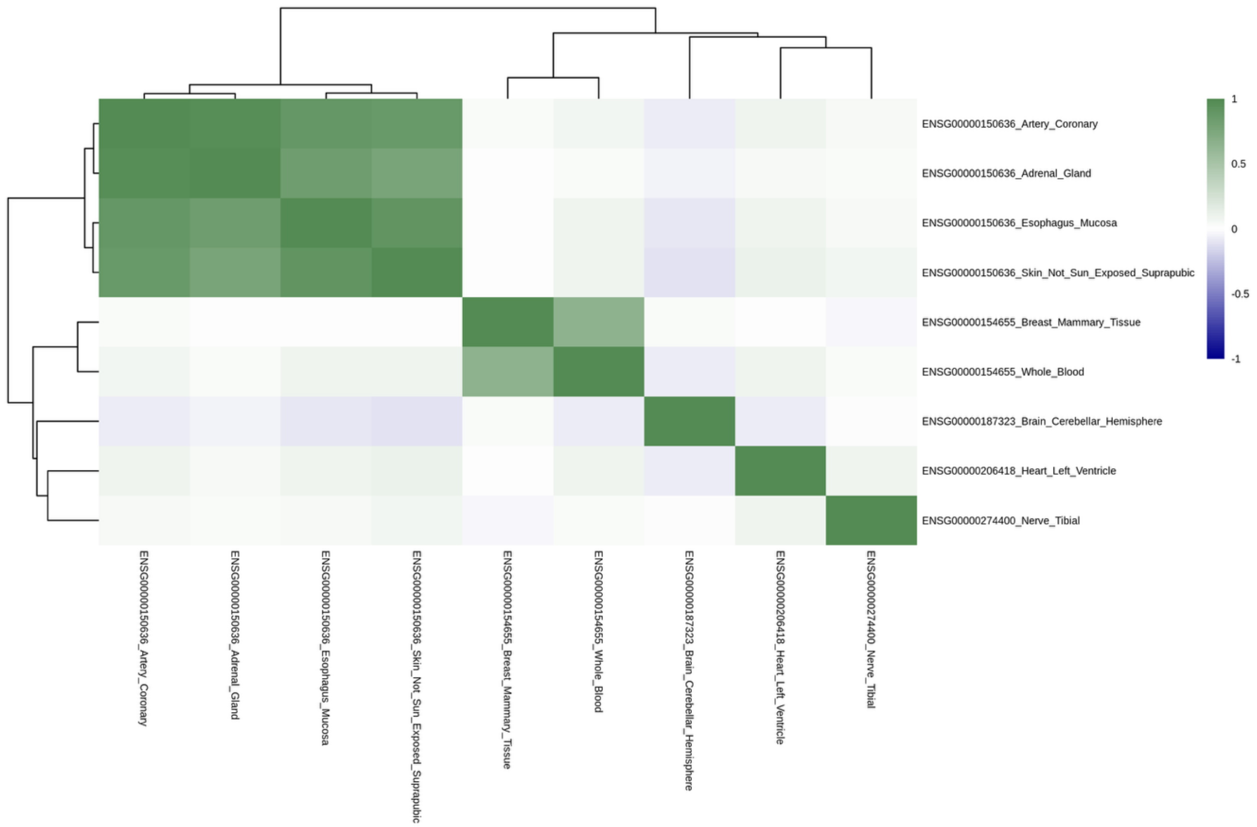


**Supplementary Figure 23. Correlation of GReX on Chromosome 18 for Sex-Combined Stuttering Gene-Tissue Pairs.**

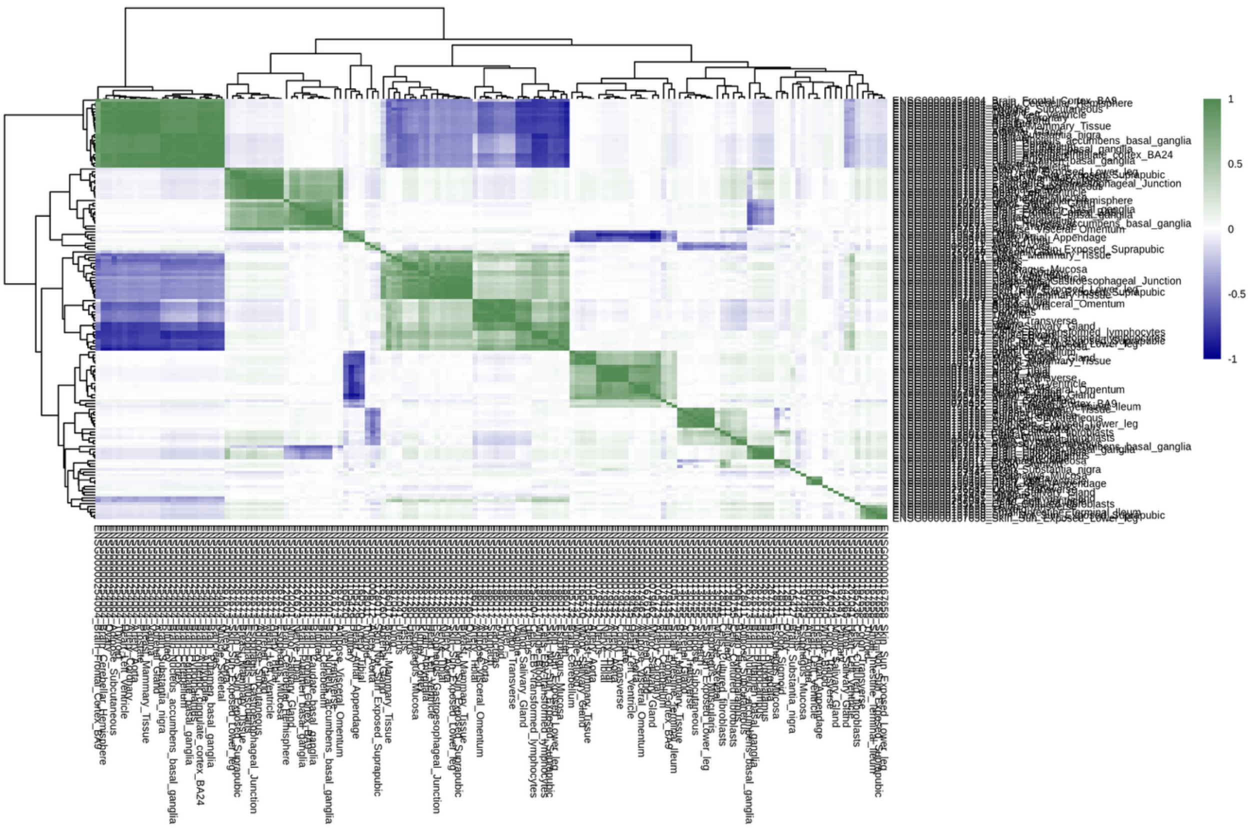


**Supplementary Figure 24. Correlation of GReX on Chromosome 19 for Sex-Combined Stuttering Gene-Tissue Pairs.**

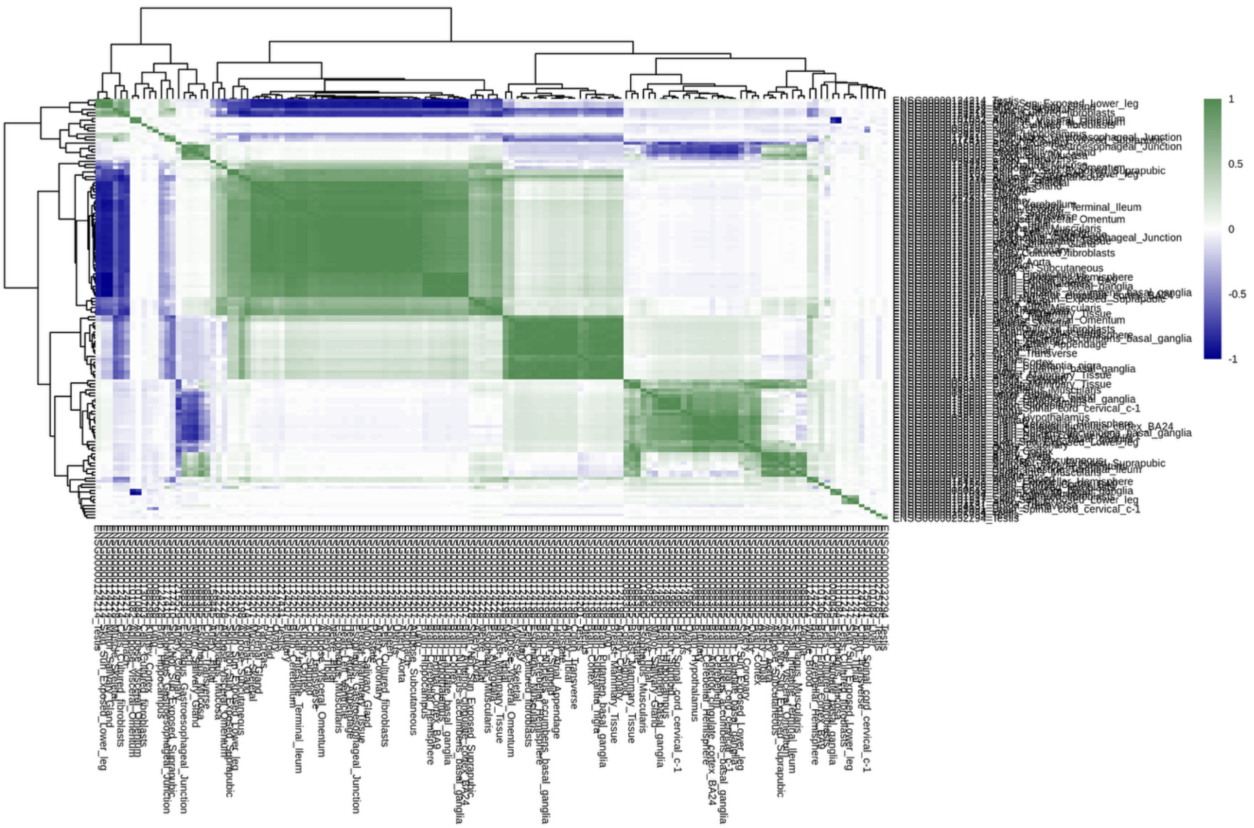


**Supplementary Figure 25. Correlation of GReX on Chromosome 20 for Sex-Combined Stuttering Gene-Tissue Pairs.**

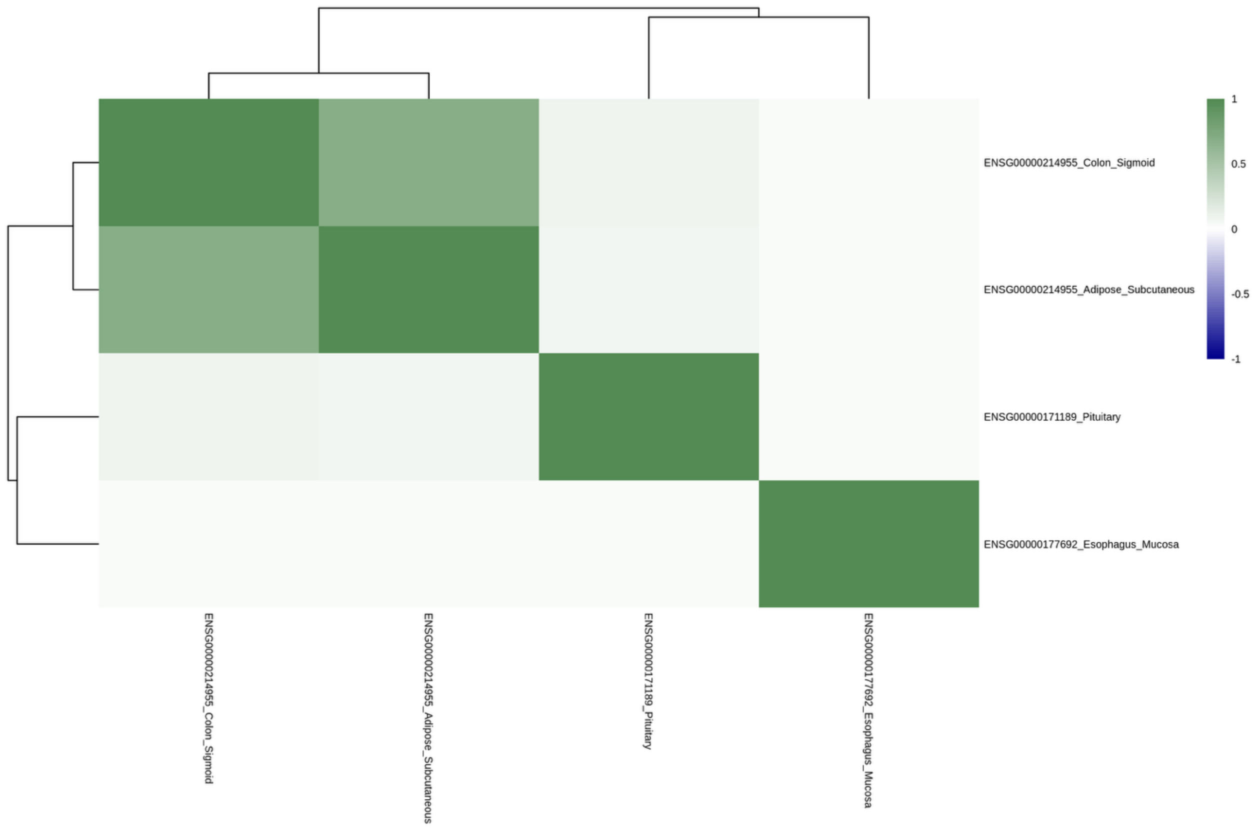


**Supplementary Figure 26. Correlation of GReX on Chromosome 21 for Sex-Combined Stuttering Gene-Tissue Pairs.**

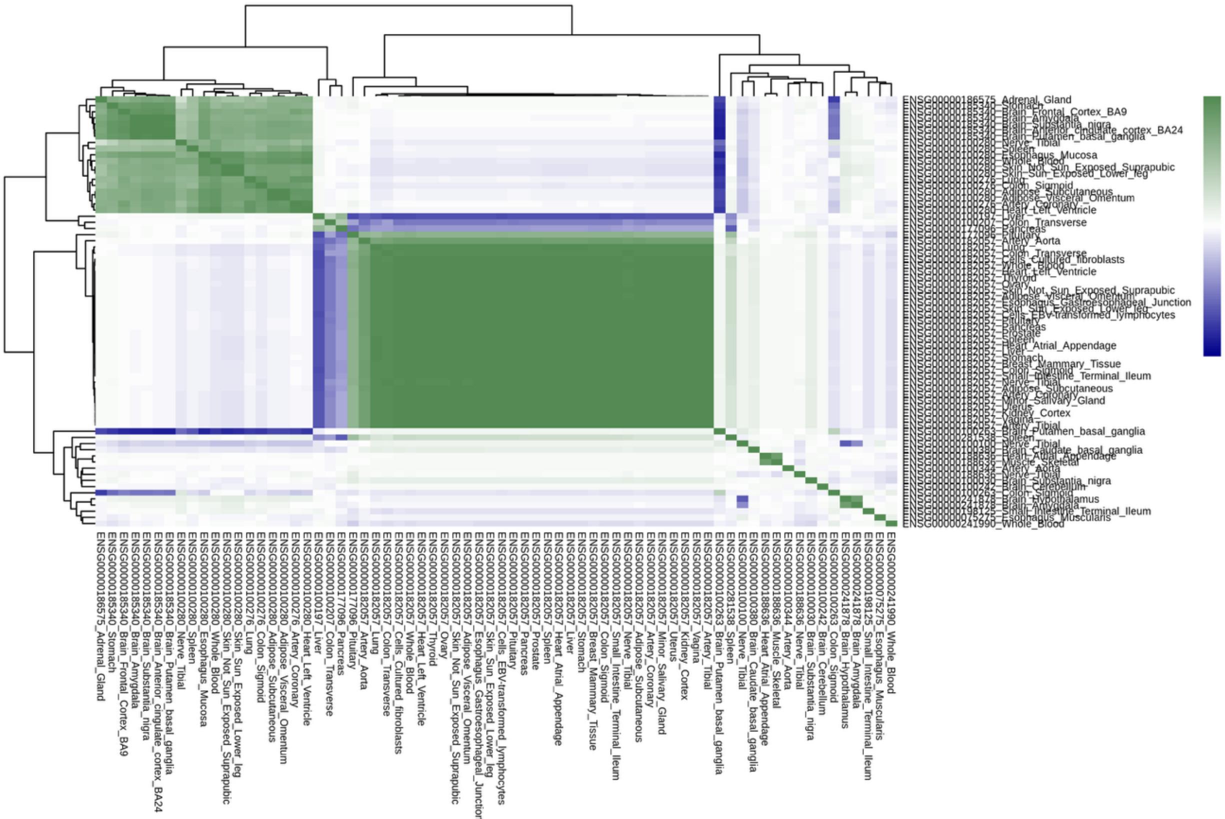


**Supplementary Figure 27. Correlation of GReX on Chromosome 22 for Sex-Combined Stuttering Gene-Tissue Pairs.**

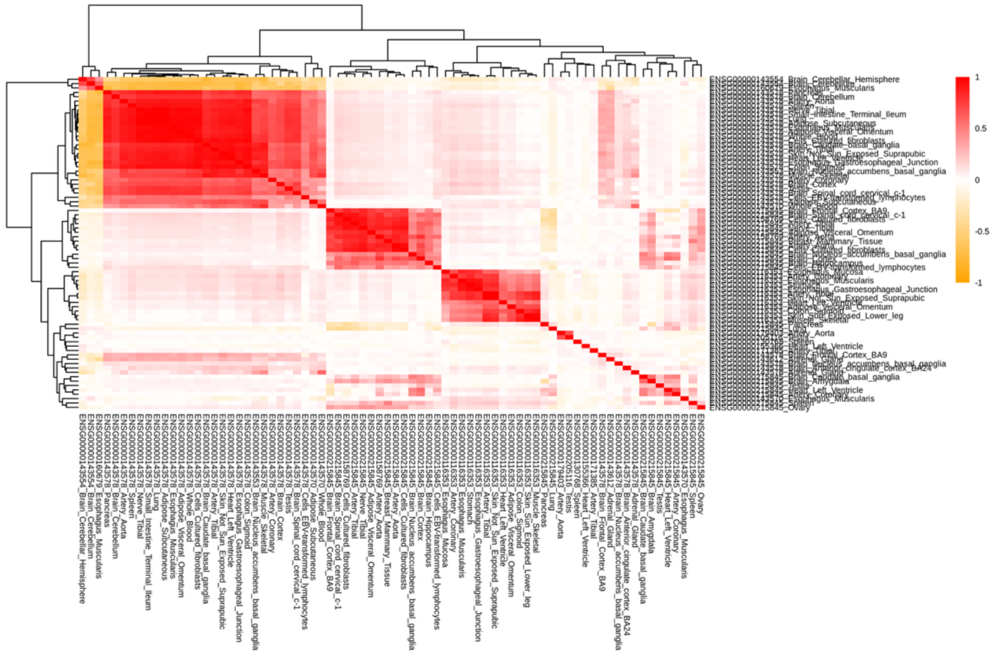


**Supplementary Figure 28. Correlation of GReX on Chromosome 1 for Stuttering in Females Gene-Tissue Pairs.**

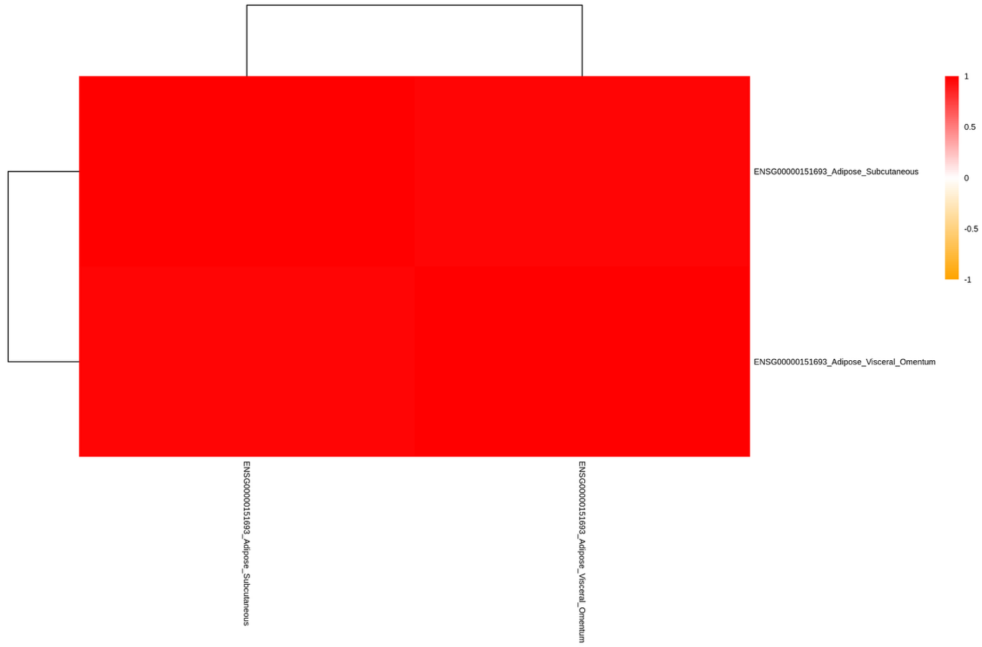


**Supplementary Figure 29. Correlation of GReX on Chromosome 2 for Stuttering in Females Gene-Tissue Pairs.**

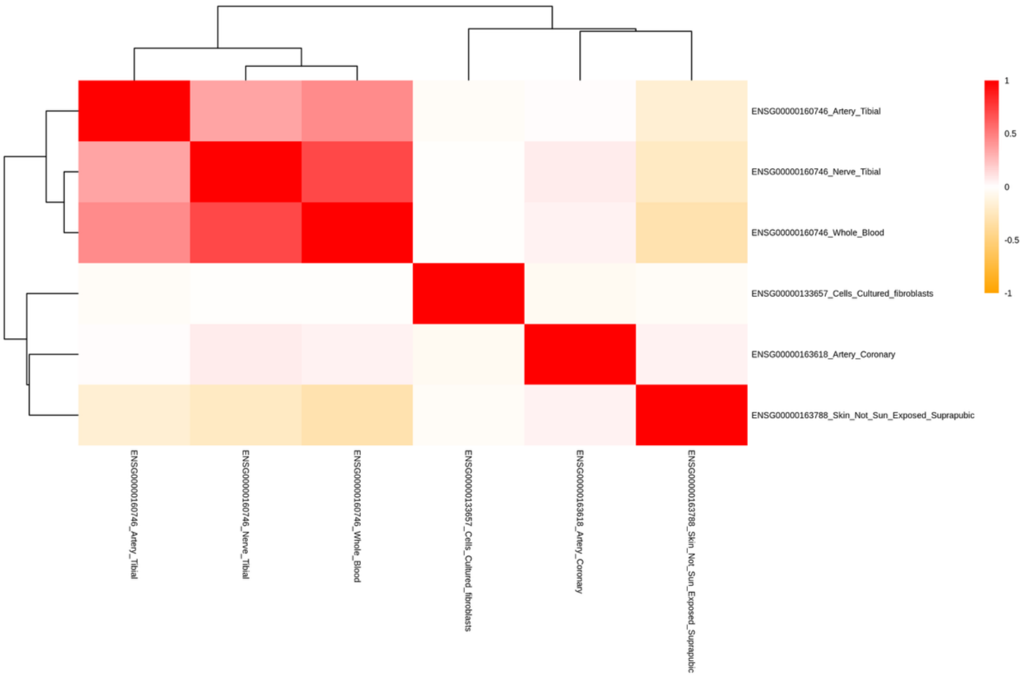


**Supplementary Figure 30. Correlation of GReX on Chromosome 3 for Stuttering in Females Gene-Tissue Pairs.**

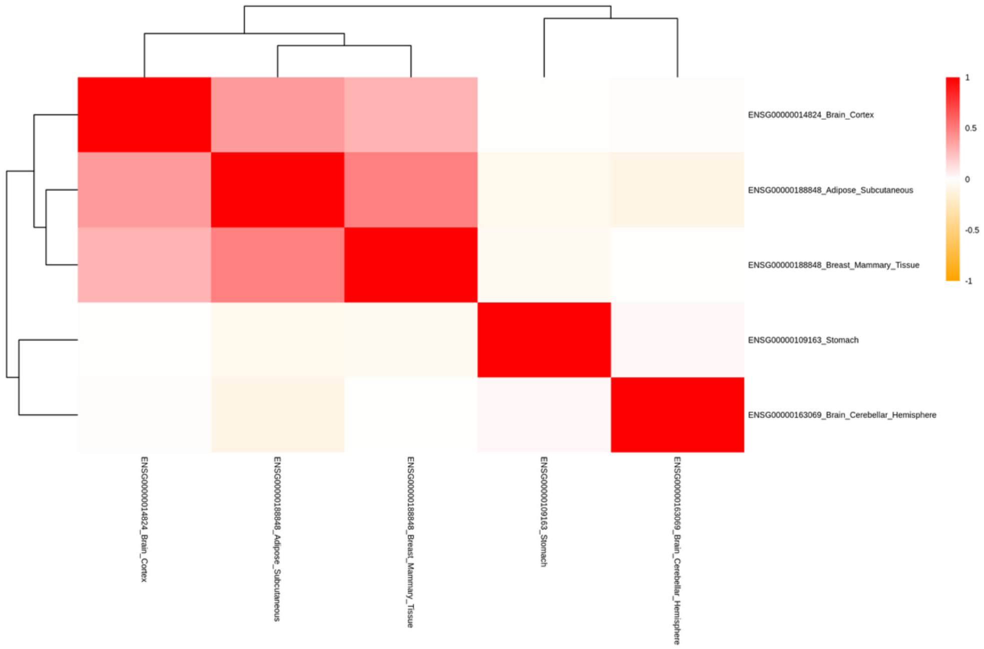


**Supplementary Figure 31. Correlation of GReX on Chromosome 4 for Stuttering in Females Gene-Tissue Pairs.**

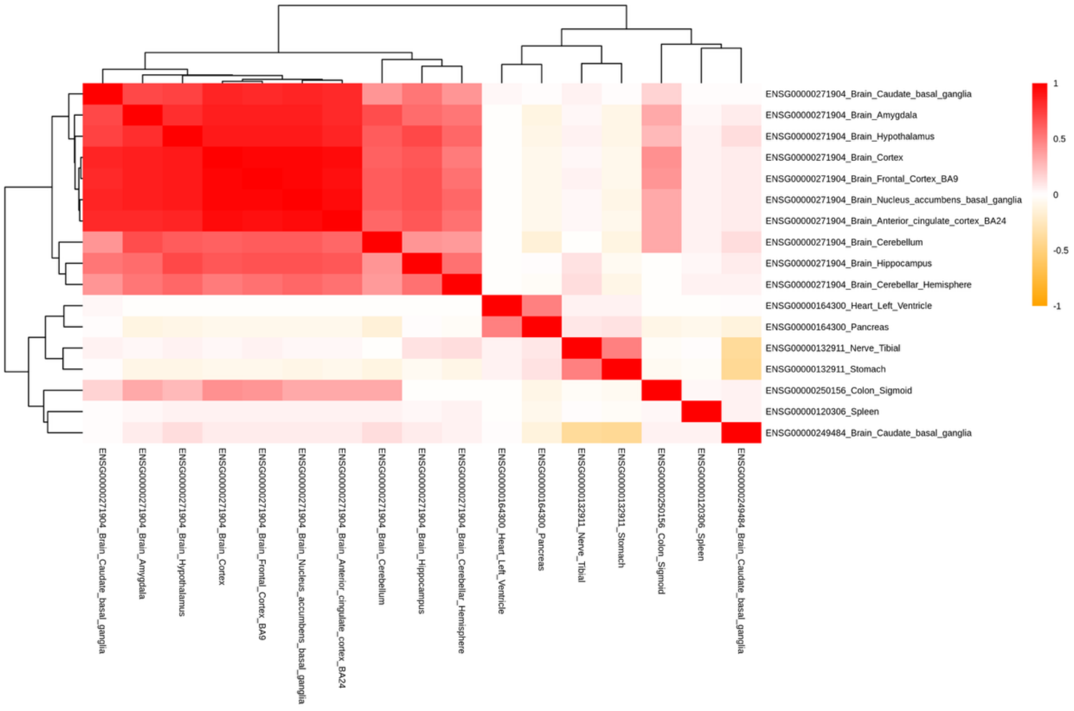


**Supplementary Figure 32. Correlation of GReX on Chromosome 5 for Stuttering in Females Gene-Tissue Pairs.**

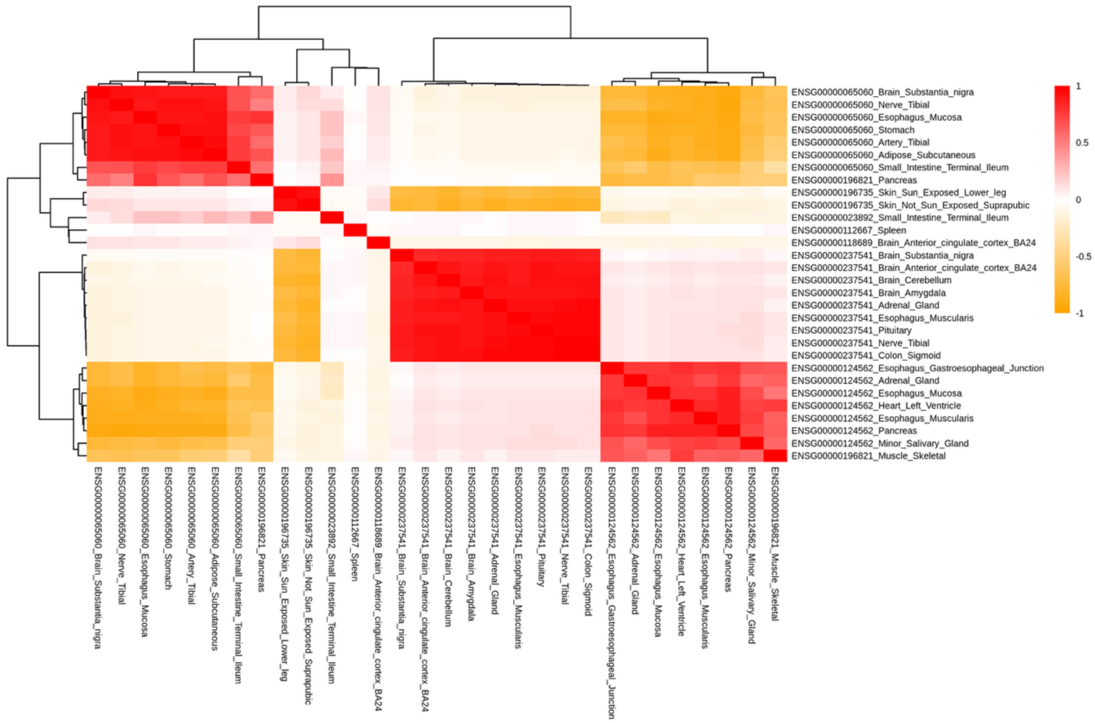

**Supplementary Figure 33. Correlation of GReX on Chromosome 6 for Stuttering in Females Gene-Tissue Pairs.**

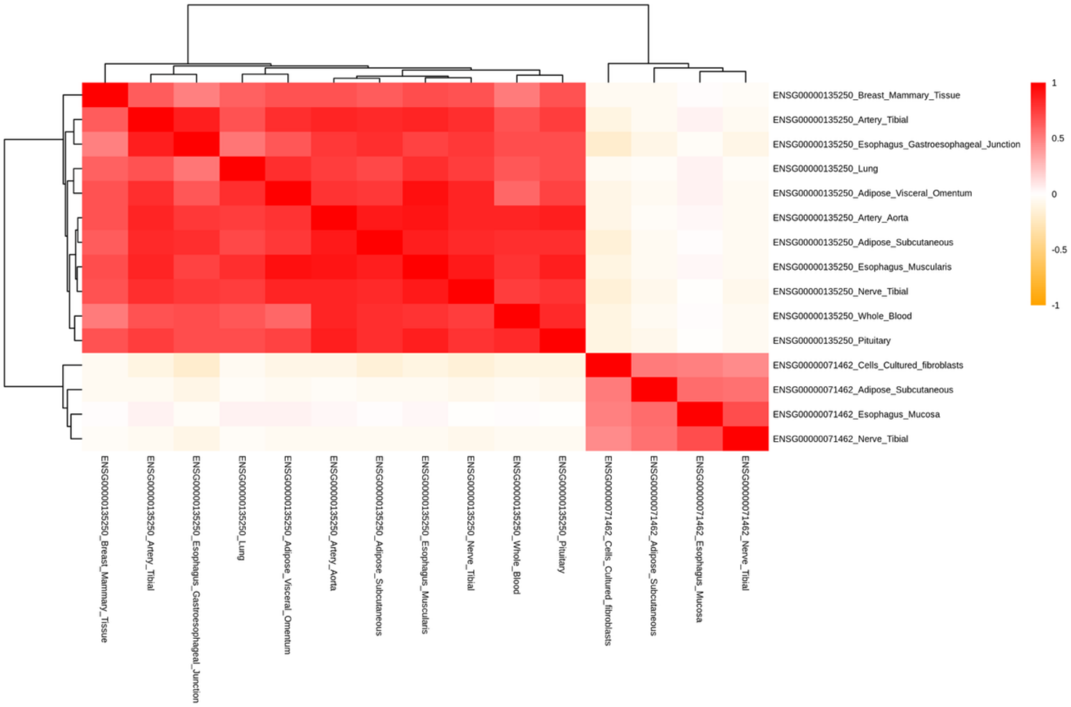


**Supplementary Figure 34. Correlation of GReX on Chromosome 7 for Stuttering in Females Gene-Tissue Pairs.**

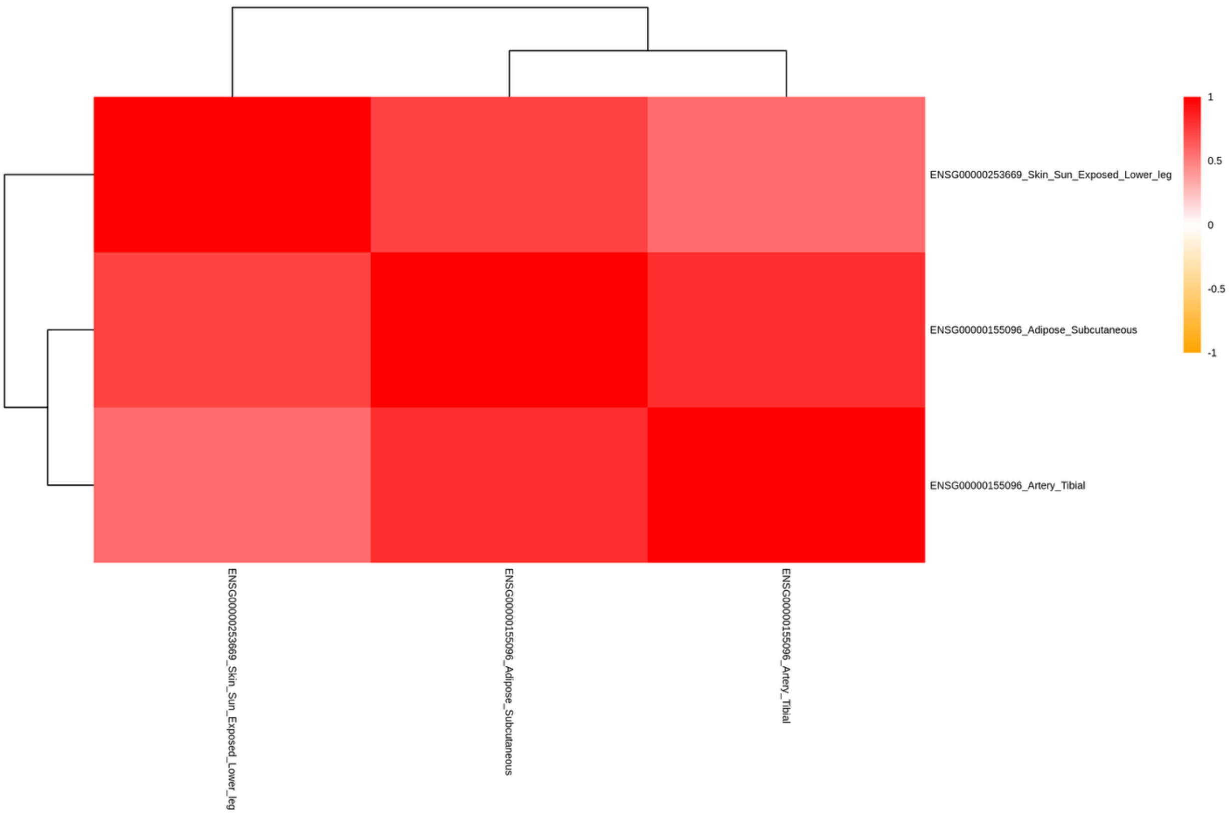


**Supplementary Figure 35. Correlation of GReX on Chromosome 8 for Stuttering in Females Gene-Tissue Pairs.**

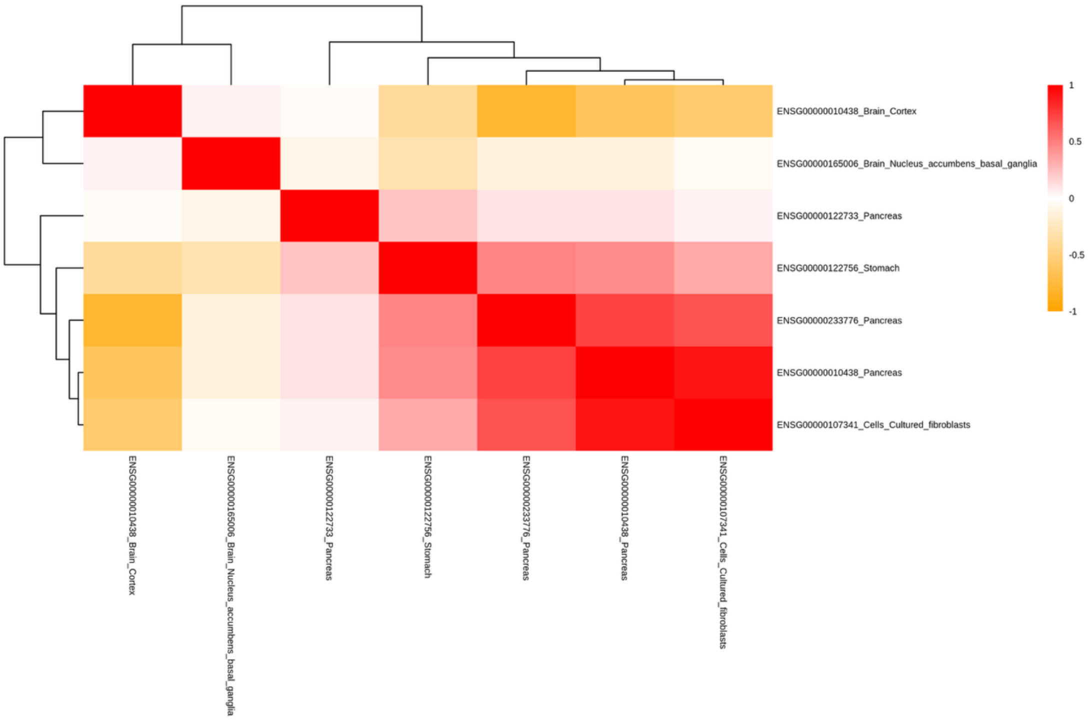

**Supplementary Figure 36. Correlation of GReX on Chromosome 9 for Stuttering in Females Gene-Tissue Pairs.**

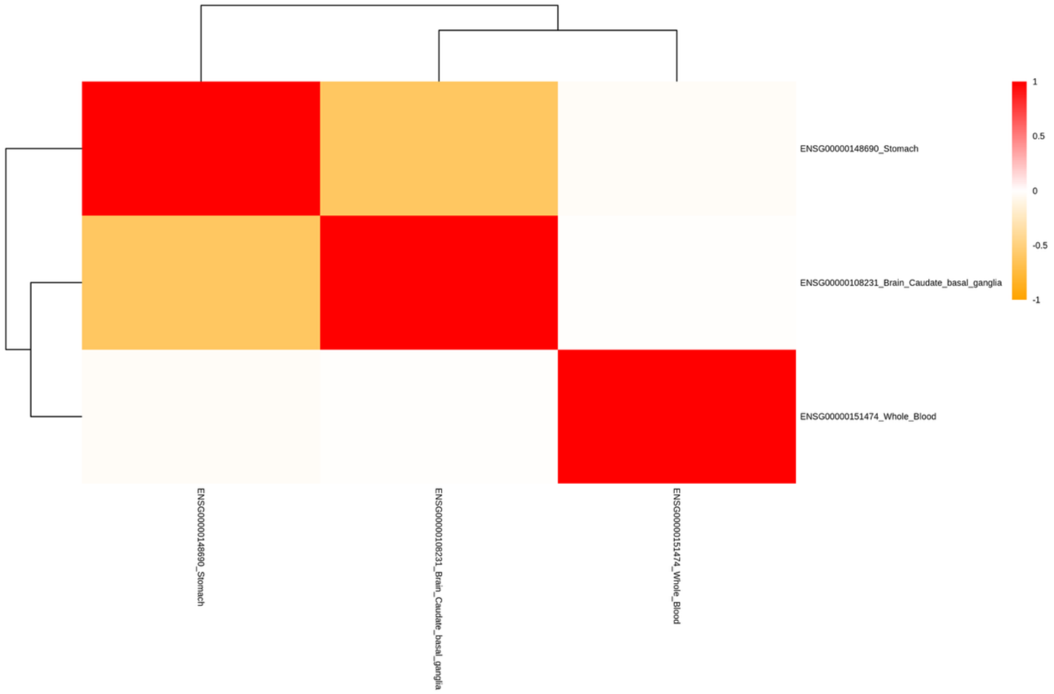


**Supplementary Figure 37. Correlation of GReX on Chromosome 10 for Stuttering in Females Gene-Tissue Pairs.**

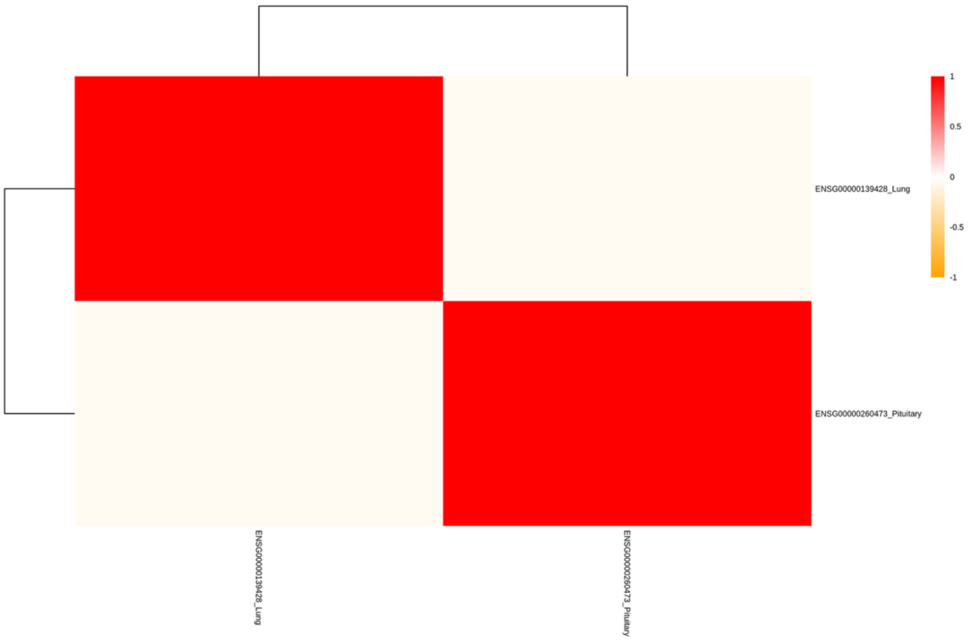

**Supplementary Figure 38. Correlation of GReX on Chromosome 12 for Stuttering in Females Gene-Tissue Pairs.**

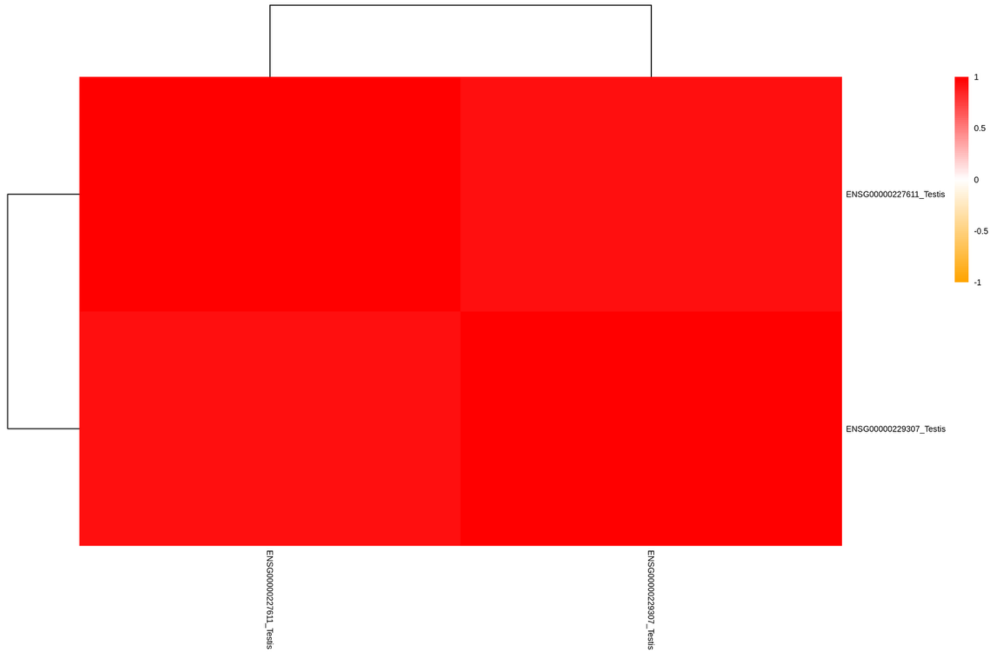


**Supplementary Figure 39. Correlation of GReX on Chromosome 13 for Stuttering in Females Gene-Tissue Pairs.**

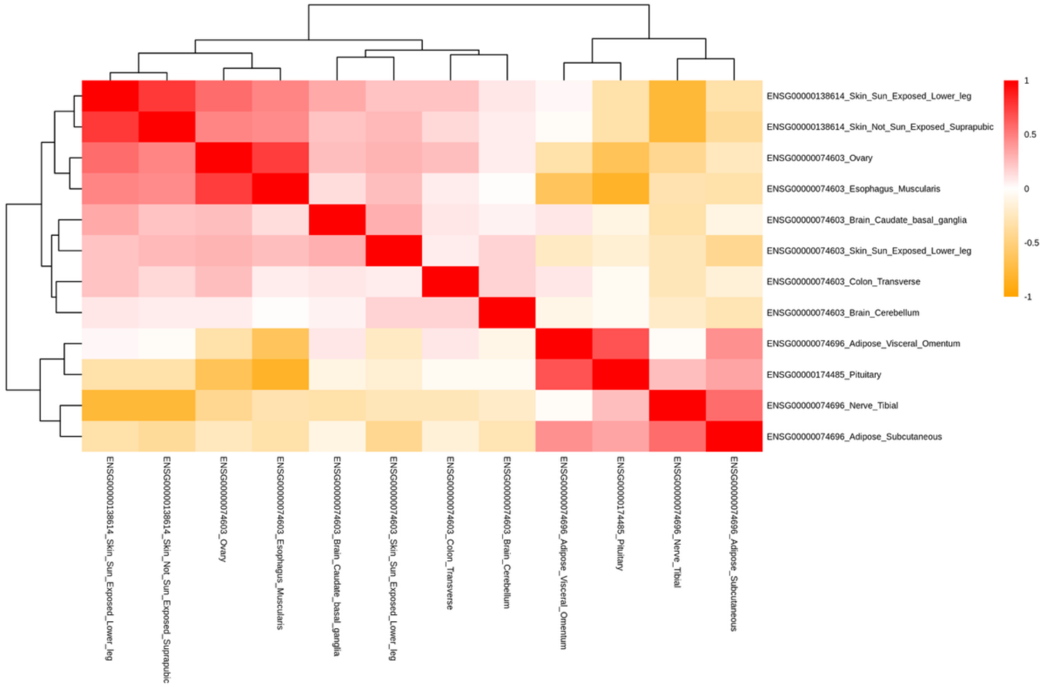

**Supplementary Figure 40. Correlation of GReX on Chromosome 15 for Stuttering in Females Gene-Tissue Pairs.**

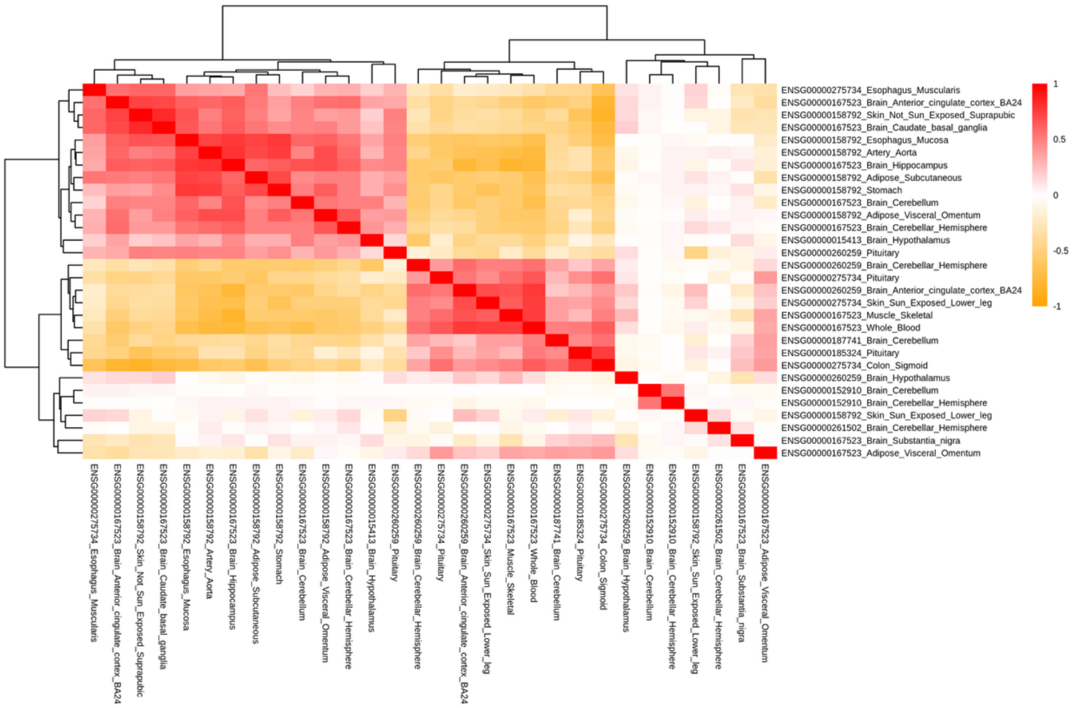


**Supplementary Figure 41. Correlation of GReX on Chromosome 16 for Stuttering in Females Gene-Tissue Pairs.**

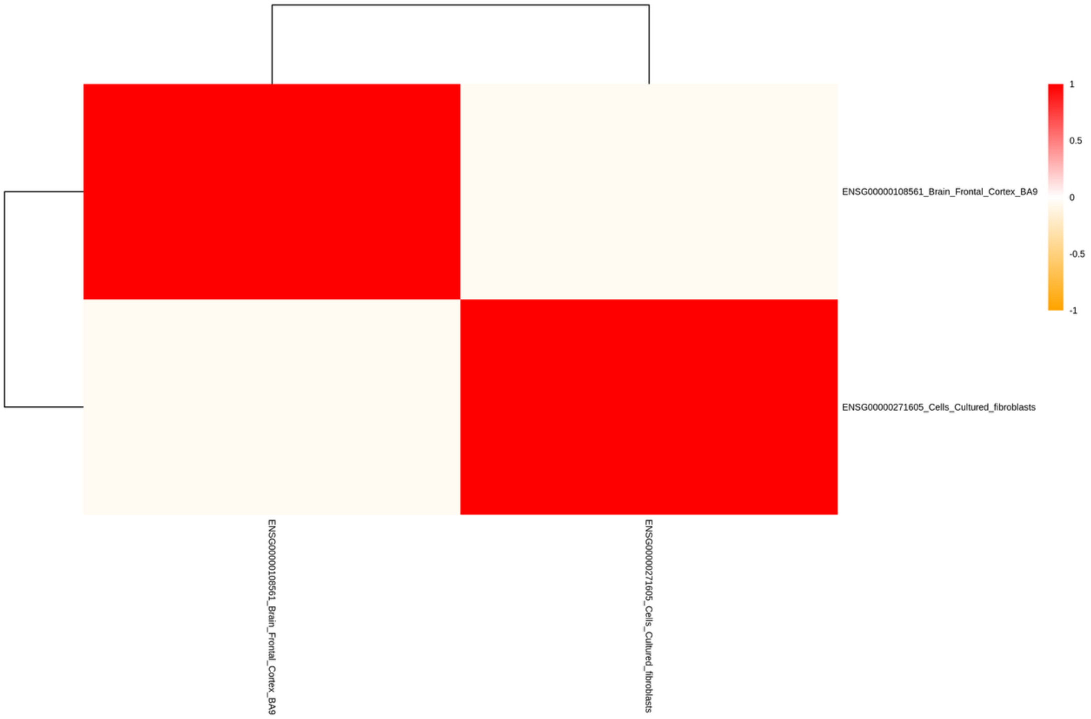

**Supplementary Figure 42. Correlation of GReX on Chromosome 17 for Stuttering in Females Gene-Tissue Pairs.**

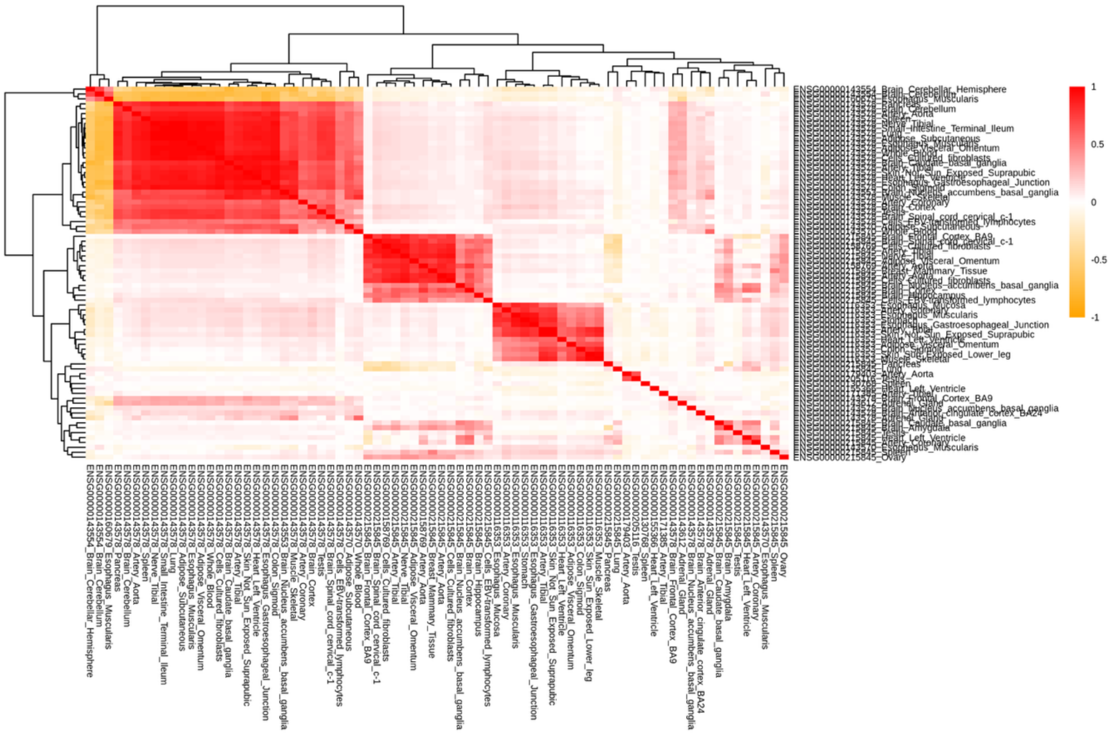

**Supplementary Figure 43. Correlation of GReX on Chromosome 18 for Stuttering in Females Gene-Tissue Pairs.**

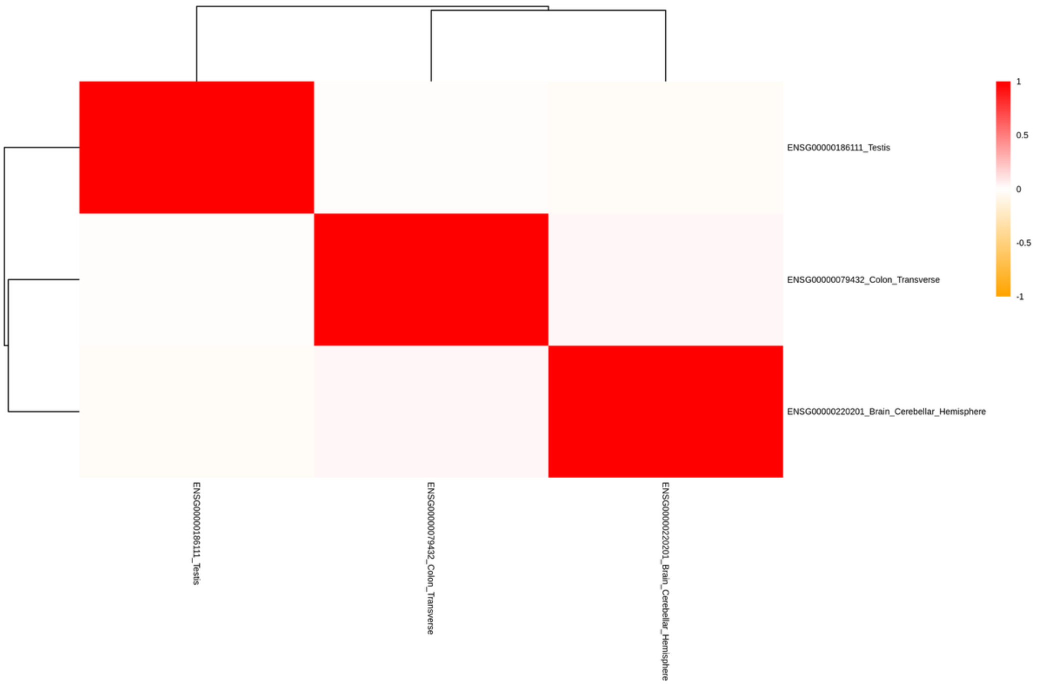

**Supplementary Figure 44. Correlation of GReX on Chromosome 19 for Stuttering in Females Gene-Tissue Pairs.**

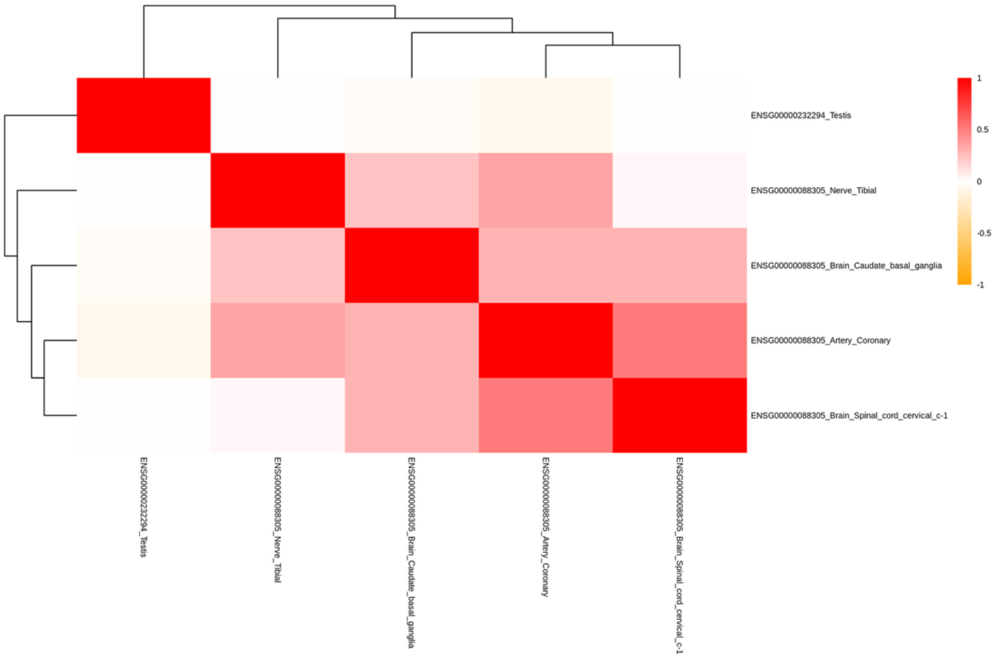


**Supplementary Figure 45. Correlation of GReX on Chromosome 20 for Stuttering in Females Gene-Tissue Pairs.**

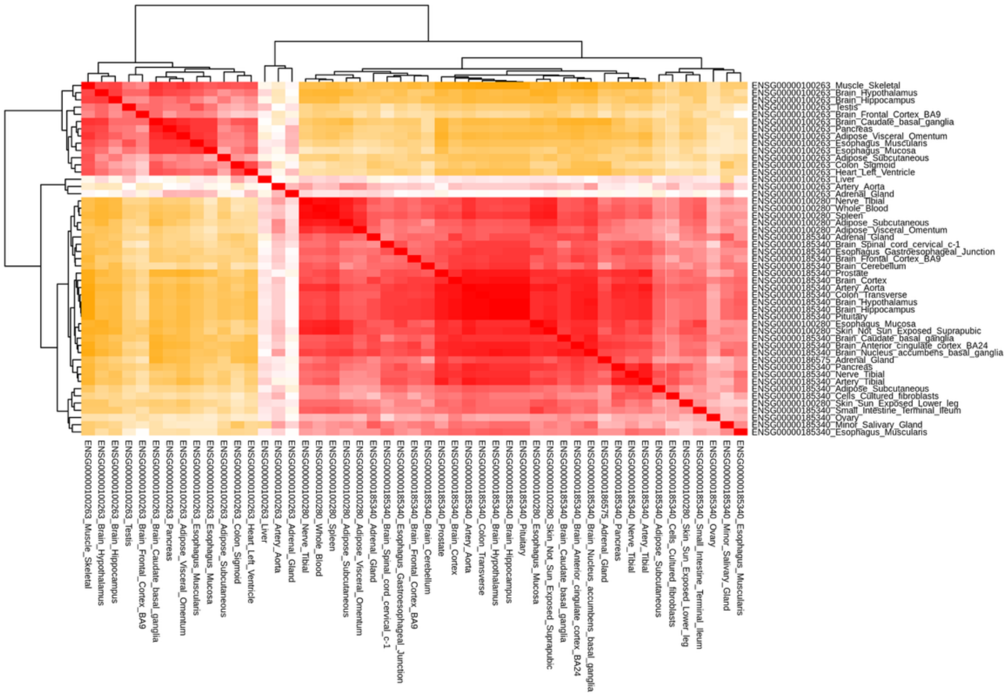


**Supplementary Figure 46. Correlation of GReX on Chromosome 22 for Stuttering in Females Gene-Tissue Pairs.**

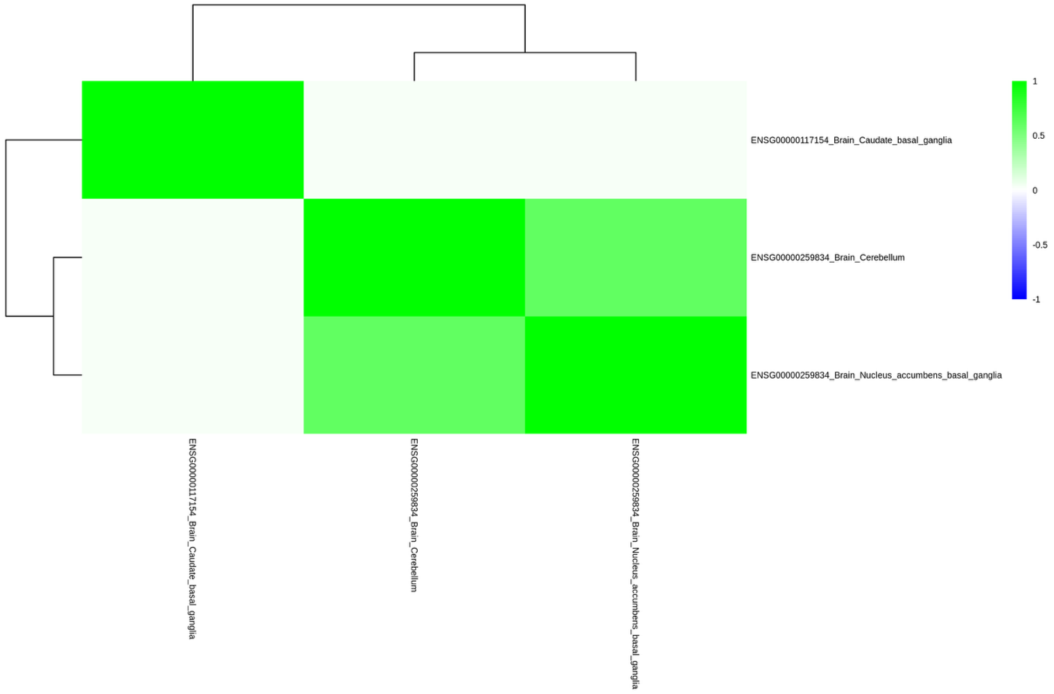


**Supplementary Figure 47. Correlation of GReX on Chromosome 1 for Stuttering in Males Gene-Tissue Pairs.**

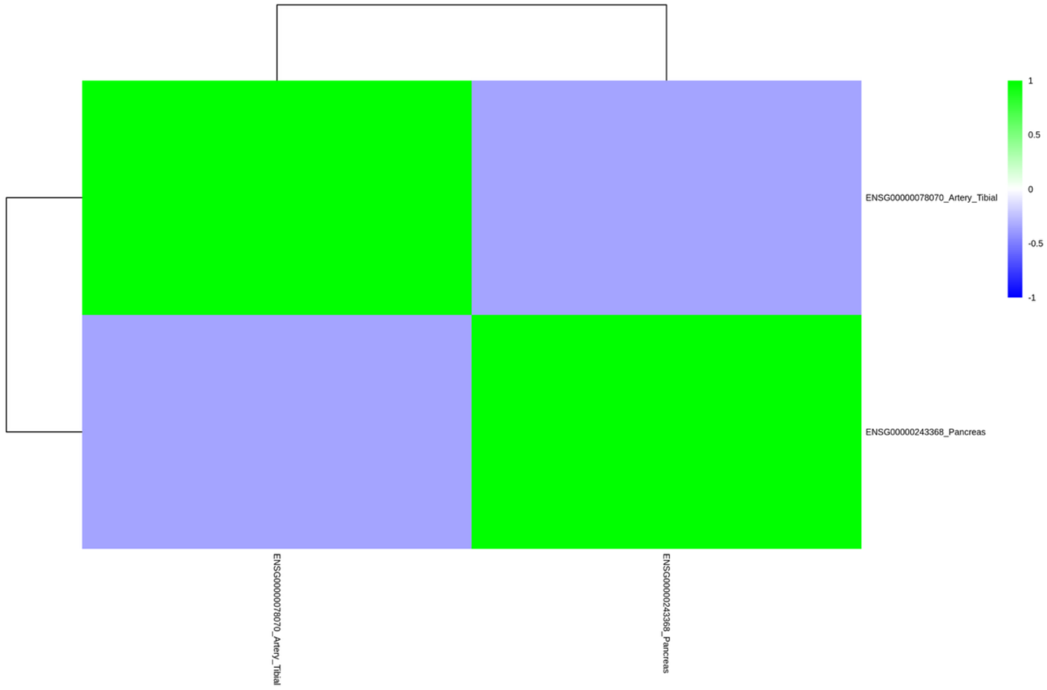


**Supplementary Figure 48. Correlation of GReX on Chromosome 3 for Stuttering in Males Gene-Tissue Pairs.**

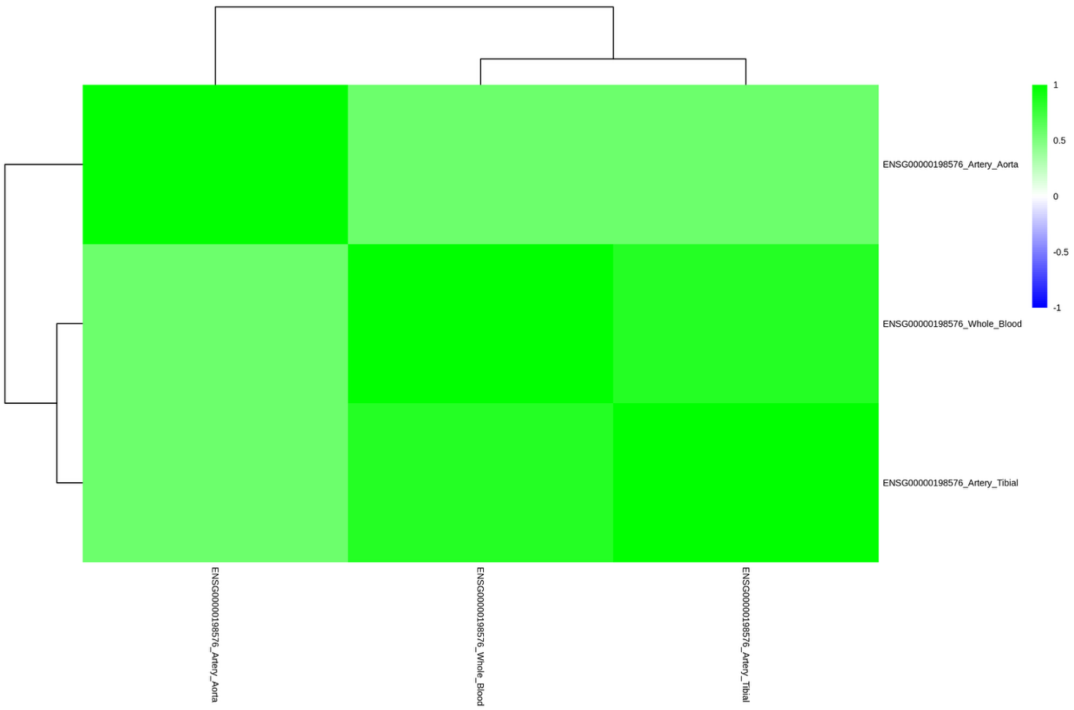


**Supplementary Figure 49. Correlation of GReX on Chromosome 8 for Stuttering in Males Gene-Tissue Pairs.**

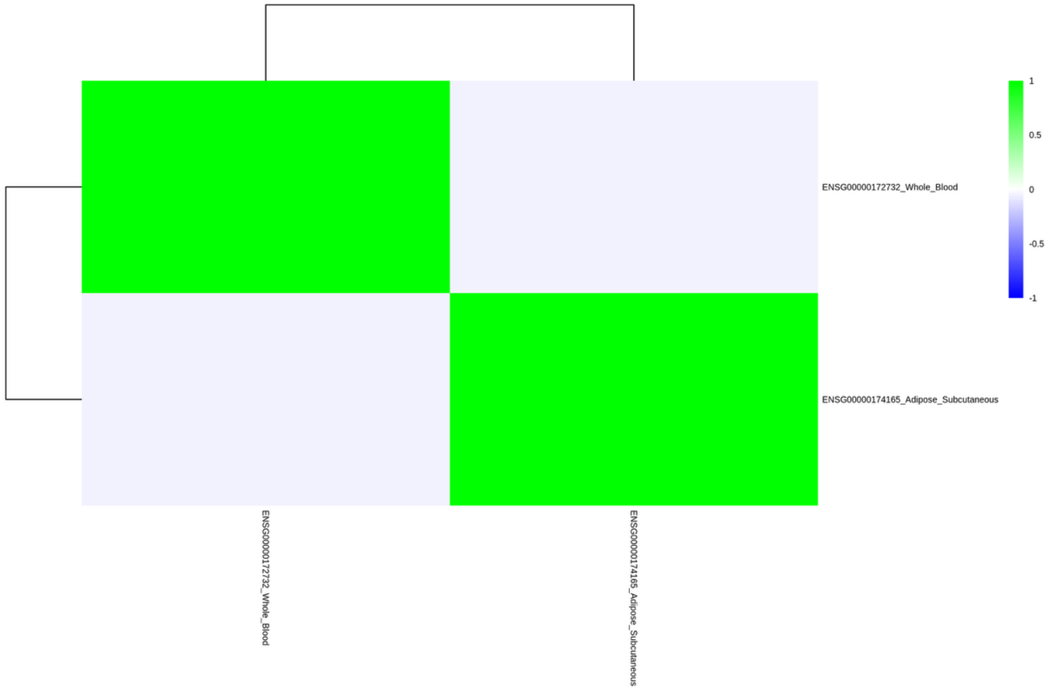


**Supplementary Figure 50. Correlation of GReX on Chromosome 11 for Stuttering in Males Gene-Tissue Pairs.**

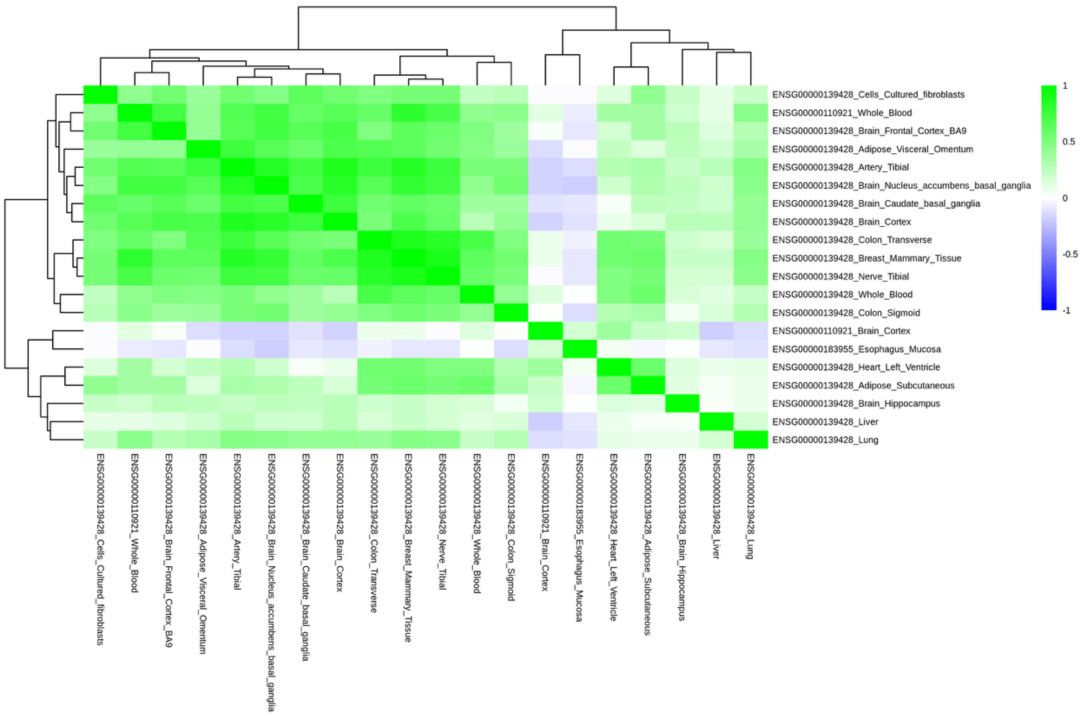


**Supplementary Figure 51. Correlation of GReX on Chromosome 12 for Stuttering in Males Gene-Tissue Pairs.**

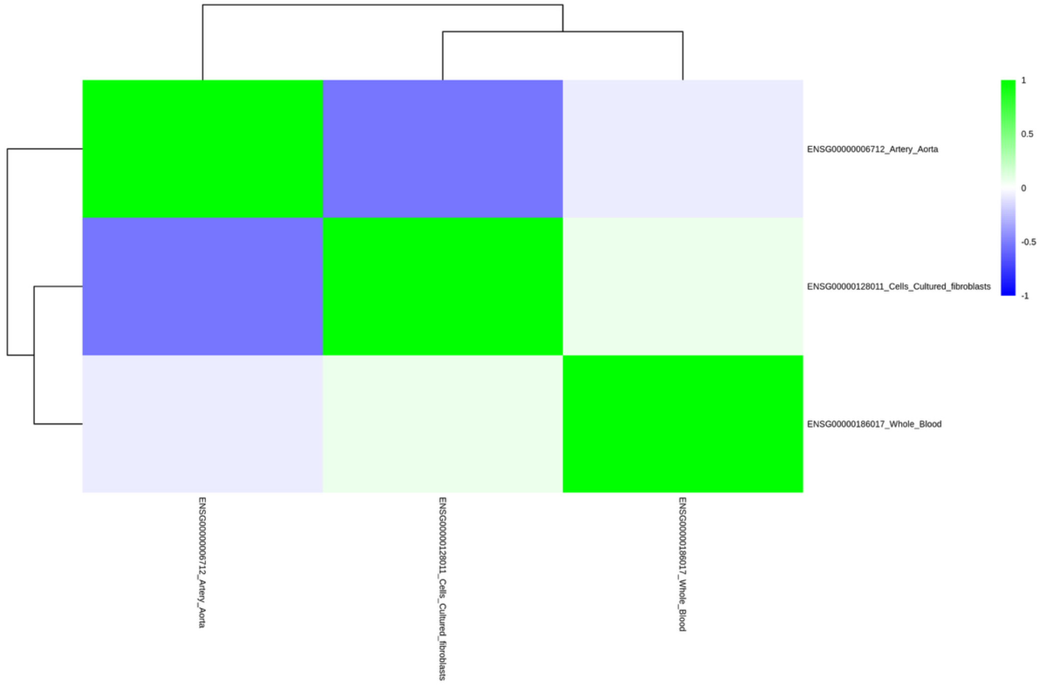


**Supplementary Figure 52. Correlation of GReX on Chromosome 19 for Stuttering in Males Gene-Tissue Pairs.**

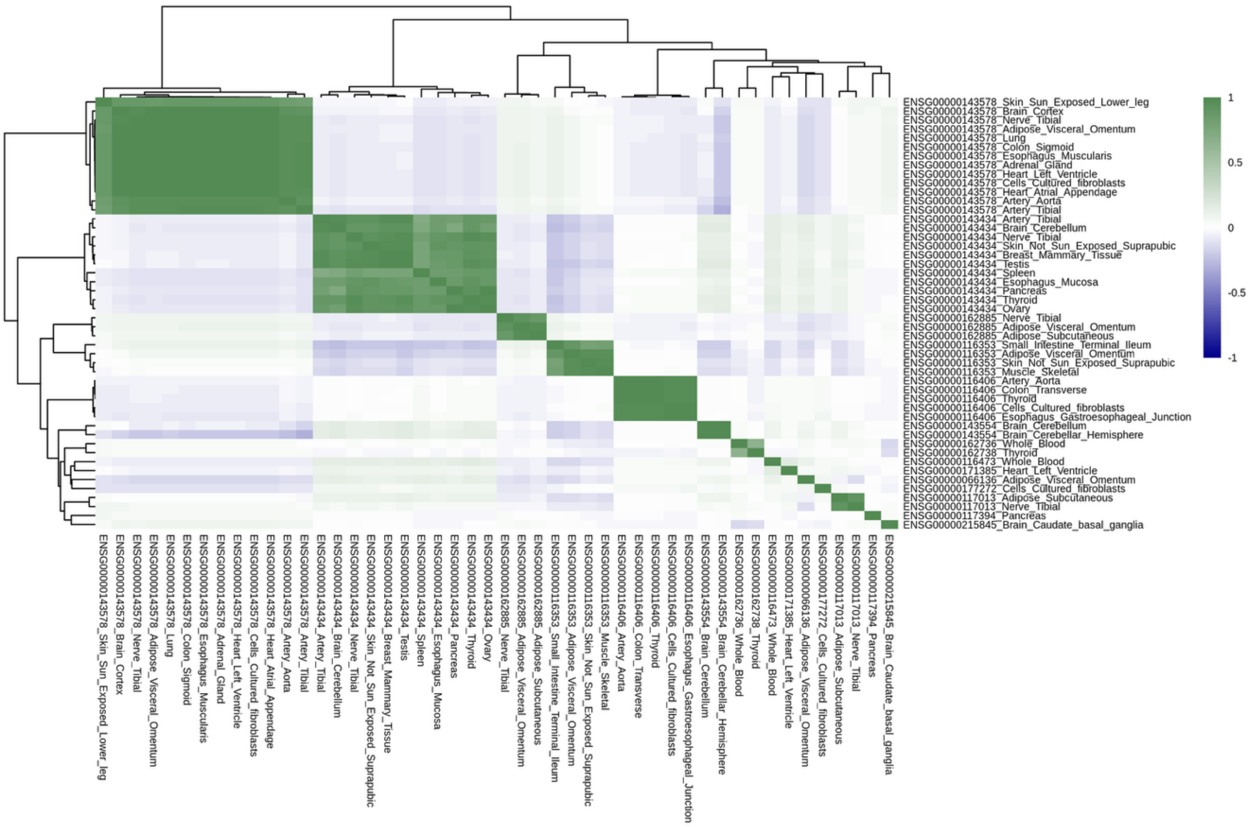


**Supplementary Figure 53. Correlation of GReX on Chromosome 1 for Sex-combined Stuttering for our MR Analyses.**

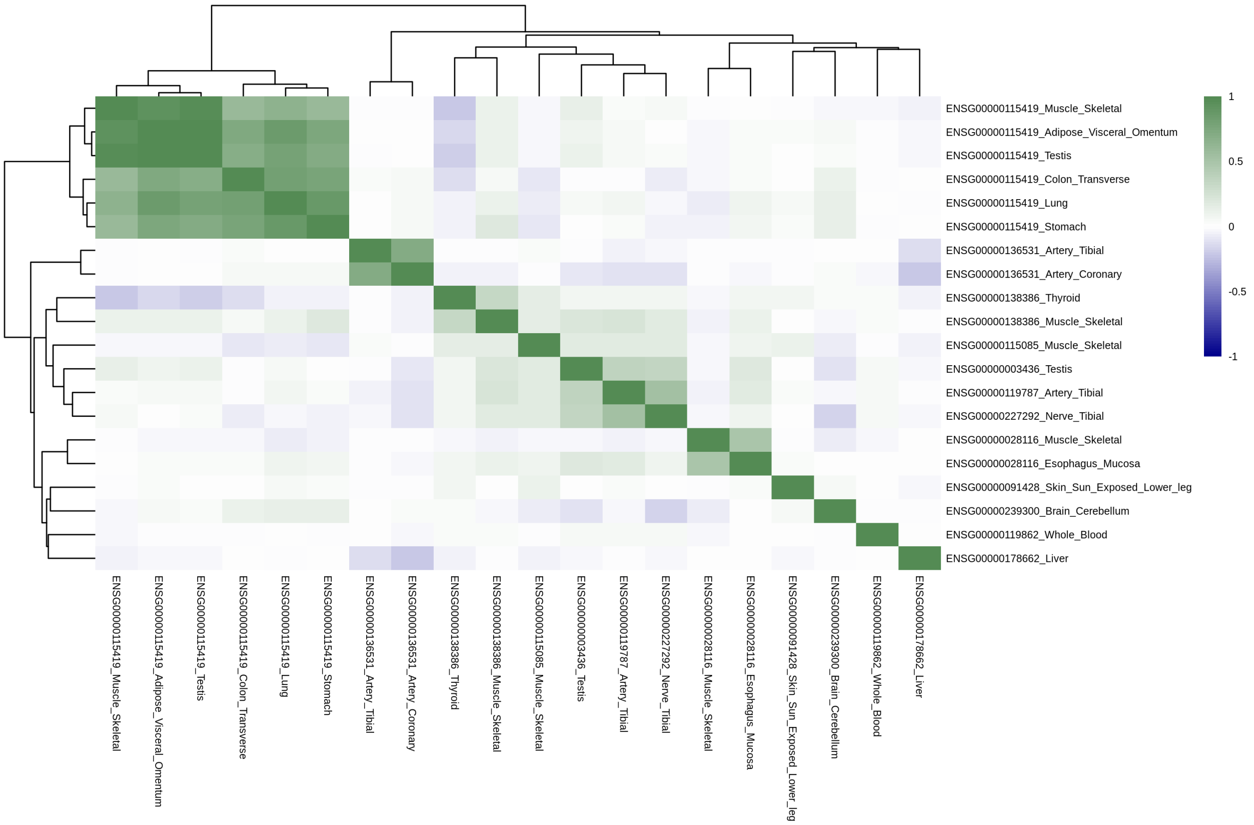


**Supplementary Figure 54. Correlation of GReX on Chromosome 2 for Sex-combined Stuttering for our MR Analyses.**

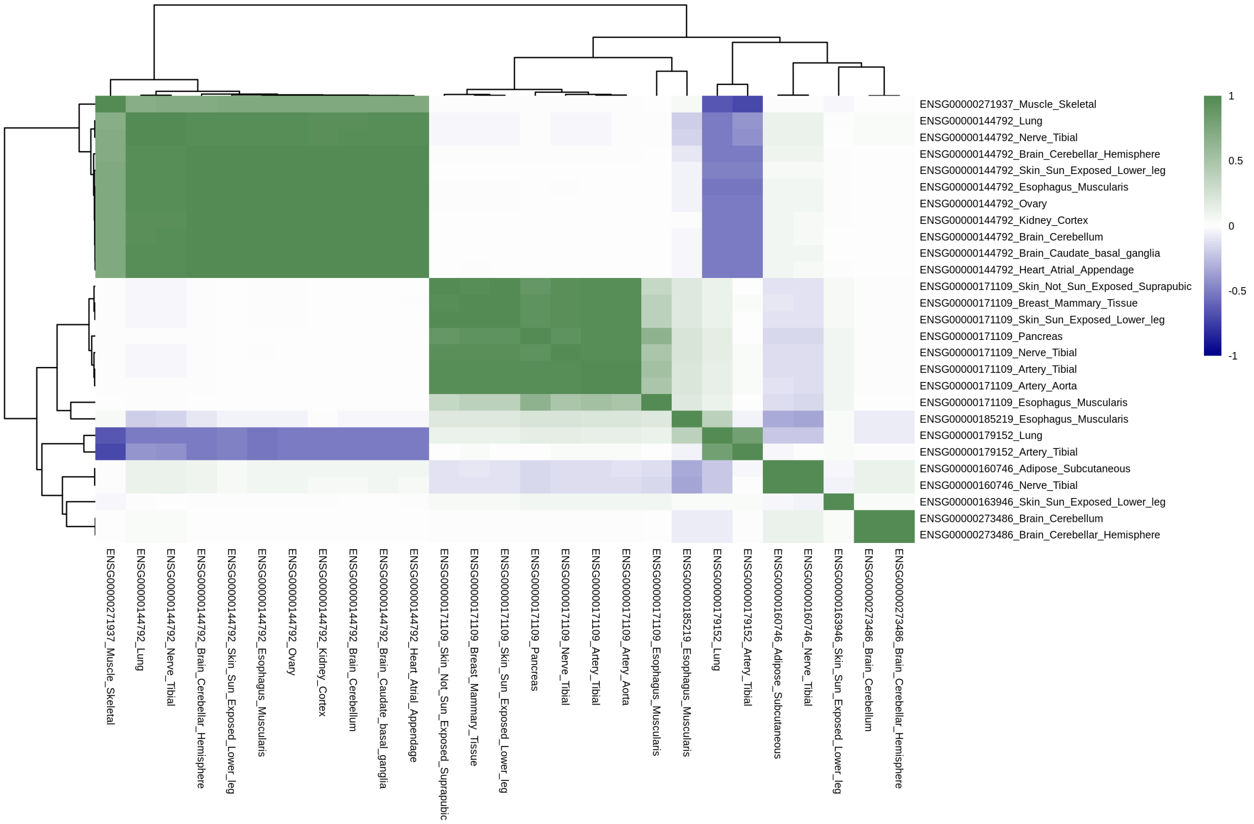


**Supplementary Figure 55. Correlation of GReX on Chromosome 3 for Sex-combined Stuttering for our MR Analyses.**

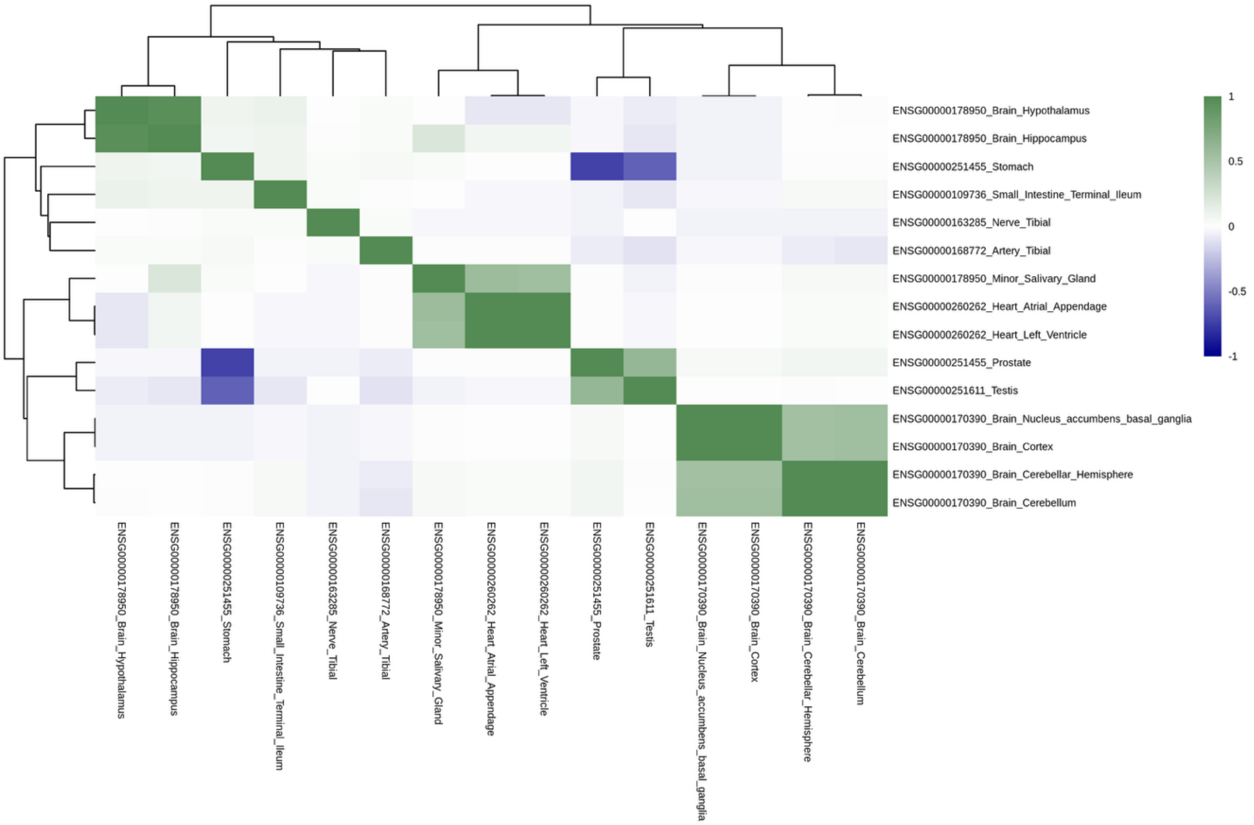


**Supplementary Figure 56. Correlation of GReX on Chromosome 4 for Sex-combined Stuttering for our MR Analyses.**

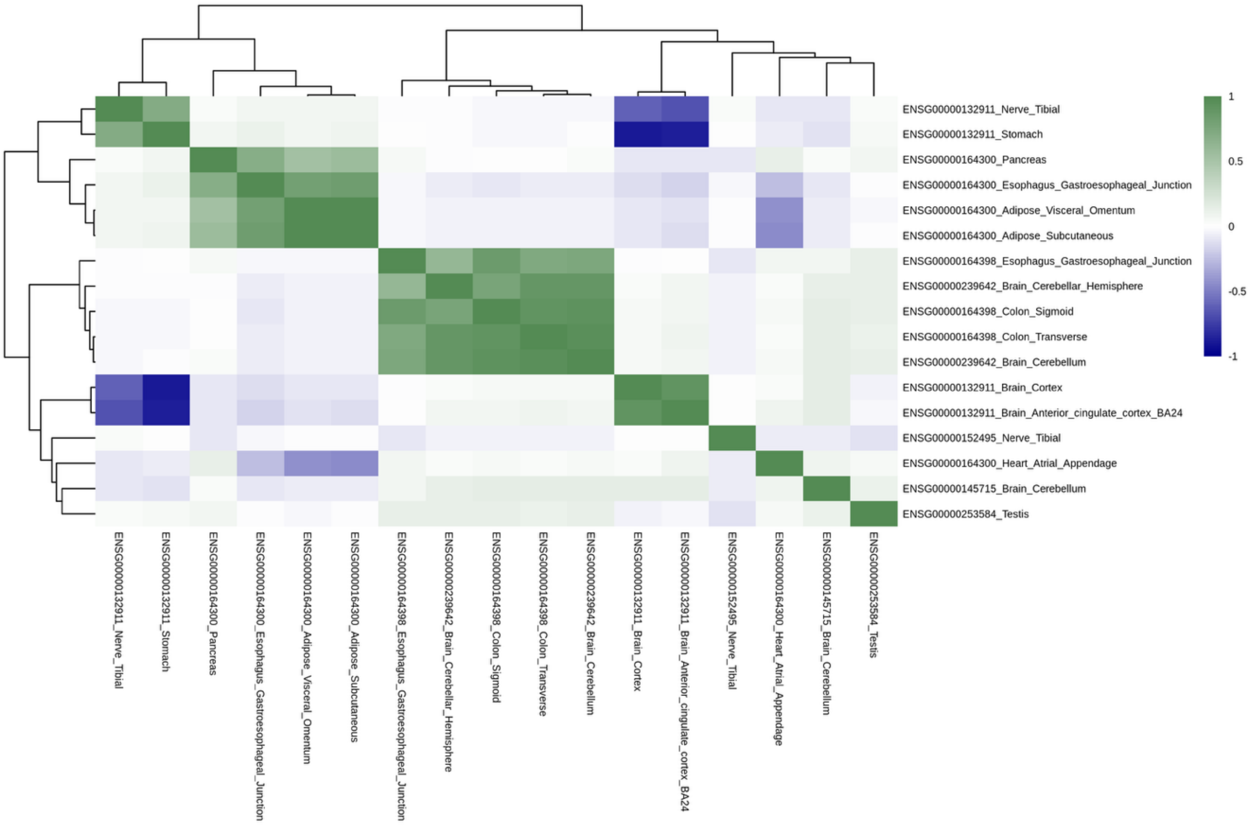


**Supplementary Figure 57. Correlation of GReX on Chromosome 5 for Sex-combined Stuttering for our MR Analyses.**

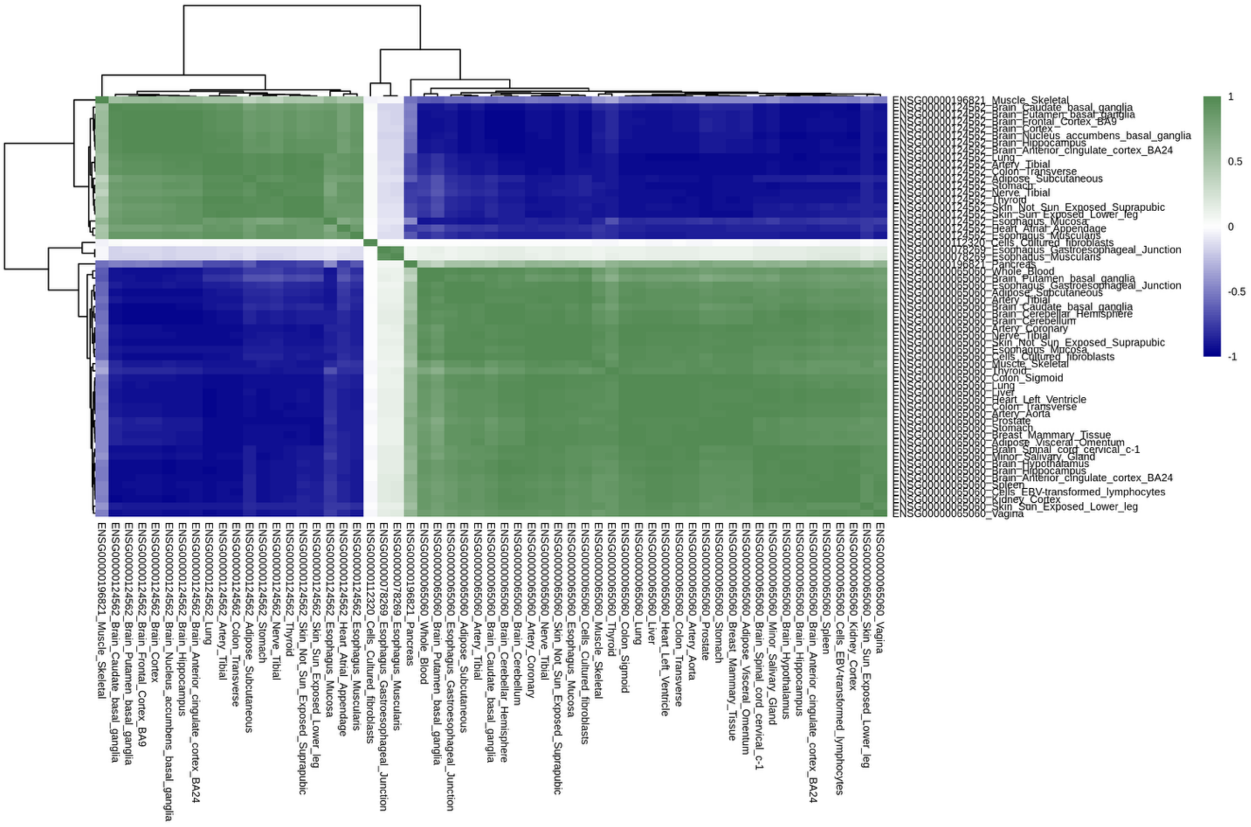


**Supplementary Figure 58. Correlation of GReX on Chromosome 6 for Sex-combined Stuttering for our MR Analyses.**

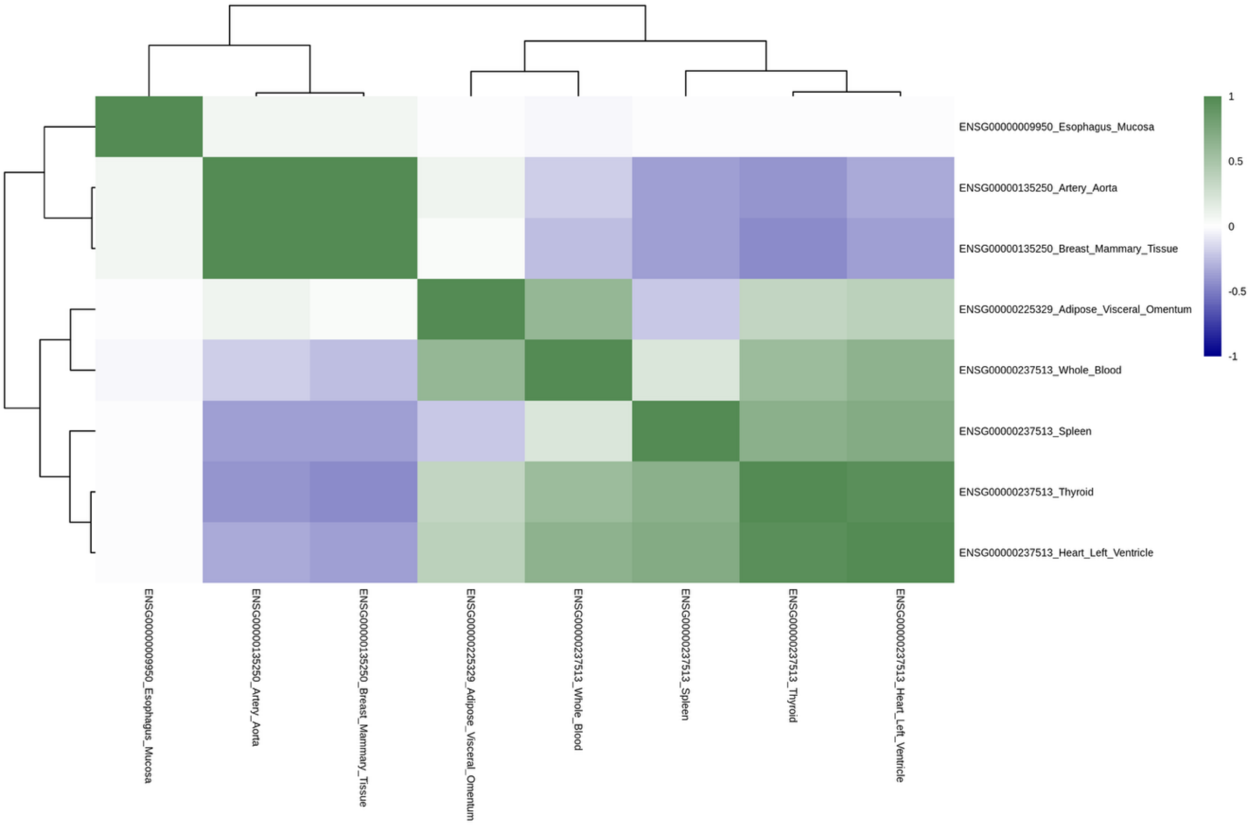


**Supplementary Figure 59. Correlation of GReX on Chromosome 7 for Sex-combined Stuttering for our MR Analyses.**

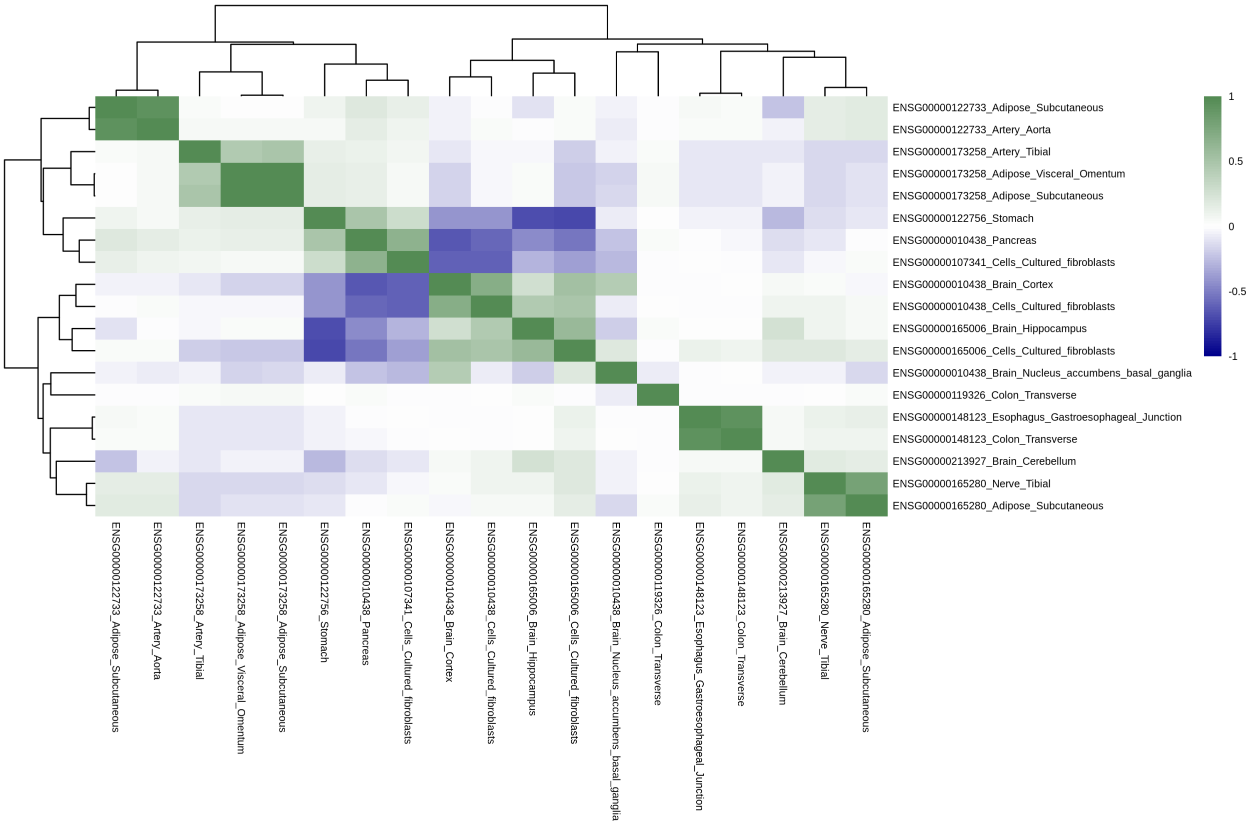


**Supplementary Figure 60. Correlation of GReX on Chromosome 9 for Sex-combined Stuttering for our MR Analyses.**

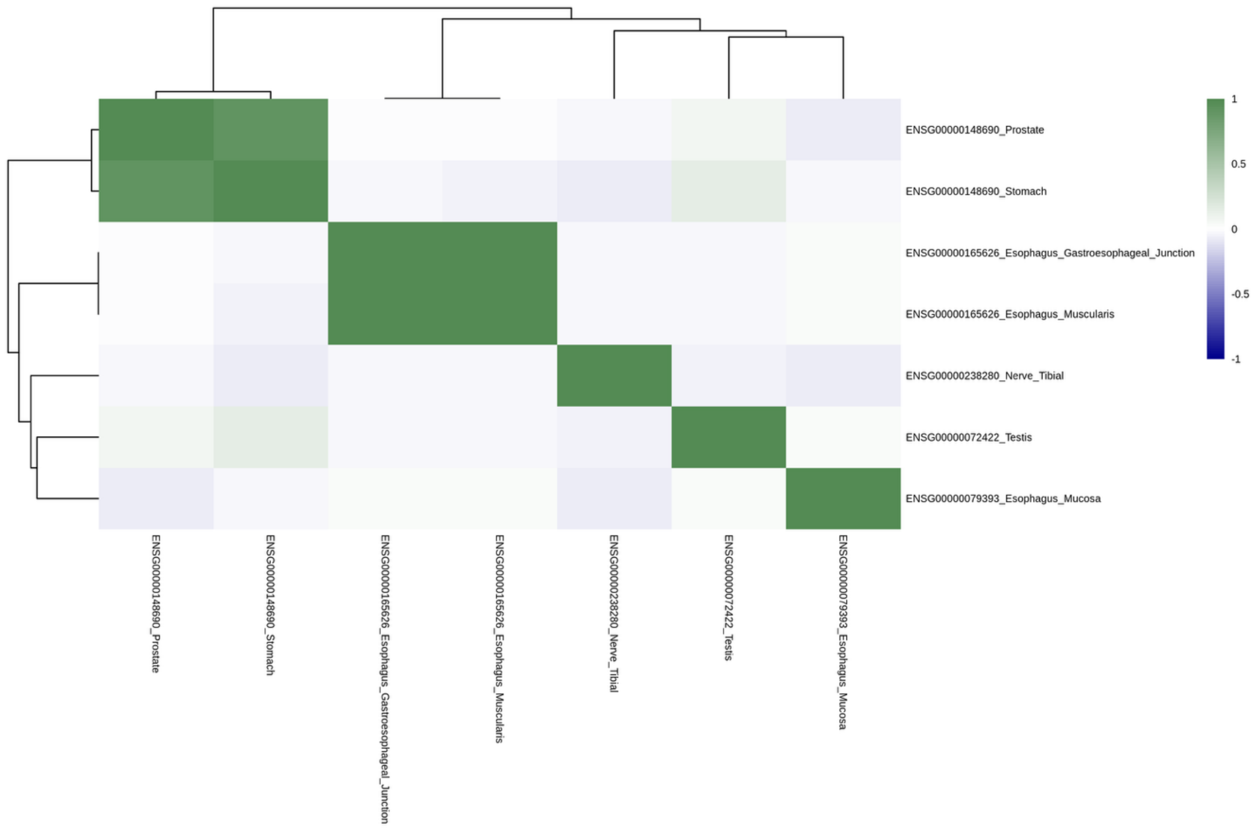


**Supplementary Figure 61. Correlation of GReX on Chromosome 10 for Sex-combined Stuttering for our MR Analyses.**

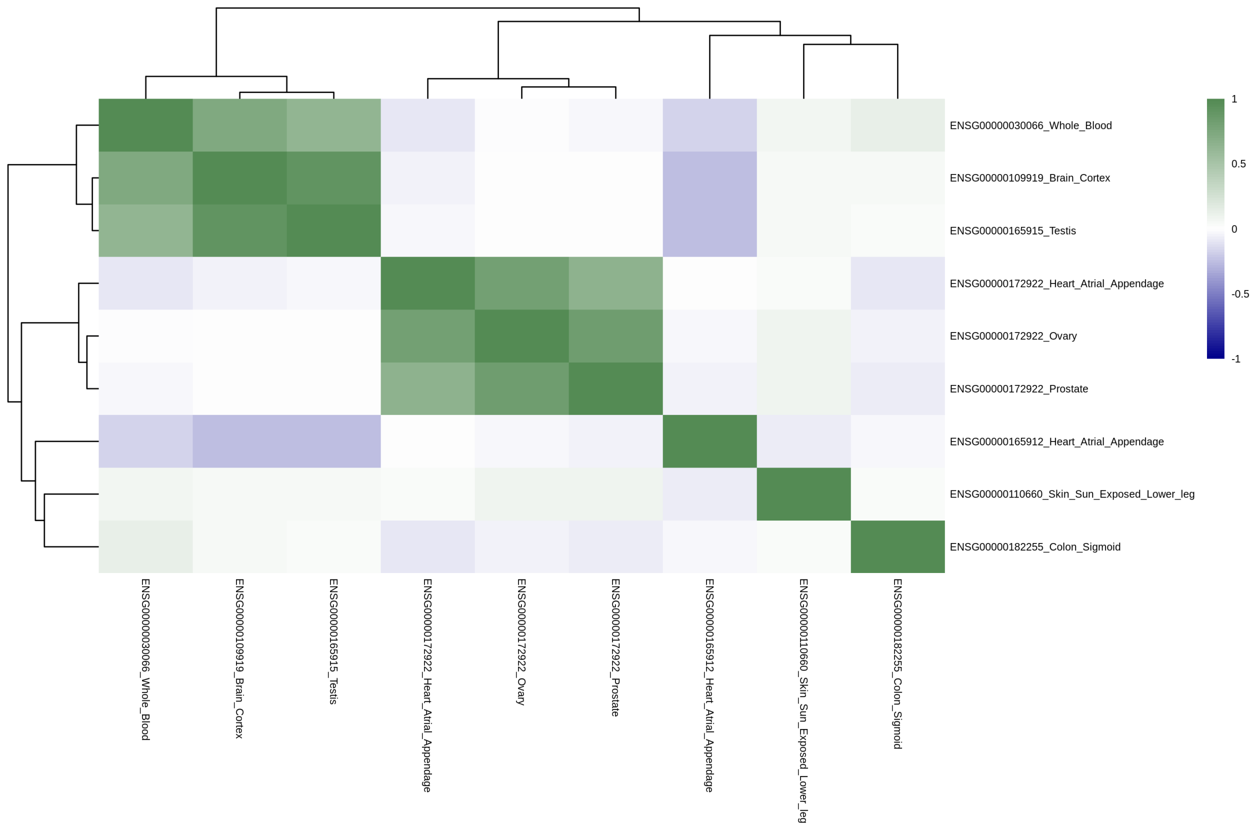


**Supplementary Figure 62. Correlation of GReX on Chromosome 11 for Sex-combined Stuttering for our MR Analyses.**

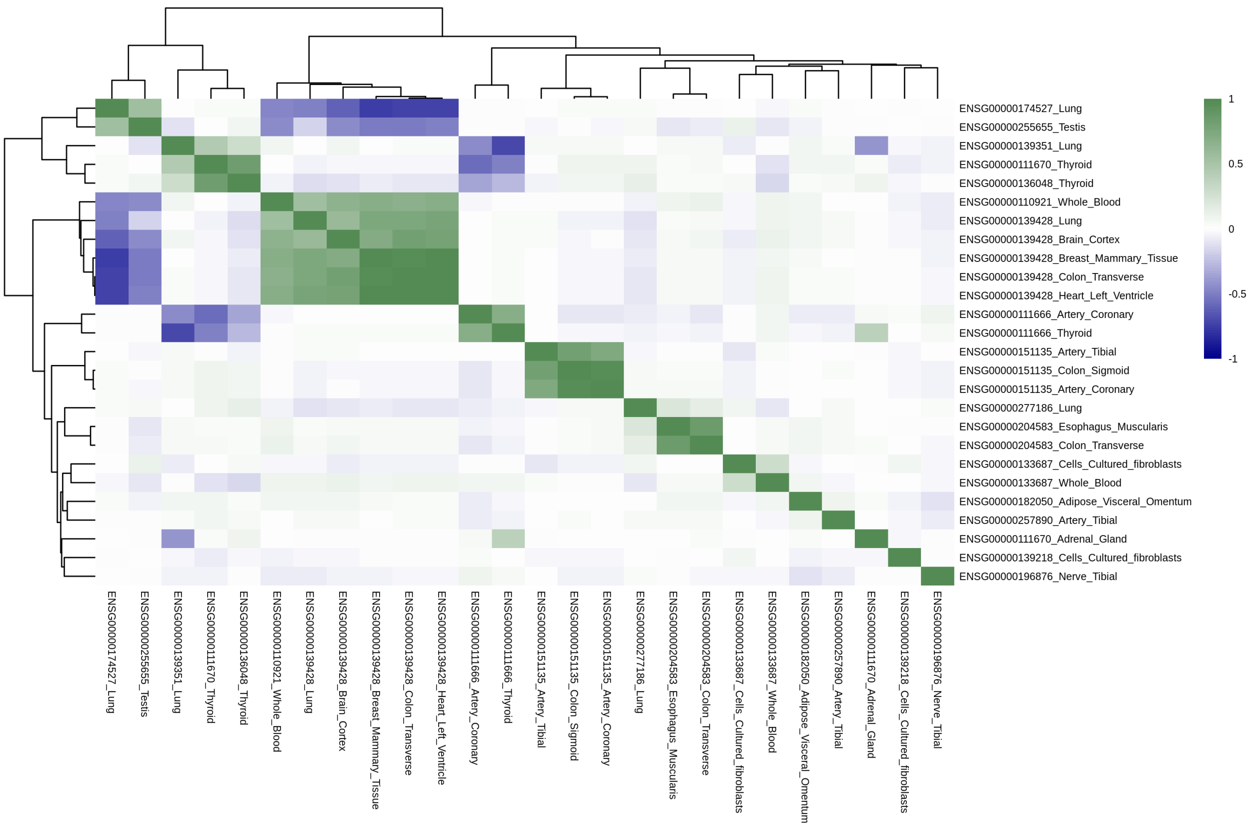


**Supplementary Figure 63. Correlation of GReX on Chromosome 12 for Sex-combined Stuttering for our MR Analyses.**

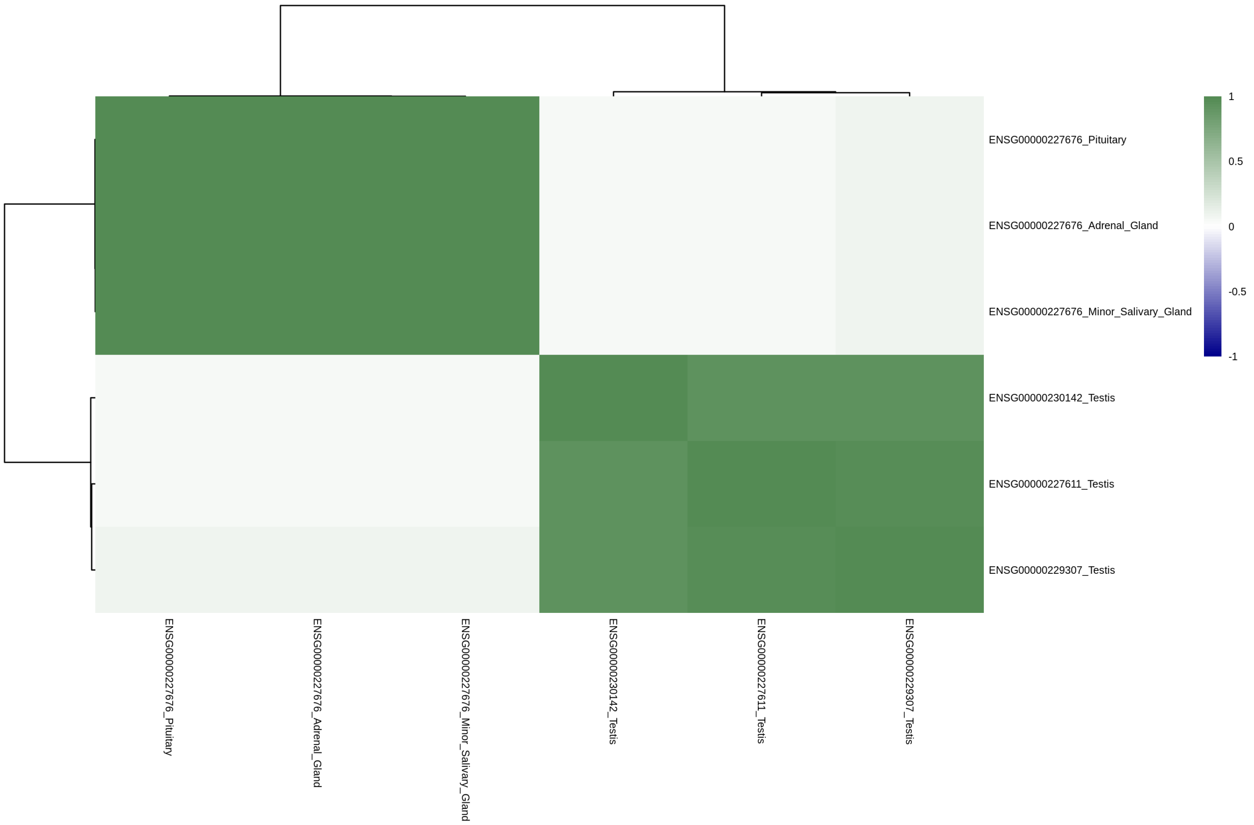


**Supplementary Figure 64. Correlation of GReX on Chromosome 13 for Sex-combined Stuttering for our MR Analyses.**

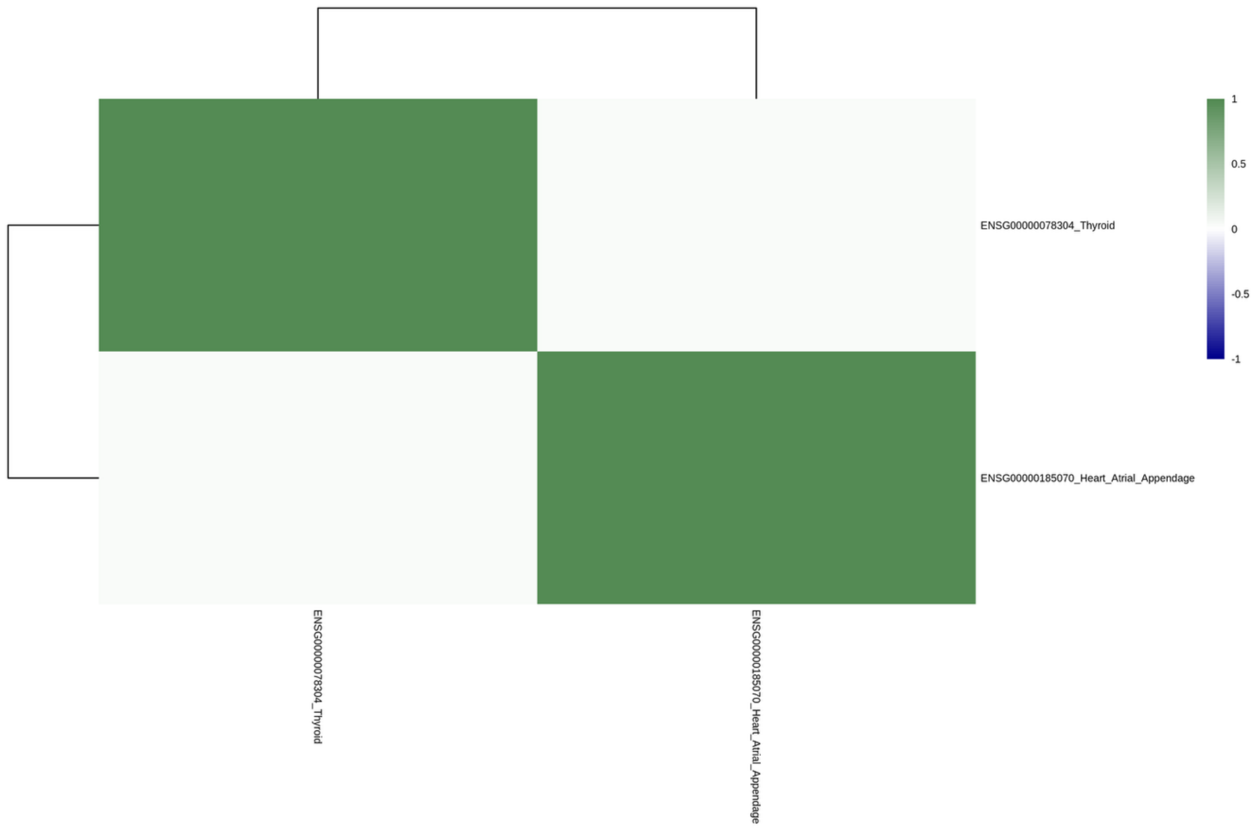


**Supplementary Figure 65. Correlation of GReX on Chromosome 14 for Sex-combined Stuttering for our MR Analyses.**

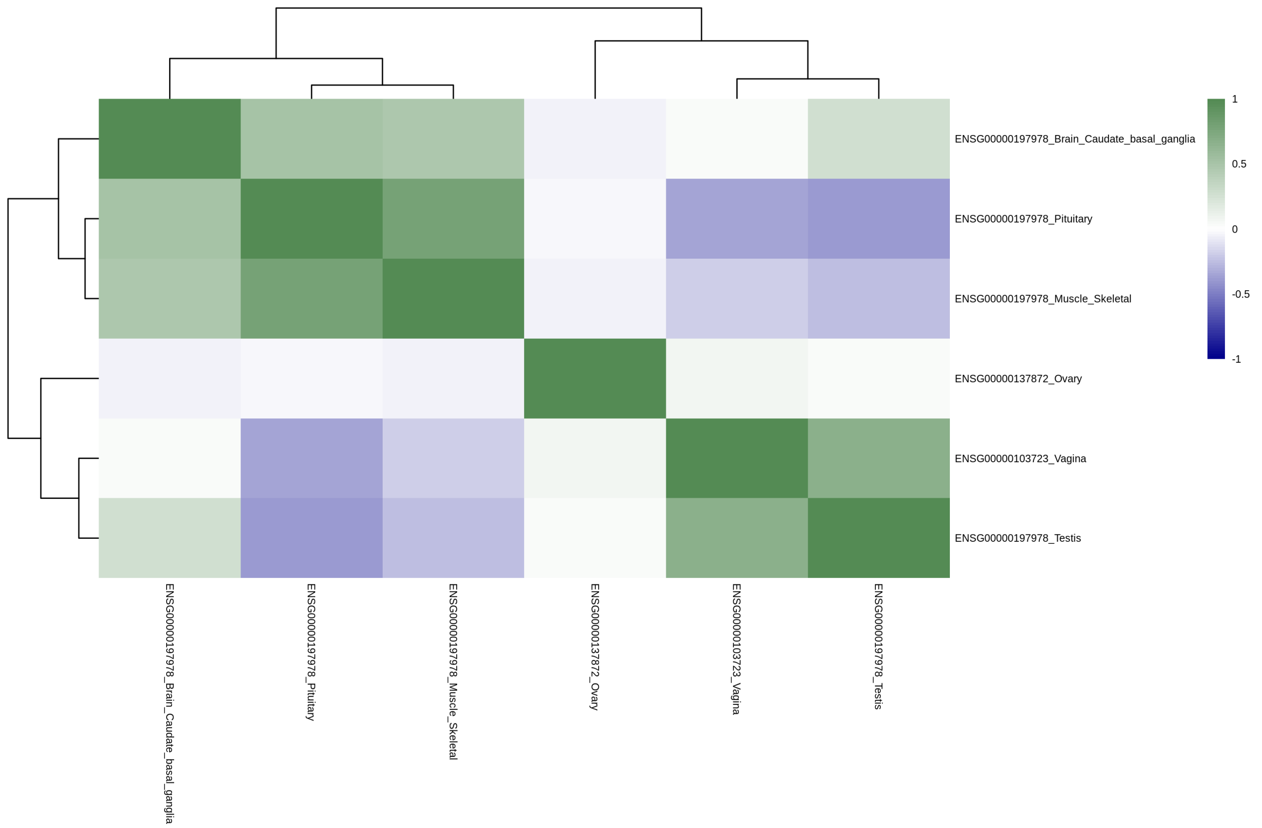


**Supplementary Figure 66. Correlation of GReX on Chromosome 15 for Sex-combined Stuttering for our MR Analyses.**

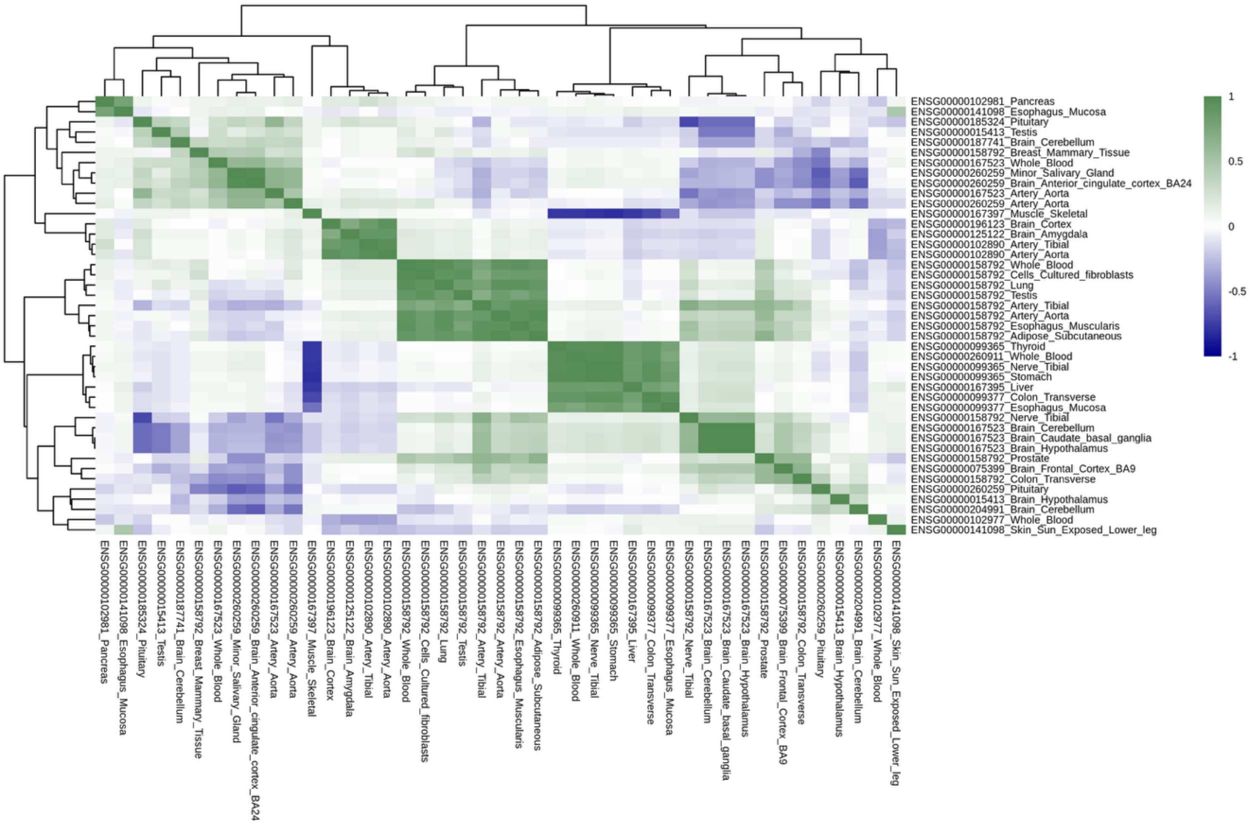


**Supplementary Figure 67. Correlation of GReX on Chromosome 16 for Sex-combined Stuttering for our MR Analyses.**

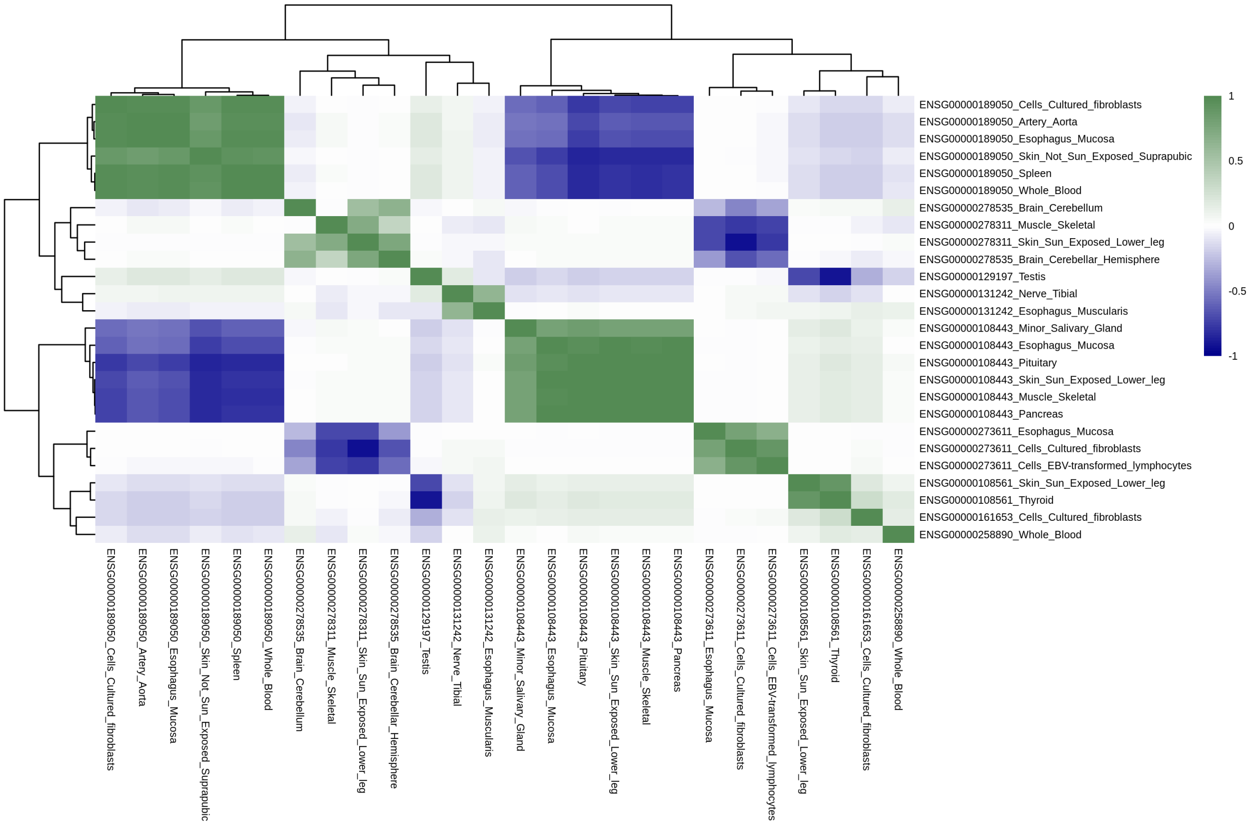


**Supplementary Figure 66. Correlation of GReX on Chromosome 17 for Sex-combined Stuttering for our MR Analyses.**

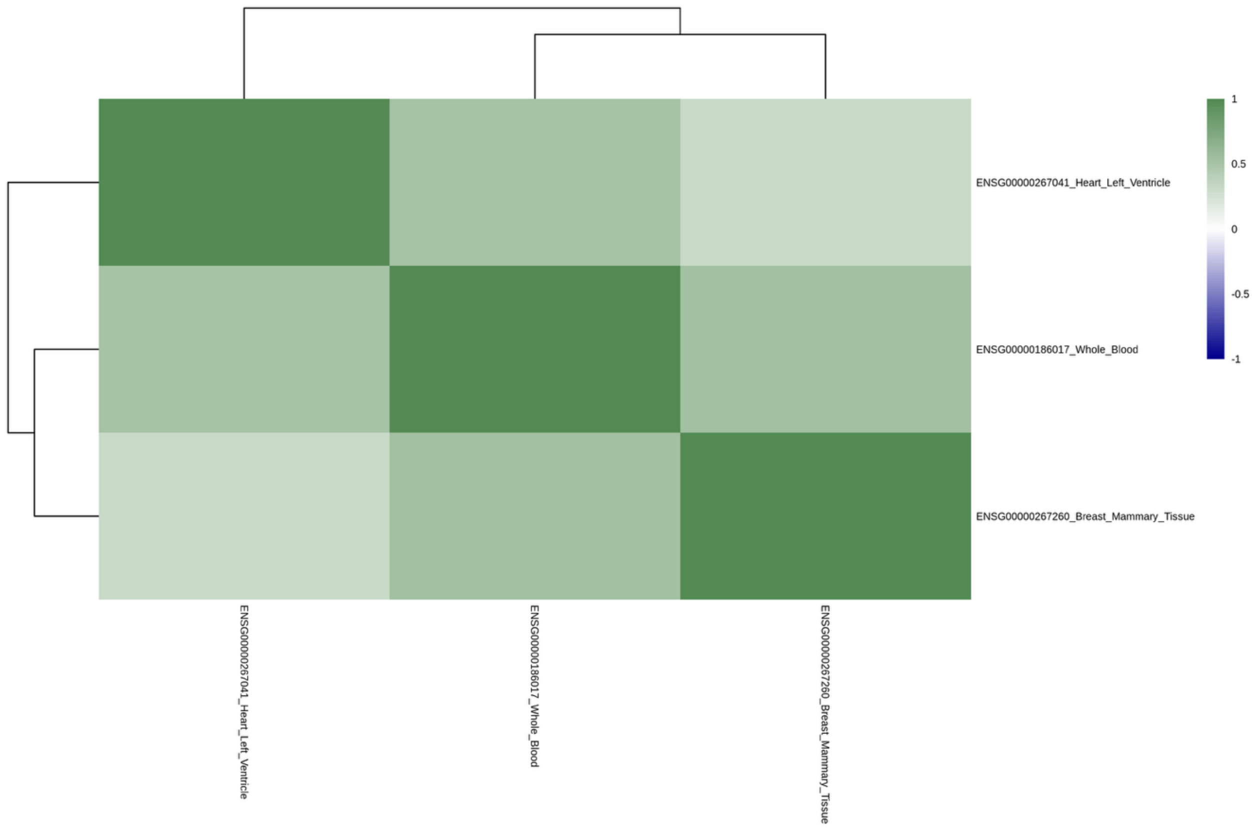


**Supplementary Figure 67. Correlation of GReX on Chromosome 19 for Sex-combined Stuttering for our MR Analyses.**

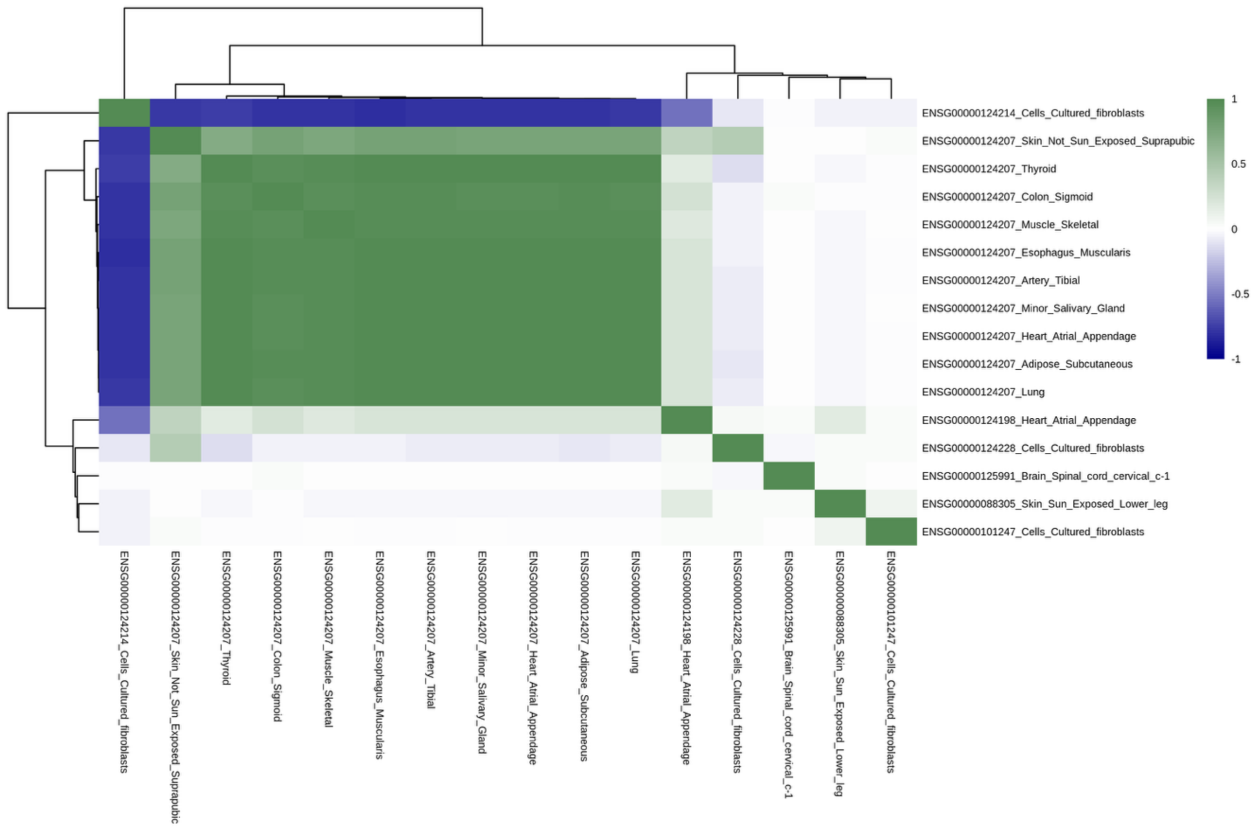


**Supplementary Figure 68. Correlation of GReX on Chromosome 20 for Sex-combined Stuttering for our MR Analyses.**

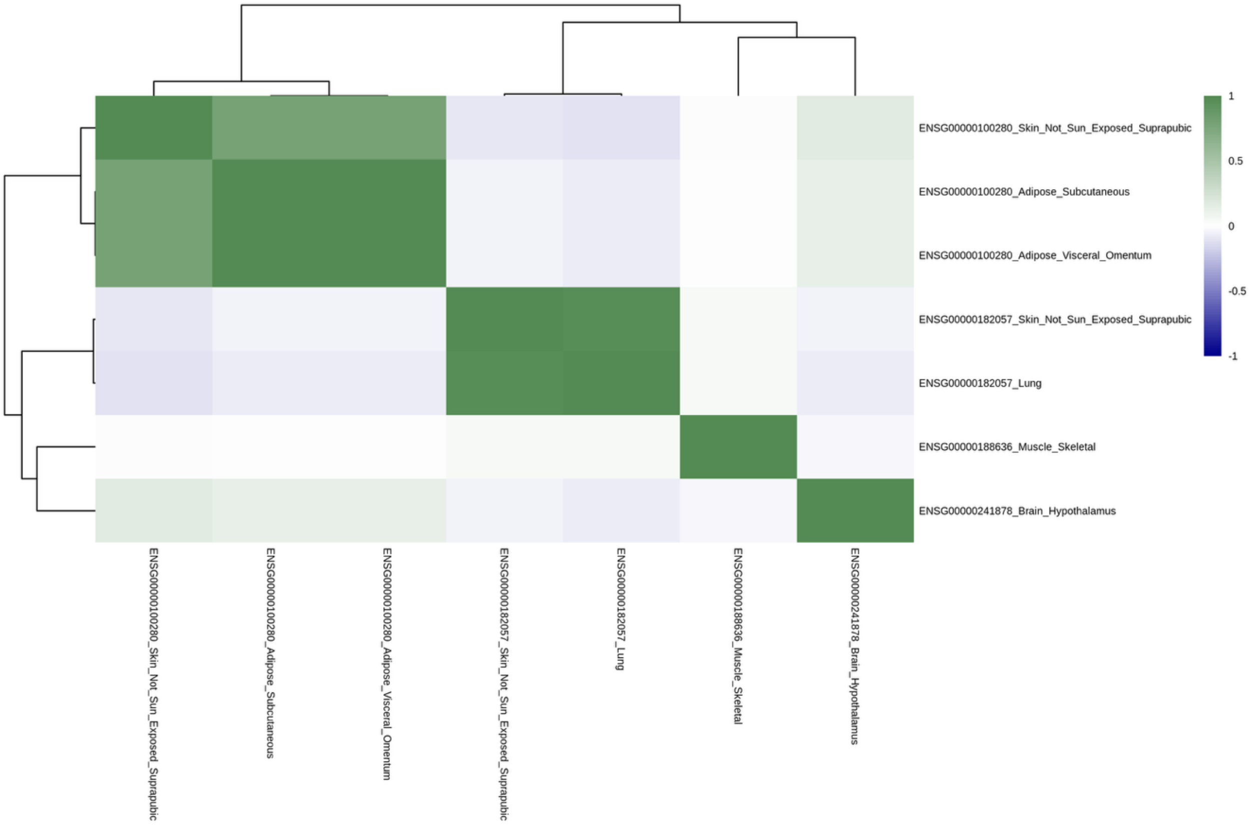


**Supplementary Figure 69. Correlation of GReX on Chromosome 22 for Sex-combined Stuttering for our MR Analyses.**
